# Cost-Benefit Analysis for Universal Cataract Surgical Coverage in India

**DOI:** 10.1101/2023.09.24.23296031

**Authors:** Anoushka Arora, Madhurima Vuddemarry, Chaitanya Reddy, Himanshu Iyer, Pushkar Nimkar, Siddhesh Zadey

**Author notes:** Corresponding author: Siddhesh Zadey BSMS, MScGH; ASAR Office Address: D2 Sai Heritage, New DP Road, Aundh, Pune, Maharashtra, India 411007. **Competing interests:** Siddhesh Zadey is the co-founding director of the Association for Socially Applicable Research (ASAR). He represents ASAR on the G4 Alliance Permanent Council as a Member. Siddhesh Zadey serves as the Chair of the Asia Working Group and the G4 Alliance, and as a Drafting Committee Member for the Maharashtra State Mental Health Policy. Other authors declare no competing interests. Funding: None.

## Abstract

**Background:** Cataract is the leading cause of curable blindness in India. We estimated enhanced coverage scale-up costs, societal value of health lost to cataract, and net benefits of covering surgery needs.

**Methods:** We conducted a retrospective cross-sectional analysis for India and its 30 states. Data was taken from Global Burden of Disease 2019, National Health Profile 2020, and National Health Accounts 2021. We used ten surgery costs. Enhanced coverage costs for the total need were calculated by multiplying prevalence by costs. For unmet need, the difference between the prevalence and surgeries achieved was used. For societal value of health lost to cataract, we multiplied disability-adjusted life-years (DALYs) with non-health GDP per capita. DALYs for unmet needs were calculated by multiplying DALYs with ratio of unmet to total needs. For net benefits, enhanced coverage costs were subtracted from societal value of health lost to cataract.

**Results:** The annual national enhanced coverage costs for covering total and unmet needs were 0.92 - 4.9 and 0.72 - 3.82 billion USD, respectively. The societal value of health lost to total and unmet cataract burden was 12.4 (95% UI: 8.9 - 16.7) and 9.7 (95% UI: 6.93 - 13) billion USD, respectively. Annual net benefits of covering total needs and unmet needs were 7.53 - 11.5 and 5.9 - 9 billion USD, respectively.

**Conclusion:** Increasing coverage for cataract surgery is financially beneficial to the public health sector. This can be done through public sector service or government-funded or social health insurance expansion.

## Introduction

Cataract is defined as “a disease of the eye in which the normally clear lens has opacified, which obscures the passage of light.”^1^ There are many causes of cataracts, including old age, metabolic diseases like diabetes, smoking, eye injury, eye surgery, use of certain medications like corticosteroids. Notably, cataract-related blindness can be prevented by the surgical removal of the opacified lens.^2^ Globally, cataract is responsible for 33% of visual impairment and 51% of blindness.^3^ It is also the leading cause of surgically curable blindness in India, the most populous country as of 2023.^4,5^ With a growing elderly population, ailments like cataracts will be on the rise in India.

To tackle the challenge of controlling cataract blindness, India runs the National Programme for Control of Blindness and Vision Impairment (NPCBVI). NPCBVI achieves this by procuring essential drugs and surgical consumables and providing financial support to government hospitals, private practitioners, non-governmental organisations, and community health workers like Accredited Social Health Activists (ASHAs) via a reimbursement system.^6^ Despite NPCBVI, India had a cataract prevalence of 30.34 million in 2018.^7^ However, the National Health Profile (NHP), which has utilised data from the National Program for Control of Blindness, noted 6.66 million cataract surgeries, which is 22% of the total prevalence.^8^ Notably, it has been recorded that 84 hospitalisations for cataracts per 100,000 population occurred in private settings, which is greater than twice that of hospitalisations in public settings (41 per 100,000 population).^9^ This preference for private settings over public settings comes at a financial cost, considering that the out-of-pocket expenditure (OOPE) for cataract surgery in private health centers was at least three times higher than in public health centers.^9^ Consequently, cataract surgeries that are being performed in hospitals not covered by the program lead to an increased OOPE.

The high OOPE in cataract care aligns with the overall Indian healthcare picture, where OOPE contributes to 63% of the total healthcare expenditure.^10^ Among other reasons, India’s persistently low government health expenditure, equaling only 1.28% of the gross domestic product (GDP) contributes to high OOPE.^11^ However, things are changing for the better since the introduction of Government Health Insurance Schemes like Pradhan Mantri Jan Arogya Yojana (PMJAY), state-led schemes, and growing private insurance packages. These insurance schemes have packages that cover the surgical cost of cataract surgeries and help reduce OOPE to some extent. PMJAY, in particular, aims to alleviate the financial burden by providing secondary and tertiary care via a combination of private and public empanelled providers. Further, the strength of a state’s public healthcare infrastructure influences the need and demand for the empanelment of private hospitals. Private hospitals accounted for 54% of total claims under PMJAY. ^12^ Leveraging both private and public facilities using insurance schemes like PMJAY can help address the high unmet need for cataract.

There are two potential approaches to address the high unmet need for cataract surgeries. First, the government could play the role of the provider and expand resources to provide surgical care in a public setting. Second, the government can play the role of the buyer so families can seek care from public or private facilities. Regardless of which method is used to fulfill the unmet need for cataract, the government must increase financial funding. In this study, we estimated the costs of enhancing coverage for cataract surgery, the societal value of health lost to cataract burden, and the net benefits of enhanced coverage.

Our analysis is conducted at the state level, which increases the robustness of our analysis. Since healthcare is a state subject and much of the healthcare planning occurs at this level, using state-level data can lead to more informed decisions that will improve cataract care regionally. The focus of our analysis is on 2018 because of the lack of recent data. We excluded Andaman and Nicobar Islands, Chandigarh, Dadra and Nagar Haveli and Daman and Diu, and Lakshadweep, due to the lack of availability of health financing indicators. We excluded Puducherry due to a lack of cataract prevalence. Additionally, we excluded Mizoram because of a lack of the National Sample Survey (NSS) mean cost per surgery.

## Methods

### Data Sources and Variables

We conducted a retrospective cross-sectional analysis for 2018 for India and its 30 states. We excluded union territories - Andaman and Nicobar, Chandigarh, Dadra and Nagar Haveli, Daman, and Diu, and Lakshadweep because we did not have their health financing indicators - government health expenditure. We excluded Puducherry due to a lack of data on cataract prevalence. For the state of Mizoram, instead of 10 surgical costs, we have used nine surgical costs, excluding the NSS mean cost per surgery.^13^ According to the NSS, no cataract surgeries were recorded for Mizoram; hence, no mean cost per surgery was recorded. However, this might be a sampling issue. Excluding the estimates for five union territories and Mizoram from our analysis could potentially underestimate the enhanced coverage scale-up costs, the societal value of health lost to cataract, and the net benefits of covering surgery needs. However, we can still demonstrate that an enhanced surgical scale-up will be cost-beneficial for the government. Data sources for all variables are listed in **Supplementary Table 1**. For better readability, we have defined key terms used in our manuscript in **Supplementary Table 2**.

### Analytic Framework

Our analysis can be broken down into three steps. First, we calculated the costs for enhanced cost coverage under two scenarios - total needs and unmet needs. Here, the ten different per-capita cataract surgical package costs used are the health intervention costs in the cost-benefit analysis. Second, we used the value-of-life-year (VLY) or the full-income approach to calculate the societal value of health lost to cataract burden. This approach utilises a macro-level perspective since we used state aggregates of averted disability-adjusted life-years (DALYs) associated with cataracts. This was also calculated under two scenarios - total and unmet need. Last, we calculated the net benefit by subtracting the costs for enhanced cost coverage from the societal value of health lost to cataract burden.

Since we used state-level costs for cataract surgical packages and DALYs, and the DALY values are obtained from modelling techniques, we conducted an uncertainty analysis instead of single-point estimates. This gives us a more accurate idea of the potential impact and increases the reliability of our results. Additionally, we conducted a sensitivity analysis taking into account the complication rate of cataract surgeries. Since not all surgeries are efficacious, we used the complication rate to calculate the new number of surgeries conducted. This was used to calculate a new cost for enhanced cataract coverage, accounting for surgical efficacy. However, the complication rates we used from the Aravind Eye Care Model will be lower than the national average due to the better quality of care provided.

### Main Analysis

First, we calculated the costs for enhanced cost coverage under two scenarios - total needs and unmet needs. The total need was defined as cataract prevalence. The unmet need was defined as the number of people with cataracts who did not get operated on, i.e., the difference between prevalence and surgeries conducted under NPCBVI. Through a literature review, we extracted ten per capita surgical package costs from multiple sources, like the NSS, PMJAY government insurance scheme, and the Aravind Eye Hospital-Madurai.^14,15^ From NSS - the nationally representative household survey, we took the state-wise mean costs for seeking cataract surgery. From PMJAY, we took six different package costs for Phacoemulsification (PHACO) and Small Incision cataract surgeries (SICS). We looked at the Aravind Eye Care Model for its efficient system of providing high-quality cataract surgery at a low cost.^15^ We considered three costs from the Aravind Eye Care Model: direct medical, patient-level indirect, which included food, travel, and lost work during the perioperative period, and societal costs, which were the sum of direct and indirect costs. ^15^

Costs and their sources are detailed in **Supplementary Table 3.** To ensure uniformity, all costs were converted from INR to USD and adjusted for inflation for the year 2020.^16,17^ To calculate the population-level enhanced coverage costs for covering total needs, we multiplied cataract prevalence by the per capita cataract surgical package costs (**Equation 1**). To calculate the enhanced coverage costs for covering the unmet needs, we multiplied the difference between prevalence and surgeries conducted with the per capita costs (**Equation 2**). Hence, we have twenty enhanced coverage costs for each Indian state and India. We conducted uncertainty propagation using the 95% uncertainty intervals associated with cataract prevalence. This accounted for potential variations in the prevalence data and made the analysis more robust and comprehensive. Mean cataract prevalence values and the 95% uncertainty intervals (upper and lower bounds) were taken from Global Burden of Disease (GBD) 2019.

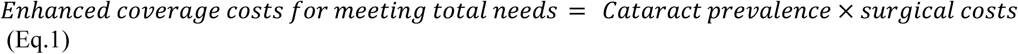

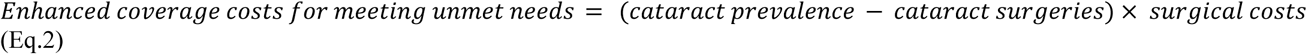

Second, the societal value of health lost to cataract burden if cataract coverage is provided was calculated using the value-of-life-year (VLY) or the full-income approach.^18,19^ The approach estimates the monetary value of each year of life, even beyond the years of active workforce contribution, considering the gains in the GDP and the population-level life expectancy due to past investments in basic public health interventions.^20^ The Lancet Commission on Investing in Health projected that for South Asia, the value of one life-year could be approximately 2.8 times the GDP per capita at a 3% discount rate.^21^ As a simpler adaptation, we multiplied non-health GDP per capita by a factor of 2.8. This gave us the life-year cost (**Equation 3**). Data for health financing indicators were extracted from the National Health Account (NHA).^22^ For India and states, we considered the national and state-level Gross Domestic Product (GDP) and Total Health Expenditure (THE). To quantify the societal value of health lost to cataract burden (value of life year), we multiplied the life-year cost with averted DALYs associated with cataracts (**Equation 4**). We assumed that surgeries are efficacious in reducing all cataract disease burdens. Mean DALY values and the 95% uncertainty intervals were taken from GBD 2019, i.e., we accounted for the smallest (lower bound) and largest (upper bound) values for DALY. To account for DALYs averted by covering unmet needs, we multiplied the DALYs averted by meeting total needs by the ratio of unmet to total needs (**Equation 5**). To account for uncertainties in the societal value of health lost to cataract burden, we propagated uncertainties from the DALY values. All calculations were conducted for two scenarios: total and unmet need.

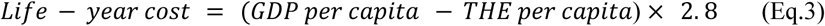

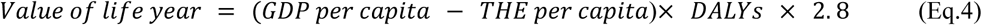

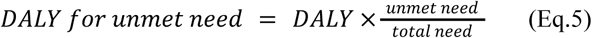

Third, we calculated the net benefits from covering total needs by subtracting the costs for covering the total needs from the societal value of health lost to cataract burden (**Equation 6**). Similarly, we calculated net benefits from costs covering the unmet needs (**Equation 7**). The error was propagated for the net benefits from covering total and unmet needs as follows. First, the lower bound value was subtracted from the mean value of the costs for enhanced cost coverage. This difference was called delta y (**Equation 8**). Similarly, the difference between the lower bound and societal value of health lost to cataract burden was called delta x (**Equation 9**). Error value for net benefits was calculated as the square root of the sum of squared delta x and squared delta y values. This was taken as delta z, and the corresponding upper and lower bound values were calculated for net benefits (**Equation 10**). The corresponding upper and lower bound values for net benefits were calculated by adding and subtracting the delta z values from the previously calculated mean value of net benefits. Since we consider a wider range of potential net benefits instead of single-point estimates, this gives us a more accurate picture of the potential impact on enhanced surgical scale-up. This increases the reliability of our results.

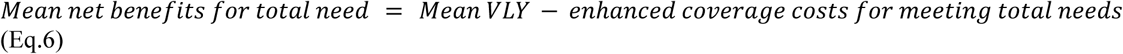

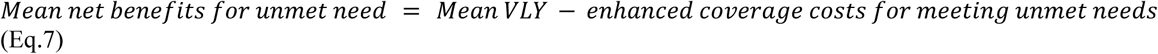

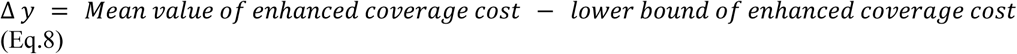

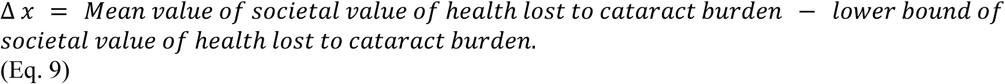

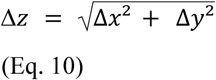

We conducted a sensitivity analysis using the complication rates from the Aravind Eye Care Model.^23^ Complication rates were used to adjust surgeries to give complication rate-adjusted unmet needs (**Equations 11-**12). Values are shown in **Supplementary Table 5**. In turn, this was used to calculate complication rate-adjusted enhanced coverage costs, societal value of health lost to cataract burden, and net benefits **(Supplementary Tables 6-9).** The percentage change in net benefits and enhanced coverage costs for meeting unmet needs is shown in **Supplementary Tables 10 and 11**.

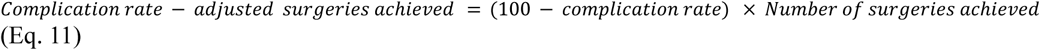

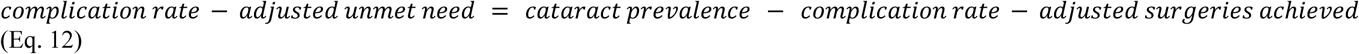

Data generated and used in this study are available in Harvard Dataverse.^24^

## Results

Annual enhanced coverage costs for covering total cataract needs in India were 0.92 - 4.9 billion USD (**Figure 1a**). While those for covering unmet needs were 0.72 - 3.82 billion USD (**Figure 1b**). We saw a 0.01% difference in enhanced coverage costs nationally on adjusting for complication rates. Despite adjusting for the complication rate, the number of surgeries achieved remains consistent. However, this could be due to the use of rates from the Aravind Eye Care Model, which underestimates the national complication rate due to the high quality of care provided.

**Figure 1:**
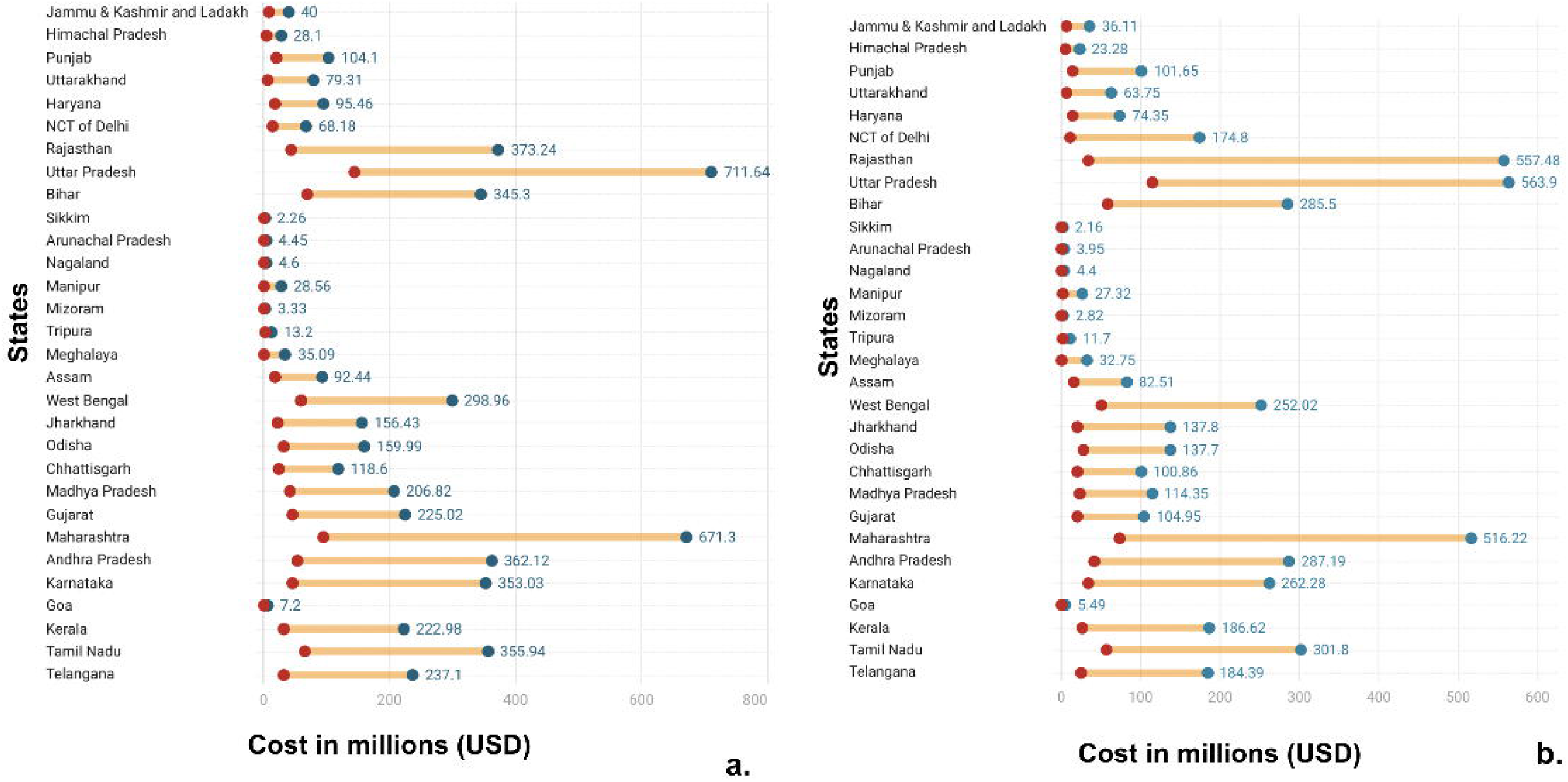
Range of Enhanced Cataract Surgical Coverage Costs across Surgical Package Costs for Meeting a. Total Needs and b. Unmet Needs. All costs are adjusted to millions of USD for the year 2020. Abbreviations: NSS - National Sample Survey, PMJAY - Pradhan Mantri Jan Arogya Yojana, PHACO - Phacoemulsification, IOL - Intraocular lens, SICS - Small Incision Cataract Surgery.

Large and populous states like Uttar Pradesh, Rajasthan, Maharashtra, and Bihar had higher enhanced coverage costs. Small Northeastern states like Sikkim and Arunachal Pradesh had lower enhanced coverage costs **(Supplementary Tables 3 and 4)**. The highest enhanced coverage costs for covering total cataract needs in India were 711.6 (95% UI: 631.2 - 809.3) million USD for Uttar Pradesh, the state with the highest need. While the lowest cost was 0.4 (95% UI: 0.3 - 0.5) million USD for Sikkim, the state with the lowest need. Whereas, for covering unmet need, the highest cost was 563.9 (95% UI: 483.5 - 661.5) million USD for Uttar Pradesh, and the lowest cost was 0.37 (95% UI: 03 - 04) million USD for Sikkim. The highest costs were calculated using PHACO with hydrophobic acrylic IOL + Glaucoma, PMJAY package, and the lowest costs were calculated using the patient (indirect costs) from the Aravind Eye Care Model.

Using the complication rate-adjusted surgeries, Uttar Pradesh had the highest enhanced coverage cost for covering unmet need, which is 610.4 million USD, using per capita costs from NSS. In contrast, using patient-indirect costs from the Aravind Eye Care Model, Sikkim had the lowest cost at 0.377 million USD. After adjusting for complication rates, Bihar had the greatest decrease in net benefits by 1360 .87 % using NSS. On the other hand, Rajasthan’s net benefits increased by 114.5 %. This indicates that policies should be additionally tailored to increase the quality of care in states with a greater percentage change after adjusting for the complication rate.

Five states - Uttar Pradesh, Maharashtra, Bihar, Tamil Nadu, and West Bengal - accounted for 47% of the national total need and 49% of the national unmet need. Scaling up surgical services in these states would reduce the cataract burden greatly. This data would also provide policymakers with an impactful starting point for working towards universal coverage. These states, except Bihar, have the largest state government expenditure, which would make the expansion of services feasible.^22^

The societal value of health lost to cataract burden for India was 12.4 (95% UI: 8.9 - 16.7) billion USD (**Figure 2a**), while that of averting unmet cataract disease burden was 9.7 (95% UI: 6.93 - 13) billion USD (**Figure 2b**). The societal value of health lost to cataract burden for total [1912.5 (95% UI: 2577 - 1370.6) million USD] and unmet needs [1470 (95% UI: 1981.7 - 1054) million USD] was highest for Maharashtra. The societal value of health lost to cataract burden for total [9.4 (95% UI: 12.7 - 6.7) million USD] and unmet need [ 8.9 (95% UI: 12.1 - 6.4) million USD] was lowest for Sikkim (**Figures 2a & b**). Although northeastern states like Sikkim, Mizoram, and Arunachal Pradesh had some of the lowest societal values of health lost to cataract and both total and unmet needs, we should not neglect scale-up in this historically underserved region with a dearth of private health facilities and poor access to health.^25^

**Figure 2:**
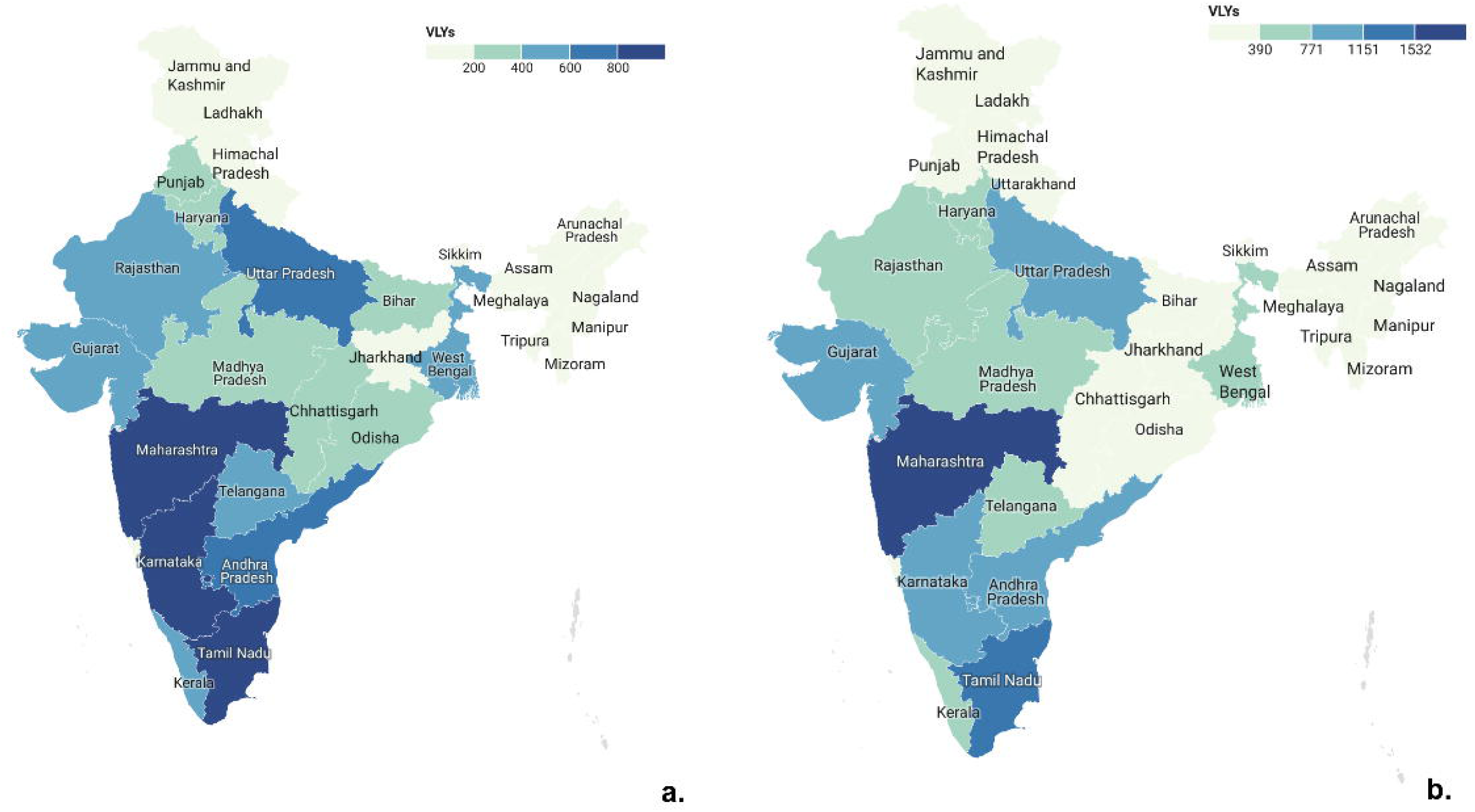
Societal Value of Health Lost to Cataract Burden Measured Using the Value of Life-Year (VLY) Approach under a. Total Needs and b. Unmet Needs Scenarios. Values in million USD.

The annual net benefits from covering total needs in India ranged from 7.53 (95%UI: 6.95 to 8.10) billion USD to 11.5 (95% UI: 11.4 to 11.6) billion USD while those from covering unmet needs ranged from 5.9 (95%UI: 3.11 to 8.7) billion USD to 9 (95% UI: 6.2 to 11.8) billion USD (**Figure 3**). Twenty-seven (90%) states depicted net benefits for both total and unmet need scenarios. State-wise net benefits/losses are presented in **Supplementary Figures 1-30**. The highest net benefits were seen in Maharashtra for total (1816.8 million USD) and unmet need (1397.1 million USD) using the patient (indirect) costs from the Aravid Eye Care Model **(Supplementary Figure 24).** The lowest net benefits were seen in Meghalaya for total (7.76 million USD) and unmet need (7.25 million USD) using the PMJAY PHACO with hydrophobic acrylic IOL + glaucoma package **(Supplementary Figure 16)**. Bihar **(Supplementary Figure 9)**, Meghalaya **(Supplementary Figure 16)**, and Manipur **(Supplementary Figure 13)** had net losses for only certain per capita surgical package costs.

**Figure 3:**
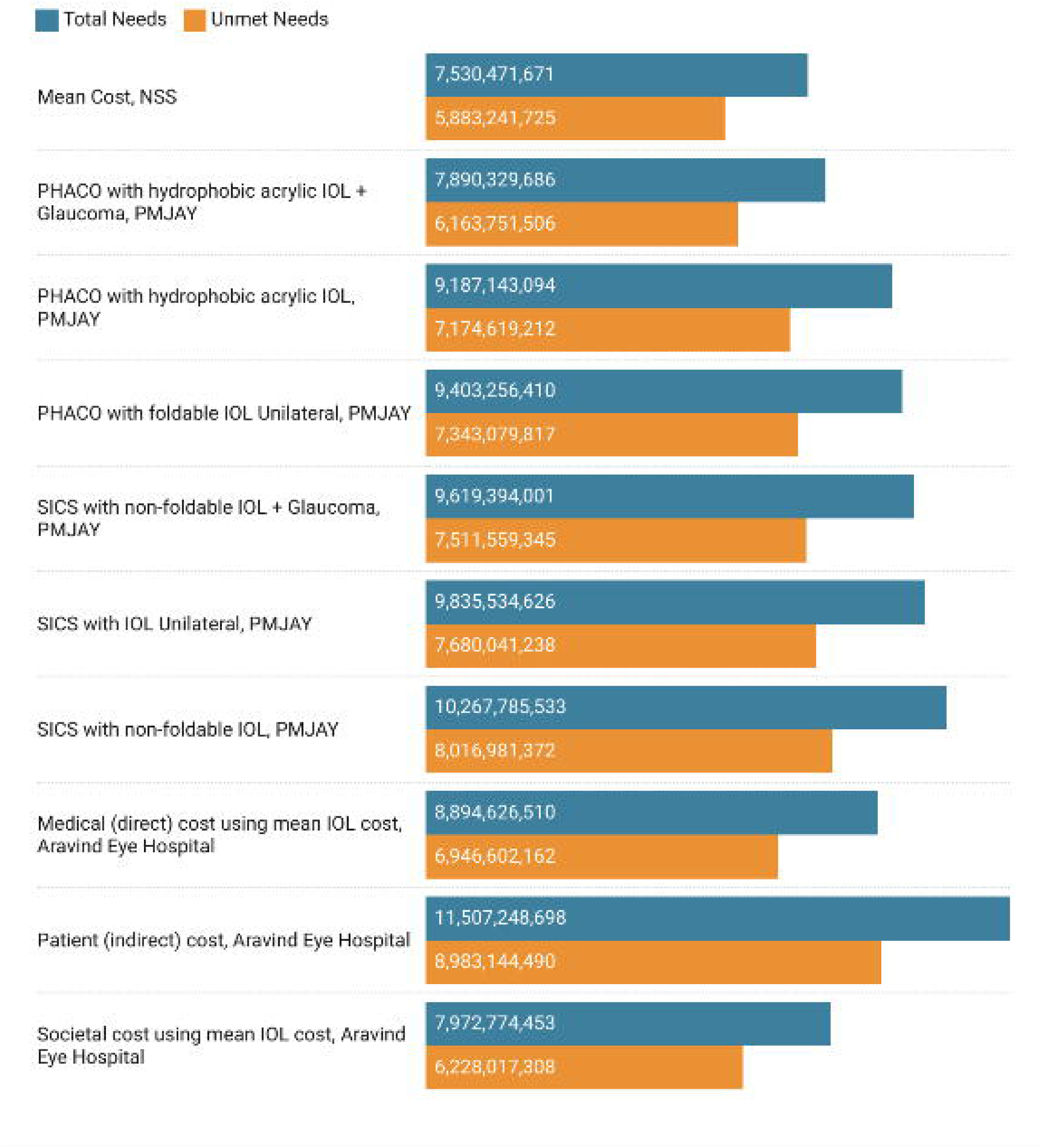
Net Benefits for Meeting Total and Unmet Needs for Universal Cataract Surgical Coverage in India. All costs are adjusted to USD for the year 2020. Positive values depict net benefit while negative values depict net loss. Abbreviations: NSS - National Sample Survey, PMJAY - Pradhan Mantri Jan Arogya Yojana, PHACO - Phacoemulsification, IOL - Intraocular lens, SICS - Small Incision Cataract Surgery

Meghalaya had net losses for total (20.3 million USD) and unmet need (19 million USD) when using one surgical package cost (mean cost, NSS) out of ten. Similarly, using the same surgical package cost, Manipura had net losses for total (12.5 million USD) and unmet need (12 million USD). Notably, Bihar, with one of the highest unmet needs, had net losses for two (PMJAY PHACO with hydrophobic acrylic IOL + glaucoma package, societal cost using mean IOL from Aravind Eye Care Model) out of the ten surgical package costs used. Considering that addressing Bihar’s high unmet need is important from a policy standpoint, the range of surgical costs examined in our analysis would be vital for determining the most cost-beneficial approach.

## Discussion

Increasing cost coverage is beneficial for the government. Costs for enhanced cataract coverage were calculated to cover total and unmet needs, showing wide interstate variability due to differences in population size, disease burden, and GDP values. Expectedly, states like Maharashtra, Uttar Pradesh, and Rajasthan have greater costs for coverage expansion than those for northeastern states like Sikkim and Nagaland. However, in most scenarios, 27 states in India had net benefits from the enhanced cost coverage of cataract surgeries. Hence, investing in universal cataract surgical coverage caters to the rising burden.

Globally, in 2020, cataract was the leading cause of blindness, with greater prevalence in South and Southeast Asia and Oceania. ^26^ The burden can be tackled through corrective surgeries for cataract are cost-effective. Overall, lower-middle-income countries (LMICs) like India and Nepal had better cost-effectiveness ratios (cost per DALY averted or quality of life years {QALY} gained) than high-income countries (HICs), noting the need for prioritization. ^26^ However, access to surgeries varies globally, which can be assessed by cataract surgical rate (CSR), defined as cataract operations performed per million population in one year. In HICs like the United States, Australia, and Japan, CSR ranged from 10,000 to 40,000. ^3^ In contrast, in countries in Latin America and Asia, the CSR ranged from 500 to 2,000. ^3^ This indicates poor access in LMICs compared to HICs. Given the significant return on investment, scaling up cataract surgeries globally is financially beneficial. ^27^

The societal losses due to cataracts, as a surgically preventable cause of blindness, and the cost-effectiveness of covering case detection strategies have been studied in India.^14,28^ Our study differs from previous cost-effectiveness modeling and cost-of-illness studies. ^29^ We rely on existing individual-level studies to compile a library of benefit-cost differences that can help policymakers understand that, at the aggregate (systemic and societal) level, the costs to the government for financing universalized cataract surgical coverage are lower than the benefits to society from averting the disease burden due to untreated cataract. Additionally, the present analysis estimates the state-wise economic benefits of expanding the cost coverage of cataract surgeries in India, which was previously missing. The net benefits are substantial, which should be an impetus for policymakers to expand and provide universal cataract coverage.

According to the NPCBVI (2015-2019) Summary Report, the private sector conducted 57.85% of cataract surgeries, whereas the public sector conducted 39.3% of surgeries.^5^ Further, OOPE was the major barrier for patients seeking surgery. This conundrum can be resolved by either the government directly providing the service or paying other service providers to offer the service at a lower cost. Hence, expanding NPCBVI by increasing direct payments to service providers in the government setup can improve access to government setups and ease the financial burden of the care seekers.^5^ Moreover, investments can be utilised to expand services in the public and private sectors to achieve universal coverage.^30^

Currently, 30% of the population is completely uninsured, resulting in several households falling into poverty.^10^ Households’ ability to handle the high OOPE depends on their income level. For instance, low-income households either choose not to get cataract surgery, opt for subsidised procedures at the public hospitals, or at hospitals enpanelled under the government health insurance schemes. Whereas, middle- and high-income households have an additional option to undergo the procedure without government insurance at the private hospitals. The existing insurance schemes can cover 70% of the population.^10^ However, the real population coverage is less due to the overlap of eligibility across PMJAY, Central Government Health Schemes, and private insurance packages.

An increase in investment in PMJAY will increase coverage of surgeries for the uninsured population due to a decrease in OOPE, leading to universal cataract surgical coverage. Investing in cataract surgery insurance can bring new providers to places lacking access, as previously observed in Rashtriya Swasthya Bima Yojana (RSBY).^31,32^ RSBY is a government-funded health insurance scheme provided to Below Poverty Line (BPL) families and 11 other categories of unorganized workers in any Government empaneled public or private hospital.^33^ RSBY allocated funds to states like Jharkhand and Chhattisgarh, which resulted in an uptick in private practitioners setting up clinics in these states. Consequently, when PMJAY was initiated, 99% of the total cataract surgery claims were raised from private hospitals in Chhattisgarh and Jharkhand. Contrarily, states such as Madhya Pradesh, where government hospitals provide care, had low utilisation of the PMJAY cataract surgery package. Low utilisation of PMJAY packages in government hospitals can be a result of coverage provided by NPCBVI. Expectedly, demand for PMJAY cataract surgery packages increased significantly even among smaller clinics, improving access to beneficiaries.^14^ Additionally, claims in Chandigarh, Nagaland, Sikkim, and Tripura were from public hospitals. This can be attributed to only a few private eye care hospitals that existed in the northeastern states.^34^ A reason for this could be that the private sector fills in for the responsibility for meeting demand in states where there is a lack of public sector resources. However, it is the public sector that helps create the demand through awareness and providing free care. Hence, enhanced investments can provide cataract surgical coverage by public sector expansion or government-funded health insurance that covers the care provided at the empanelled public and private hospitals.

Since our study looks at state-wise benefits over a range of costs, it is helpful for local policymakers. By evaluating the strength of a state’s public health infrastructure, government health expenditures, and the availability of private facilities, we can determine which states would benefit from expanding insurance coverage and which would gain from expanding public/government services. Since Uttar Pradesh, Maharashtra, Bihar, Tamil Nadu, and West Bengal accounted for almost 50% of the national unmet need, prioritizing them would lead to a significant decrease in the national cataract burden. Out of these states, Uttar Pradesh, Maharashtra, Tamil Nadu, and West Bengal have the largest state health expenditures. ^22^ Hence, scale-up can be managed at the state level with little coordination with the central government via public service expansion. Since Bihar has a high unmet need and low state health expenditure, it will require significant central and state-level coordination for an effective scale-up with government health insurance coverage expansion. Conversely, Chhattisgarh and northeastern states such as Sikkim and Meghalaya have a low unmet need for healthcare. However, they face challenges due to limited access to healthcare services and a shortage of private facilities. ^25,35^ Therefore, these regions will require support from the central government to develop healthcare infrastructure and expand human resources at the public hospitals, to expand services, and also ensure greater government health insurance coverage. For states like Gujarat with high government state health expenditure and robust health infrastructure, scale-up can be managed effectively at the state level. Additionally, it is also important to consider the percentage of the population and the rate of increase in the ageing population. Since states like Kerala have the highest aging population, scaling up cataract surgeries should be a priority. ^36^

While PMJAY increases coverage for the uninsured population, there are some drawbacks to it. A significant portion of grievances were related to demands for payment to receive treatment. Studies conducted show that enrollment under the PMJAY insurance scheme does not lead to a decrease in OOPE. ^37^ This can be attributed to two things. First, a lack of regulatory mechanisms that prevent private hospitals from charging extra money. Second, a lack of awareness of approved hospitals, covered services, where to receive the treatment, treatment costs, and card balance. ^38^

To our knowledge, this is the first state-wise pan-India analysis of the costs and benefits of universal cataract surgical coverage. We provide a library of over 900 estimates under multiple scenarios and cost assumptions. We also ensured robustness through uncertainty propagation. However, the study has the following limitations. First, we assumed that surgeries could treat the entire disease burden, which may not be true. This could lead to an overestimation of the net benefits of cataract surgical scale-up. Future studies should consider the post-surgery site restoration rate while calculating estimates. Second, we adjusted for the complication rate using data from the Aravind Eye Care Model. Given Aravind’s high standards of care, these rates likely conservatively underestimate the true complication rate. Future studies should utilize national complication rates for a more accurate assessment. Third, we only analyzed the annual net benefits of 2018 and did not project future benefits. Nonetheless, the overtime benefits for future years would only rise further. Hence, our findings could be considered as the lower bound of the net benefits.

## Data Availability

All data produced are available online at https://doi.org/10.7910/DVN/DTEZXS

## Acknowledgements

The authors would like to thank Ram Pachpor and Tejali Gangane for their assistance with data and Dr. Sweta Dubey for their helpful comments.

## Declarations

### Ethics approval and consent to participate

Not applicable

### Consent for publication

Not applicable

## Authors’ contributions

***Study concept and design:*** Siddhesh Zadey

***Acquisition, analysis, or interpretation of data:*** Anoushka Arora, Pushkar Nimkar, Himanshu Iyer, Siddhesh Zadey

***Drafting of the manuscript:*** Anoushka Arora, Madhurima Vuddemarry, Chaitanya Reddy

***Critical revision of the manuscript for important intellectual content:*** All authors

***Statistical analysis:*** Anoushka Arora, Pushkar Nimkar

***Administrative, technical, or material support:*** Siddhesh Zadey

***Study supervision:*** Siddhesh Zadey

## Dataset Availability

Data used and generated in this manuscript can be found in the associated repository at Harvard Dataverse (https://doi.org/10.7910/DVN/DTEZXS)

