## Supplement for "Cost-Benefit Analysis for Universal Cataract Surgical Coverage in India"

**Supplementary Table 1.** Variables Used for the Analysis and their Sources.

**Supplementary Table 2**. Definition of Key Terminology.

**Supplementary Table 3.** Cost of Cataract Surgery Adjusted for Inflation for the Year 2020 and Converted to USD.

**Supplementary Table 4.** Enhanced Cataract Coverage Costs for Meeting Total Needs for Cataract Surgery.

**Supplementary Table 5.** Enhanced Cataract Coverage Costs for Meeting Unmet Needs for Cataract Surgery.

**Supplementary Table 6.** Complication Rate-Adjusted Surgeries and Unmet Needs.

**Supplementary Table 7.** Complication Rate-Adjusted Enhanced Coverage Costs for Meeting Unmet Needs for Cataract Surgery.

**Supplementary Table 8.** Complication Rate-Adjusted Economic Benefits for Meeting Unmet Needs for Cataract Surgery.

**Supplementary Table 9.** Complication Rate-Adjusted Net Benefits for Meeting Unmet Needs for Cataract Surgery.

**Supplementary Table 10.** Percentage Change in Net Benefits for Meeting Unmet Needs for Cataract Surgery After Adjusting for Complication Rate.

**Supplementary Table 11.** Percentage Change in Enhanced Coverage Costs for Meeting Unmet Needs for Cataract Surgery After Adjusting for Complication Rate.

**Supplementary Figure 1.** Net Benefits for Meeting Total and Unmet Needs for Universal Cataract Surgical Coverage in Jammu & Kashmir and Ladakh.

**Supplementary Figure 2.** Net Benefits for Meeting Total and Unmet Needs for Universal Cataract Surgical Coverage in Himachal Pradesh.

**Supplementary Figure 3.** Net Benefits for Meeting Total and Unmet Needs for Universal Cataract Surgical Coverage in Punjab.

**Supplementary Figure 4.** Net Benefits for Meeting Total and Unmet Needs for Universal Cataract Surgical Coverage in Uttarakhand.

**Supplementary Figure 5.** Net Benefits for Meeting Total and Unmet Needs for Universal Cataract Surgical Coverage in Haryana.

**Supplementary Figure 6.** Net Benefits for Meeting Total and Unmet Needs for Universal Cataract Surgical Coverage in Delhi.

**Supplementary Figure 7.** Net Benefits for Meeting Total and Unmet Needs for Universal Cataract Surgical Coverage in Rajasthan.

**Supplementary Figure 8.** Net Benefits for Meeting Total and Unmet Needs for Universal Cataract Surgical Coverage in Uttar Pradesh.

**Supplementary Figure 9.** Net Benefits for Meeting Total and Unmet Needs for Universal Cataract Surgical Coverage in Bihar.

**Supplementary Figure 10.** Net Benefits for Meeting Total and Unmet Needs for Universal Cataract Surgical Coverage in Sikkim.

**Supplementary Figure 11.** Net Benefits for Meeting Total and Unmet Needs for Universal Cataract Surgical Coverage in Arunachal Pradesh.

**Supplementary Figure 12.** Net Benefits for Meeting Total and Unmet Needs for Universal Cataract Surgical Coverage in Nagaland.

**Supplementary Figure 13.** Net Benefits for Meeting Total and Unmet Needs for Universal Cataract Surgical Coverage in Manipur.

**Supplementary Figure 14.** Net Benefits for Meeting Total and Unmet Needs for Universal Cataract Surgical Coverage in Mizoram.

**Supplementary Figure 15.** Net Benefits for Meeting Total and Unmet Needs for Universal Cataract Surgical Coverage in Tripura.

**Supplementary Figure 16.** Net Benefits for Meeting Total and Unmet Needs for Universal Cataract Surgical Coverage in Meghalaya.

**Supplementary Figure 17.** Net Benefits for Meeting Total and Unmet Needs for Universal Cataract Surgical Coverage in Assam.

**Supplementary Figure 18.** Net Benefits for Meeting Total and Unmet Needs for Universal Cataract Surgical Coverage in West Bengal.

**Supplementary Figure 19.** Net Benefits for Meeting Total and Unmet Needs for Universal Cataract Surgical Coverage in Jharkhand.

**Supplementary Figure 20.** Net Benefits for Meeting Total and Unmet Needs for Universal Cataract Surgical Coverage in Odisha.

**Supplementary Figure 21.** Net Benefits for Meeting Total and Unmet Needs for Universal Cataract Surgical Coverage in Chhattisgarh.

**Supplementary Figure 22.** Net Benefits for Meeting Total and Unmet Needs for Universal Cataract Surgical Coverage in Madhya Pradesh.

**Supplementary Figure 23.** Net Benefits for Meeting Total and Unmet Needs for Universal Cataract Surgical Coverage in Gujarat.

**Supplementary Figure 24.** Net Benefits for Meeting Total and Unmet Needs for Universal Cataract Surgical Coverage in Maharashtra.

**Supplementary Figure 25.** Net Benefits for Meeting Total and Unmet Needs for Universal Cataract Surgical Coverage in Andhra Pradesh.

**Supplementary Figure 26.** Net Benefits for Meeting Total and Unmet Needs for Universal Cataract Surgical Coverage in Karnataka.

**Supplementary Figure 27.** Net Benefits for Meeting Total and Unmet Needs for Universal Cataract Surgical Coverage in Goa.

**Supplementary Figure 28.** Net Benefits for Meeting Total and Unmet Needs for Universal Cataract Surgical Coverage in Kerala.

**Supplementary Figure 29.** Net Benefits for Meeting Total and Unmet Needs for Universal Cataract Surgical Coverage in Tamil Nadu.

**Supplementary Figure 30.** Net Benefits for Meeting Total and Unmet Needs for Universal Cataract Surgical Coverage in Telangana.

**Supplementary Tables**

**Supplementary Table 1: Variables Used for the Analysis and their Sources.**

| Variable | Source |
|---|---|
| Cataract prevalence (mean, lower bound, and upper bound) | Global Burden of Disease (GBD) Study 2019.<br>Institute for Health Metrics and Evaluation (IHME) |
| Cataract disability-adjusted life-year (DALYs) (mean, lower bound, and upper bound) | Global Burden of Disease (GBD) Study 2019.<br>Institute for Health Metrics and Evaluation (IHME) |
| Number of cataract surgeries performed | Report: National Health Profile (NHP) 2020<br>Table No: 3.2.2<br>Variable: Cataract Surgeries Achievement |
| Mean total cost per surgery | National Sample Survey (NSS) 2017-18 |
| PM-JAY cataract package costs | Report: Cataract Management under Ayushman Bharat Pradhan Mantri Jan Arogya Yojana<br>(PM-JAY)<br>Working paper - 003<br>Figure: 2<br>Variable(s): Cataract Management Packages under PM-JAY (per capita surgical costs) |
| Per surgery costs of cataract surgery at Aravind Eye Hospital–Madurai, July 2013 | Report: A Sustainable Model For Delivering High-Quality, Efficient Cataract Surgery In Southern India.<br>Table: Exhibit 1<br>Variables: Societal cost using mean IOL cost, medical (direct) cost using mean IOL cost, and patient (indirect) cost. |
| Government Health Financing Indicators for all the states and UTs with Legislature (2017-18) | Report: National Health Accounts 2021.<br>Table: A-7 - Government Health Financing indicators for all the states and UTs with Legislature (2017-18)<br>Variable: Gross Health Expenditure (GHE), Gross State Domestic Product (GSDP), GHE as % of GSDP, Per capita Total Gross Health Expenditure (TGHE), Population |

**Supplementary Table 2**. **Definition of Key Terminology.**

| Key Terms | Definition |
|---|---|
| Unmet Need | Number of people with cataract who did not get cataract surgery |
| Enhanced coverage scale-up costs | The total cost of cataract surgeries to eliminate cataract |
| Societal value of health lost | The loss of economic productivity due to cataract |
| Value of Life-Year | Monetary value of each year of life, even beyond the years of active workforce contribution, considering the gains in the GDP and the population-level life expectancy |
| Net Benefits | Enhanced coverage scale-up costs subtracted from the societal value of health lost |

**Supplementary Table 3: Cost of Cataract Surgery Adjusted for Inflation for the Year 2020 and Converted to USD.** IOL-Intraocular lens, NSS - National Sample Survey, PHACO -Phacoemulsification, PMJAY - Pradhan Mantri Jan Arogya Yojana, SICS -Small Incision Cataract Surgery.

| Source | Cost and year | Cost in INR (2020) | Cost in USD (2020) | Patient/Provider Cost |
|---|---|---|---|---|
| Mean cost, NSS | INR 10038.30242 (2017) | 11962.75 | 161.436 | Direct and indirect costs (patient's perspective) |
| PMJAY Cataract Surgery (PHACO) with hydrophobic acrylic IOL + Glaucoma | INR 10500 (2019) | 11083.94 | 149.577 | Direct costs (provider's perspective) |
| PMJAY Cataract Surgery (PHACO) with hydrophobic acrylic IOL | INR 7500 (2019) | 7917.10 | 106.84 | Direct costs (provider's perspective) |
| PMJAY Cataract Surgery (PHACO) with foldable IOL Unilateral | INR 7000 (2019) | 7389.30 | 99.7179 | Direct costs (provider's perspective) |
| PMJAY Cataract Surgery (SICS) with non-foldable IOL + Glaucoma | INR 6500 (2019) | 6861.49 | 92.595 | Direct costs (provider's perspective) |

| | | | | |
|---|---|---|---|---|
| PMJAY Cataract Surgery (SICS) with IOL Unilateral | INR 6000 (2019) | 6333.68 | 85.472 | Direct costs (provider's perspective) |
| PMJAY Cataract Surgery (SICS) with non-foldable IOL | INR 5000 (2019) | 5278.07 | 71.227 | Direct costs (provider's perspective) |
| Per surgery costs of cataract surgery at Aravind Eye Hospital–Madurai, July 2013<br><br>Societal cost using mean IOL cost | USD 120.24 (2016) | 10882.62 | 146.86 | Direct (patient's and provider's perspective) and patient-indirect cost |
| Per surgery costs of cataract surgery at Aravind Eye Hospital–Madurai, July 2013<br><br>Medical (direct) cost using mean IOL cost | USD 95.37 (2016) | 8631.401 | 116.48 | Direct costs (patient's and provider's perspective) |
| Per surgery costs of cataract surgery at Aravind Eye Hospital–Madurai, July 2013<br><br>Patient (indirect) cost | USD 24.87 (2016) | 2251.219 | 30.38 | Patient-indirect Costs |

**Supplementary Table 4: Enhanced Cataract Coverage Costs for Meeting Total Needs for Cataract Surgery.** All costs are adjusted to millions of USD for the year 2020. NSS - National Sample Survey, PMJAY - Pradhan Mantri Jan Arogya Yojana, PHACO -Phacoemulsification, IOL-Intraocular lens, SICS -Small Incision Cataract Surgery.

| States | Mean Cost, NSS | PHACO with hydrophobic acrylic IOL + Glaucoma, PMJAY | PHACO with hydrophobic acrylic IOL, PMJAY | PHACO with foldable IOL Unilateral, PMJAY | SICS with non-foldable IOL + Glaucoma, PMJAY | SICS with IOL Unilateral, PMJAY | SICS with non-foldable IOL, PMJAY | Societal cost using mean IOL cost, Aravind Eye Hospital | Medical (direct) cost using mean IOL cost, Aravind Eye Hospital | Patient (indirect) cost, Aravind Eye Hospital |
|---|---|---|---|---|---|---|---|---|---|---|
| Jammu & Kashmir and Ladakh | 36.66 (32.29, 41.55) | 40 (35.22,45.33) | 28.57 (25.16, 32.38) | 26.66 (23.48, 30.22) | 24.76 (21.80, 28.06) | 22.90 (20.13, 25.90) | 19.04 (17, 21.50) | 39.27 (34.59, 44.51) | 31.11 (27.43, 35.30) | 8.13 (7.15, 9.20) |
| Himachal Pradesh | 19.41(16.80,22.28) | 28.10 (24.30,32.23) | 20.05 (17.36, 23.02) | 18.71 (16.20, 21.48) | 17.38 (15.04, 19.95) | 16.04 (13.90, 18.14) | 13.37 (11.57, 15.34) | 27.57 (23.87, 31.65) | 21.90 (18.93, 25.10) | 5.70 (4.93, 6.54) |
| Punjab | 15.60 (13.46,18.04) | 104.10 (89.83,120.40) | 74.33 (64.17, 86) | 69.37 (59.90, 80.26) | 64.42 (55.61, 74.52) | 59.46 (51.33, 69.80) | 49.60 (42.80, 57.33) | 102.18 (88.21, 118.21) | 81.04 (69.95, 93.75) | 21.12 (18.21, 24.45) |
| Uttarakhand | 79.31 (69.70,90.50) | 38.61 (33.91,44.10) | 27.60 (24.22, 31.49) | 25.74 (22.60, 29.39) | 23.90 (21, 27.29) | 22.06 (19.37, 25.19) | 18.38 (16.14, 21) | 37.91 (33.3, 43.29) | 30.07 (26.40, 34.33) | 7.84 (6.88, 8.95) |
| Haryana | 90.60 (79.61, 102.70) | 95.46 (83.90,108.20) | 68.18 (59.91, 77.27) | 63.65 (55.91, 72.12) | 59.09 (51.92, 67) | 54.60 (47.93, 61.82) | 45.45 (40, 51.51) | 93.73 (82.36, 106.23) | 74.34 (65.31, 84.25) | 19.38 (17.03, 21.97) |

| | | | | | | | | | | |
|---|---|---|---|---|---|---|---|---|---|---|
| NCT of Delhi | 16.06(14.10,18.23) | 68.18 (59.81,77.36) | 48.70 (42.72, 55,26) | 45.45 (39.88, 51.60) | 42.21 (37.02, 48.20) | 39 (34.18, 44.21) | 32.47 (28.48, 36.90) | 66.95 (58.73, 75.9) | 52.10 (46.58, 60.24) | 13.84 (12.12, 15.71) |
| Rajasthan | 373.24 (326.27,424.90) | 217.02 (189.71, 247.05) | 155.01 (135.50, 176.46) | 144.70 (126.47, 164.70) | 134.34 (117.44, 152.93) | 124.01 (108.40, 141.17) | 34 (30.05, 38.53) | 213.08 (186.27, 242.57) | 169.00 (147.73, 192.38) | 44.07 (38.53, 50.17) |
| Uttar Pradesh | 624.42 (562.80, 721.46) | 711.64 (631.24,809.30) | 508.31 (450.89, 578.05) | 474.43 (420.83, 539.95) | 440.54 (391, 501.10) | 406.65 (360.70, 462.44) | 34 (30.05, 39.10) | 698.72 (619.78, 94.58) | 554.18 (491.56, 630.20) | 144.54 (128.20, 164.34) |
| Bihar | 285 (250.07, 324.41) | 345.30 (302.94,393) | 246.62 (216.38, 280.71) | 230.18 (201.96, 262) | 213.74 (187.53, 243.28) | 197.30 (173.10, 224.57) | 164.41 (144.25, 187.14) | 339.01 (297.44, 385.87) | 268.10 (235.91, 306.04) | 70.12 (61.52, 79.82) |
| Sikkim | 2.26 (1.99, 2.59) | 1.94 (1,71,2.21) | 1.39 (1.22, 1.60) | 1.29 (1.14, 1.47) | 1.20 (1.06, 1.37) | 1.11 (0.97, 1.26) | 0.9 (0.81, 1.05) | 1.91 (1.68, 2.18) | 1.54 (1.33, 1.72) | 0.39 (0.34, 0.45) |
| Arunachal Pradesh | 4.45 (3.91,5.06) | 3.15 (2.80,3.60) | 2.25 (1.98, 2.60) | 2.10 (1.85, 2.39) | 1.95 (1.71, 2.22) | 1.80 (1.56, 2.05) | 1.50 (1.32, 1.71) | 3.1 (2.73, 3.53) | 2.45 (2.16, 2.80) | 0.64 (0.56, 0.73) |
| Nagaland | 3.61 (3.18,4.08) | 4.60 (4.06,5.20) | 3.28 (2.89, 3.71) | 3.06 (2.70, 3.46) | 2.85 (2.51, 3.21) | 2.63 (2.31, 2.97) | 2.19 (1.93, 2.47) | 4.52 (3.99, 5.11) | 3.58 (3.16, 4.05) | 0.93 (0.82, 1.05) |
| Manipur | 28.56 (25.10,32.66) | 9.79 (8.60, 11.88) | 6.99 (6.14, 3.71) | 6.52 (5.73, 7.45) | 6.05 (5.32, 6.92) | 5.59 (4.91, 6.39) | 4.65 (4.09, 5.32) | 9.61 (8.44, 10.99) | 7.62 (6.69, 8.71) | 1.98 (1.74, 2.27) |
| Mizoram | 0 | 3.33 | 2.37 (2.08, | 2.22 | 2.06 (1.80, | 1.90 (1.66, | 1.58 (1.39, | 3.27 (2.87, | 2.60 (2.27, | 0.67 (0.60, |

| | | | | | | | | | | |
|---|---|---|---|---|---|---|---|---|---|---|
| | | (2.91, 3.80) | 2.71) | (1.94, 2.53) | 2.35) | 2.17) | 1.81) | 3.74) | 3.01) | 0.77) |
| Tripura | 3.47 (3.06,3.96) | 13.20 (11.61, 14.20) | 9.41 (8.29, 10.71) | 8.80 (7.74, 10) | 8.16 (7.19, 9.28) | 7.53 (6.63, 8.56) | 6.27 (5.53, 7.14) | 12.94 (11.41, 12.72) | 10.2 (9.04, 11.60) | 2.67 (2.35, 3.04) |
| Meghalaya | 35.09 (30.83,39.89) | 7.01 (6.16, 7.98) | 5 (4.40, 5.70) | 4.67 (4.11, 5.31) | 4.34 (3.81, 4.93) | 4 (3.35, 4.55) | 3.33 (2.39, 3.79) | 6.89 (6.06, 7.83) | 5.40 (4.80, 6.21) | 1.42 (1.25, 1.62) |
| Assam | 60 (52.80,68.07) | 9.24 (8.14, 10.49) | 66.03 (58.15, 74.97) | 61.60 (54.27, 70.20) | 57.22 (50.40, 65) | 52.83 (46.52, 60) | 44.02 (38.77, 50.01) | 90.76 (79.93, 103.06) | 71.98 (63.40, 81,70) | 18.72 (16.53, 21.31) |
| West Bengal | 277.28 (242.24,316.88) | 298.96 (261.17, 341.62) | 215.35 (186.55, 244.01) | 199.30 (174.11, 227.80) | 185.50 (161.67, 211.48) | 17.08 (14.92, 19.52) | 142.35 (124.36, 162.70) | 293.52 (256.43, 335.42) | 232.80 (203.38, 266.03) | 60.71 (53.04, 69.38) |
| Jharkhand | 156.43 (137.46,178.01) | 114.60 (100.70, 130.39) | 81.85 (71.92, 93.13) | 76.39 (67.13, 86.93) | 70.94 (62.33, 80.73) | 65.48 (57.54, 74.51) | 54.57 (48.05, 62.09) | 112.52 (98.87, 128.03) | 89.24 (78.41, 101.54) | 23.27 (20.45, 26.48) |
| Odisha | 136.63 (119,155.66) | 159.99 (139.31, 182.25) | 114.27 (99.50, 130.18) | 106.65 (92.87, 121.50) | 99.04 (86.24, 112.82) | 91.42 (79.60, 104.14) | 76.18 (66.33, 86.78) | 157.08 (136.78, 17895) | 124.60 (108.48, 141.92) | 32.49 (28.29, 37.01) |
| Chhattisgarh | 100.25 (88.41,112.80) | 118.60 (104.60, 133.45) | 84.71 (74.71, 95.32) | 79.06 (69.73, 89) | 73.42 (64.75, 82.61) | 67.72 (59.80, 76.25) | 56.47 (50.01, 63.54) | 116.45 (102.7, 131.03) | 92.35 (81.45, 103.92) | 24.08 (21.24, 27.10) |
| Madhya Pradesh | 135.35 (117.77,154.3 | 206.82 (179.94, | 147.72 (128.53, | 137.89 (120.01, | 128.03 (111.39, 145) | 118.18 (102.82, | 98.50 (86.54, 112.29) | 203.07 (176.68, | 161.05 (140.12, | 42.01 (36.54, |

| | | | | | | | | | | |
|---|---|---|---|---|---|---|---|---|---|---|
| | 3) | 235.82) | 168.44) | 157.21) | | 134.80) | | 231.54) | 183.64) | 48.00) |
| Gujarat | 165.21 (145.60,187.38) | 225.02 (198.29, 255.22) | 160.72 (141.63, 182.30) | 150.03 (132.19, 170.15) | 139.28 (122.90, 158) | 128.60 (113.30, 146) | 107.15 (94.42, 121.53) | 220.93 (194.69, 250.59) | 175.23 (154.41, 199) | 45.70 (40.27, 51.83) |
| Maharashtra | 671.30 (590.05,763.40) | 471.38 (414.32, 536.03) | 336.69 (295.94, 382.89) | 314.25 (276.21, 357.35) | 291.90 (256.48, 331.83) | 269.35 (237, 306.30) | 224.46 (197.29, 255,25) | 462.82 (406.8, 526.3) | 367.07 (322.64, 417.42) | 95.74 (84.15, 108.87) |
| Andhra Pradesh | 362.12 (323.31,402.7) | 264.53 (236.18, 294.18) | 188.95 (168.70, 210.12) | 176.35 (157.45, 196.12) | 163.76 (146.21, 182.11) | 151.16 (135, 168.10) | 125.90 (112.47, 140.08) | 259.73 (231.9, 288.84) | 206.1 (183.92, 229.08) | 53.72 (47.97, 59.73) |
| Karnataka | 353.03 (312.43,397.63) | 229.04 (202.46, 258.10) | 163.60 (144.61, 184.36) | 152.70 (133, 172.07) | 141.80 (125.33, 159.80) | 130.34 (116.10, 147.48) | 109.06 (96.41, 122.90) | 224.88 (198.79, 253.42) | 178.36 (157.66, 200.97) | 46.50 (41.21, 52.42) |
| Goa | 3.81 (3.34,4.37) | 7.20 (6.30, 8.25) | 5.14 (4.50, 5.90) | 4.80 (4.20, 5.50) | 4.46 (3.90, 5.10) | 4.11 (3.60, 4.71) | 3.43 (3.01, 3.92) | 7.07 (6.19, 8.1) | 5.61 (4.90, 6.42) | 1.46 (1.28, 1.67) |
| Kerala | 222.98 (192.10,257.66) | 160.32 (139.11, 185.24) | 114.51 (98.64, 132.31) | 106.90 (92.07, 123.49) | 99.24 (85.49, 114.70) | 91.61 (78.91, 105.90) | 76.34 (65.76, 88.20) | 157.41 (135.60, 181.88) | 124.84 (107.55, 144.25) | 32.51 (28.05, 37.62) |
| Tamil Nadu | 355.94 (310.30,407.78) | 327.50 (285.49, 375.20) | 233.92 (203.92, 268) | 218.33 (190.33, 250.13) | 202.73 (176.73, 232.27) | 187.14 (163.13, 214.40) | 155.95 (135.95, 178.67) | 321.56 (280.31, 368.39) | 255.03 (222.30, 292.12) | 66.51 (58, 76.20) |
| Telangana | 237.10 (209.26,270.04) | 159 (140.33, 181.91) | 113.57 (100.20, 129.35) | 106.01 (93.60, 120.72) | 98.42 (86.90, 112.10) | 90.8 (80.10, 103.48) | 75.71 (66.82, 86.23) | 156.11 (137.79, 177.8) | 123.81 (109.28, 141.02) | 32.29 (28.50, 37) |

| | | | | | | | | | | |
|---|---|---|---|---|---|---|---|---|---|---|
| India | 4898.62 (4320.61,5567.40) | 4538.78 (4003.22, 5158.41) | 3241.96 (2859.42, 3684.56) | 3025.84 (2668.81, 3438.42) | 2809.70 (2478.17, 3193.32) | 2593.56 (2287.53, 2947.64) | 2161.31 (1906.29, 2456.38) | 4456.33 (3930.51, 5064.72) | 3534.70 (3117.42, 4017.01) | 921.90 (813.07, 104.71) |

**Supplementary Table 5: Enhanced Cataract Coverage Costs for Meeting Unmet Needs for Cataract Surgery.** All costs are adjusted to millions of USD for the year 2020. NSS - National Sample Survey, PMJAY - Pradhan Mantri Jan Arogya Yojana, PHACO -Phacoemulsification, IOL-Intraocular lens, SICS -Small Incision Cataract Surgery.

| States | Mean Cost, NSS | PHACO with hydrophobic acrylic IOL + Glaucoma, PMJAY | PHACO with hydrophobic acrylic IOL, PMJAY | PHACO with foldable IOL Unilateral, PMJAY | SICS with non-foldable IOL + Glaucoma, PMJAY | SICS with IOL Unilateral, PMJAY | SICS with non-foldable IOL, PMJAY | Societal cost using mean IOL cost, Aravind Eye Hospital | Medical (direct) cost using mean IOL cost, Aravind Eye Hospital | Patient (indirect), Aravind Eye Hospital |
|---|---|---|---|---|---|---|---|---|---|---|
| Jammu & Kashmir and Ladakh | 33.17 (29.01, 38.01) | 36.11 (31.36, 41.47) | 25.80 (22.40, 29.62) | 24.08 (20.90, 27,64) | 22.36 (19.41, 25.67) | 24.08 (20.90, 27.64) | 22.37 (19.41, 25.67 ) | 34.47(30.80, 40.71) | 28.18 (24.42, 32.29) | 7.33 (6.37, 8.42) |
| Himachal Pradesh | 16.08 (13.48, 18.94) | 23.28 (19.50, 27.43) | 16.62 (13.93, 19.60) | 15.52 (13.01, 18.28) | 14.41 (12.07, 16.98) | 15.52 (13.01, 18.28) | 14.41 (12.07, 16.98) | 22.85 (19.15, 26.93) | 18.12 (15.19, 21.32) | 4.72 (3.96, 5.57) |
| Punjab | 101.65 (80.33, 126.12) | 67.84 (53.61, 84.16) | 48.45 (38.29, 60.11) | 45.22 (35.74, 56.11) | 41.99 (33.18, 52.10) | 45.22 (35.74, 56.11) | 42.10 (33.18, 52.10) | 66.60 (52.63, 82.63) | 52.83 (41.74, 65.54) | 13.77 (10.88, 17.09) |
| Uttarakhand | 63.75 (54.09, 75.10) | 31.04 (26.33, 36.51) | 22.17 (18.81, 26.08) | 20.69 (17.55, 24.34) | 19.21 (16.30, 22.60) | 20.69 (17.55, 24.34) | 19.21 (16.30, 22.60) | 30.47 (25.86, 35.85) | 24.17 (20.05, 28.43) | 6.30 (5.34, 7.41) |
| Haryana | 70.57 (59.57, | 74.35 (62.76, | 53.18 (44.83, 62.21) | 49.56 (41.84, | 46.02 (38.85, 53.90) | 49.56 (41.84, | 46.02 (38.85, 53.90) | 73.10 (61.62, 85,49) | 57.90 (48.87, | 15.10 (12.47, 17.68) |

| | | | | | | | | | | |
|---|---|---|---|---|---|---|---|---|---|---|
| | 82.65) | 87.07) | | 58.05) | | 58.05) | | | 67.81) | |
| NCT of Delhi | 13.13 (11.16, 15.3) | 175.80 (148.50,206 .10 ) | 39.82 (33.83, 46.37) | 37.16 (31.5, 43.20) | 34.51 (29.32, 40.19) | 37.16 (31.50, 43.28) | 34.51 (29.23, 40.19) | 54.73 (46.51, 63.74) | 43.41 (36.90, 50.56) | 11.32 (9.62, 13.18) |
| Rajasthan | 302.46 (255.50, 354.11) | 557.48 (473.75, 649.03) | 125.60 (106.11, 147. 07) | 117.24 (99.03, 137.26) | 108.87 (91.96, 127.46) | 117.24 (99.03, 137.26) | 34.90 (29.93, 40.95) | 172.67 (145.86, 202.15) | 136.95 (115.68, 160.34) | 35.72 (30.17, 41.18) |
| Uttar Pradesh | 502.71 (431.03, 589.74) | 563.90 (483.50, 661.52) | 402.78 (345.35, 47.25) | 375.94 (322.33, 441.02) | 349.90 (299.30, 409.50) | 375.93 (322.22, 441.03) | 349.08 (299.30, 409.51) | 553.65 (474.17, 649.51) | 439.12 (376.51, 515.15) | 114.53 (98.20, 134.36) |
| Bihar | 235.93 (200.97, 275.32) | 285.50 (243.46, 333.50) | 204.14 (173.90, 238.23) | 190.50 (162.31, 222.36) | 176.92 (150.71, 206.47) | 190.53 (162.31, 222.23) | 176.92 (150.71, 206.47) | 280.61 (239.04, 327.47) | 222.56 (189.01, 259.73) | 58.04 (49.44, 67.77) |
| Sikkim | 2.16 (1.90, 2.47) | 1.85 (1.62, 2.21) | 1.32 (1.15, 1.52) | 1.23 (1.08, 1.41) | 1.15 (1.05, 1.21) | 1.23 (1.08, 1.41) | 1.15 (1.05, 1.31) | 1.82 (1.59, 2.09) | 1.44 (1.26, 1.65) | 0.37 (0.32, 0.43) |
| Arunachal Pradesh | 3.95 (3.41, 4.57) | 2.80 (2.42, 3.24) | 2.01 (1.73, 2.31) | 1.87 (1.61, 2.16) | 1.73 (1.50, 1.31) | 1.87 (1.61, 2.16) | 1.73 (1.50, 2.09) | 2.57 (2.38, 3.18) | 2.18 (1.88, 2.52) | 0.57 (0.49, 0.65) |
| Nagaland | 3.45 (3.03, 3.92) | 4.40 (3.86, 5.01) | 3.14 (2.75, 3.57) | 2.93 (2.57, 3.33) | 2.72 (2.39, 3.09) | 2.93 (2.57, 3.33) | 2.72 (2.39, 3.09) | 4.32 (3.80, 4.91) | 3.43 (3.08, 3.89) | 0.89 (0.78, 1.01) |
| Manipur | 27.32 (23.85, 31.14) | 9.36 (8.17, 10.70) | 6.68 (5.83, 7.68) | 6.24 (5.44, 7.17) | 5.79 (5.06, 6.66) | 6.24 (5.44, 7.17) | 5.79 (5.06, 6.66) | 9.19 (8.02, 1.05) | 7.29 (6.36,8.38) | 1.90 (1.66, 2.18) |

| | | | | | | | | | | |
|---|---|---|---|---|---|---|---|---|---|---|
| Mizoram | 0 | 2.80 (2.46, 3.35) | 2.05 (1.76, 2.39) | 1.92 (1.64, 2.23) | 1.78 (1.52, 2.07) | 1.92 (1.64, 2.23) | 1.78 (1.52, 2.07) | 2.82 (2.42, 3.29) | 2.24 (1.92, 2.61) | 0.58 (0.50, 0.68) |
| Tripura | 3.10 (2.68, 3.57) | 11.7 (10.19, 13.57) | 8.39 (7.27, 9.68) | 7.83 (6.79, 9.04) | 7.27 (6.30, 8.39) | 7.83 (6.79, 9.04) | 7.27 (6.30, 8.39) | 11.5 (10.10, 13.22) | 9.15 (7.92, 10.50) | 2.38 (2.06, 2.75) |
| Meghalaya | 32.75 (28.52, 37.57) | 6.55 (5.70, 7.51) | 4.67 (4.07, 5.36) | 4.36 (3.80, 5.01) | 4.05 (3.53, 4.65) | 4.36 (3.80, 5.02) | 4.05 (3.52, 4.65) | 6.43 (5.60, 7.37) | 5.10 (4.44, 5.85) | 1.33 (1/25, 1.52) |
| Assam | 52.51 (46.36, 61.63) | 82.51 (71.48, 95.03) | 58.93 (51.06, 67.88) | 55.08 (47.65, 63.35) | 51.07 (44.25, 58.83) | 55.08 (47.65, 63.35) | 51.07 (44.25, 58.83) | 81.01 (70.18, 93.33) | 64.25 (55.66, 74.01) | 16.74 (14.53, 19.30) |
| West Bengal | 233.80 (198.72, 273.34) | 252.02 (214.26, 294.70) | 180.01 (153.03, 210.53) | 168.01 (142.83, 196.47) | 156.07 (132.63, 182.43) | 168.01 (142.83, 196.47) | 156.01 (132.63, 182.43) | 247.45 (210.35, 289.35) | 196.26 (166.84, 229.49) | 51.18 (43.51, 60.10) |
| Jharkhand | 137.80 (118.83, 159.36) | 100.94 (87.05, 116.74) | 72.10 (62.18, 83.39) | 67.29 (58.03, 77.83) | 62.49 (54.10, 72.27) | 67.29 (58.03, 77.83) | 62.49 (54.10, 72.27) | 99.11 (85.47, 114.60) | 78.61 (67.79, 90.91) | 20.50 (17.68, 23.71) |
| Odisha | 117.60 (99.94, 136.62) | 137.70 (117.02, 159.97) | 98.36 (83.59, 114.26) | 91.83 (78.01, 016.68) | 85.24 (72.44, 99.03) | 91.83 (78.01, 106.64) | 85.24 (72.44, 99.03) | 135.20 (114.90, 157.06) | 107.23 (91.18, 124.60) | 27.96 (23.76, 32.49) |
| Chhattisgarh | 85.24 (73.41, 97.80) | 100.86 (86.86, 115.71) | 72.04 (62.04, 82.65) | 67.24 (57.90, 77.14) | 62.43 (54.10, 71.63) | 67.24 (57.90, 77.14) | 62.43 (53.77, 71.66) | 99.03 (85.28, 113.61) | 78.54 (67.64, 90.17) | 20.48 (17.64, 23.50) |
| Madhya Pradesh | 74.84 (57.25, | 114.35 (87.47, | 81.68 (62.48, 102.39) | 76.23 (58.31, | 70.79 (54.15, 88.74) | 76.23 (58.31, | 70.80 (54.15, 88.74) | 112.27 (85.88, 140.74) | 89.05 (68.12, | 23.33 (17.67, 29.11) |

| | | | | | | | | | | |
|---|---|---|---|---|---|---|---|---|---|---|
| | 93.81) | 143.35) | | 95.56) | | 95.56) | | | 111.63) | |
| Gujarat | 77.05 (57.43, 93.81) | 104.95 (78.22, 13.51) | 74.96 (55.87, 96.54) | 68.96 (52.14, 90.14) | 64.97 (48.42, 83.66) | 70.10 (52.14,90.10) | 64.91 (48.42, 84.10) | 103.04 (76.80, 132.70) | 81.72 (60.91, 105.25) | 21.31 (15.88, 27.45) |
| Maharashtra | 516.22 (435.10, 608.39) | 362.48 (305.42, 427.89) | 258.91 (218.16, 305.10) | 24.16 (20.36, 28.47) | 224.3 (189.07, 264.42) | 241.65 (203.61, 285.10) | 224.39 (189.07, 264.42) | 355.90 (299.90, 419.38) | 282.27 (237.83, 332.62) | 73.62 (62.03, 86.75) |
| Andhra Pradesh | 287.19 (248.38, 327.77) | 209.80 (181.45, 239.44) | 149.85 (129.60, 171.80) | 139.86 (120.96, 159.63) | 129.87 (112.32, 142.22) | 139.86 (120.96, 159.62) | 129.90 (112.32, 148.22) | 206.10 (178.15, 235.09) | 163.38 (141.30, 186.46) | 42.61 (36.85, 48.63) |
| Karnataka | 262.28 (221.89, 307.62) | 170.61 (144.03, 199.67) | 121.85 (101.89, 142.62) | 113.74 (96.02, 133.11) | 105.61 (89.16, 123.61) | 113.74 (96.22, 133.41) | 105.61 (89.63, 123.61) | 167.51 (141.41, 196.05) | 132.86 (112.16, 155.49) | 34.65 (29.25, 40.55) |
| Goa | 2.91 (2.43, 3.46) | 5.49 (4.59, 6.53) | 3.92 (3.27, 4.67) | 3.66 (3.06, 4.35) | 3.40 (2.84, 4.04) | 3.66 (3.06, 4.35) | 3.40 (2.84, 4.04) | 5.39 (4.50, 6.41) | 4.27 (3.57,5.09) | 1.11 (0.93, 1.32) |
| Kerala | 186.62 (155.72, 221.30) | 134.16 (111.19, 159.30 ) | 95.83 (79.96, 113.63) | 89.44 (74.63, 106.05) | 83.05 (69.30, 98.48) | 89.44 (74.63, 106.05) | 83.05 (69.30, 98.48) | 131.73 (109.92, 156.19) | 104.48 (87.18, 123.88) | 27.25 (22.73, 32.31) |
| Tamil Nadu | 301.80 (256.12, 353.62) | 277.66 (235.65, 326.54) | 198.33 (168.32, 232.40) | 185.10 (157.10, 216.91) | 171.88 (145.90, 201.41) | 185.10 (157.10, 216.91) | 171.89 (145.62, 210.41) | 272.62 (231.37, 319.45) | 216.22 (183.51, 253.37) | 56.39 (47.86, 66.08) |
| Telangana | 184.39 (156.56, 217.33) | 123.65 (104.99, 145.74) | 88.32 (75.10, 104.10) | 82.4 (69.60, 97.16 ) | 76.54 (65.10, 90.22) | 82.43 (69.99, 97.16) | 76.54 (65.10, 90.22) | 121.41 (103.08, 143.10) | 96.29 (82.10, 113.49) | 25.11 (21.32, 29.60) |

| | | | | | | | | | | |
|---|---|---|---|---|---|---|---|---|---|---|
| India | 3823.70 (3245.69, 4492.47) | 3542.80 (3007.25, 4162.45) | 2530.56 (2148.02, 2973.16) | 2361.90 (2004.83, 2778.10) | 219.31 (186.16, 257.67) | 2361.87 (2004.83, 2774.96) | 2193.61 (1861.63, 2576.75) | 3478.45 (2952.63, 4086.84) | 2760.10 (2341.84, 3241.42) | 719.56 (610.80, 845.20) |

**Supplementary Table 6: Complication Rate-Adjusted Cataract Surgeries and Unmet Needs.**
NHP - National Health Profile, NSS - National Sample Survey, PHACO -Phacoemulsification, IOL-Intraocular lens, SICS -Small Incision Cataract Surgery.

| States | Complication rate-adjusted surgeries achieved total | Complication rate-adjusted surgeries achieved PHACO | Complication rate-adjusted surgeries achieved SICS | Complication rate-adjusted unmet need total | Complication rate-adjusted unmet need PHACO | Complication rate-adjusted unmet need SICS |
|---|---|---|---|---|---|---|
| Jammu & Kashmir and Ladakh | 25543.83 | 25554.16 | 25580.01 | 241860.37 (209985.67, 277558.77) | 241850.04 (209975.34, 277548.44) | 241824.19 (209949.49, 277522.59) |
| Himachal Pradesh | 31720.97 | 31733.80 | 31765.89 | 156008.44 (130801.54, 183769.54) | 155995.60 (130788.70, 183756.70) | 155963.51 (130756.61, 183724.61) |
| Punjab | 239406.79 | 239503.67 | 239745.86 | 456342.81 (361211.11, 565495.21) | 456245.93 (361114.23, 565398.33) | 456003.74 (360872.04, 565156.14) |
| Uttarakhand | 50050.72 | 50070.97 | 50121.61 | 208107.18 (176669.68, 244692.78) | 208086.93 (176649.43, 244672.53) | 208036.29 (176598.79, 244621.89) |
| Haryana | 139532.71 | 139589.17 | 139730.32 | 498705.99 (421239.39, 583788.39) | 498649.53 (421182.93, 583731.93) | 498508.38 (421041.78, 583590.78) |
| NCT of Delhi | 82217.50 | 82250.77 | 82333.94 | 373665.20 (317687.50, 435037.90) | 373631.93 (317654.23, 435004.63) | 373548.76 (317571.06, 434921.46) |
| Rajasthan | 271966.99 | 272077.05 | 272352.18 | 1178940.01 996359.01, 1379710.01) | 1178829.95 (996248.95, 1379599.95) | 1178554.82 (995973.82, 1379324.82) |
| Uttar Pradesh | 976398.78 | 976793.89 | 977781.64 | 3781344.22 (3243809.22, 4434044.22) | 3780949.11 (3243414.11, 4433649.11) | 3779961.36 (3242426.36, 4432661.36) |

| | | | | | | |
|---|---|---|---|---|---|---|
| Bihar | 393051.32 | 393210.37 | 393608.00 | 1915331.68 (1632284.68, 2234409.68) | 1915172.63 (1632125.63, 2234250.63) | 1914775.00 (1631728.00, 2233853.00) |
| Sikkim | 596.07 | 596.31 | 596.91 | 12432.30 (10861.57, 14241.76) | 12432.06 (10861.33, 14241.52) | 12431.46 (10860.73, 14240.92) |
| Arunachal Pradesh | 2315.07 | 2316.00 | 2318.35 | 18801.06 (16247.41, 21728.39) | 18800.13 (16246.48, 21727.46) | 18797.78 (16244.13, 21725.11) |
| Nagaland | 1298.89 | 1299.41 | 1300.73 | 29484.82 (25841.04, 33471.83) | 29484.30 25840.52, 33471.31) | 29482.98 (25839.20, 33469.99) |
| Manipur | 2793.50 | 2794.63 | 2797.46 | 62625.81 (54681.08, 72002.71) | 62624.68 (54679.95, 72001.58) | 62621.85 (54677.12, 71998.75) |
| Mizoram | 2973.41 | 2974.61 | 2977.62 | 19291.22 (16546.46, 22483.24) | 19290.02 (16545.26, 22482.04) | 19287.01 (16542.25, 22479.03) |
| Tripura | 9424.36 | 9428.17 | 9437.71 | 78714.68 (68239.23, 90819.64) | 78710.87 (68235.42, 90815.83) | 78701.33 (68225.88, 90806.29) |
| Meghalaya | 3051.50 | 3052.73 | 3055.82 | 43831.46 (38180.48, 50279.18) | 43830.23 (38179.25, 50277.95) | 43827.14 (38176.16, 50274.86) |
| Assam | 65628.49 | 65655.05 | 65721.44 | 552401.01 (478690.91, 636123.21) | 552374.45 (478664.35, 636096.65) | 552308.06 (478597.96, 636030.26) |
| West Bengal | 310086.52 | 310212.00 | 310525.69 | 1688554.48 (1435989.48, 1973881.48) | 1688429.00 (1435864.00, 1973756.00) | 1688115.31 (1435550.31, 1973442.31) |
| Jharkhand | 90198.65 | 90235.15 | 90326.40 | 675945.95 (583041.85, 781564.65) | 675909.45 (583005.35, 781528.15) | 675818.20 (582914.10, 781436.90) |
| Odisha | 147268.71 | 147328.30 | 147477.28 | 922345.29 (784112.89, 1071220.29) | 922285.70 (784053.30, 1071160.70) | 922136.72 (783904.32, 1071011.72) |

| | | | | | | |
|---|---|---|---|---|---|---|
| Chhattisgarh | 117241.04 | 117288.48 | 117407.09 | 675681.26 (582095.06, 774966.96) | 675633.82 (582047.62, 774919.52) | 675515.21 (581929.01, 774800.91) |
| Madhya Pradesh | 611090.70 | 611337.98 | 611956.18 | 771626.30 (591936.30, 965502.30) | 771379.02 (591689.02, 965255.02) | 770760.82 (591070.82, 964636.82) |
| Gujarat | 793488.72 | 793809.81 | 794612.53 | 710892.28 (532188.28, 912827.28) | 710571.19 (531867.19, 912506.19) | 709768.47 (531064.47, 911703.47) |
| Maharashtra | 719644.80 | 719936.01 | 720664.03 | 2431778.20 (2050314.20, 2864047.20) | 2431486.99 (2050022.99, 2863755.99) | 2430758.97 (2049294.97, 2863027.97) |
| Andhra Pradesh | 361725.76 | 361872.13 | 362238.07 | 1406855.24 (1217320.24, 1605035.24) | 1406708.87 (1217173.87, 1604888.87) | 1406342.93 (1216807.93, 1604522.93) |
| Karnataka | 386137.76 | 386294.01 | 386684.64 | 1145142.25 (967429.25, 1339456.25) | 1144985.99 (967272.99, 1339299.99) | 1144595.36 (966882.36, 1338909.36) |
| Goa | 11316.35 | 11320.93 | 11332.38 | 36854.44 (30822.20, 43844.22) | 36849.86 (30817.62, 43839.64) | 36838.41 (30806.17, 43828.19) |
| Kerala | 172838.24 | 172908.18 | 173083.03 | 899006.76 (750503.36, 1065587.76) | 898936.82 (750433.42, 1065517.82) | 898761.97 (750258.57, 1065342.97) |
| Tamil Nadu | 329371.17 | 329504.45 | 329837.65 | 1860161.83 (1579316.83, 2179089.83) | 1860028.55 (1579183.55, 2178956.55) | 1859695.35 (1578850.35, 2178623.35) |
| Telangana | 233572.67 | 233667.18 | 233903.47 | 829428.34 (704640.34, 977116.34) | 829333.82 (704545.82, 977021.82) | 829097.53 (704309.53, 976785.53) |
| India | 6581951.96 | 6584615.37 | 6591273.90 | 23762092.04 (20181664.04, 27904762.04) | 23759428.63 (20179000.63, 27902098.63) | 23752770.10 (20172342.10, 27895440.10) |

**Supplementary Table 7: Complication Rate-Adjusted Enhanced Coverage Costs for Meeting Unmet Needs for Cataract Surgery.** All costs are adjusted to millions of USD for the year 2020. NSS - National Sample Survey, PMJAY - Pradhan Mantri Jan Arogya Yojana, PHACO -Phacoemulsification, IOL-Intraocular lens, SICS -Small Incision Cataract Surgery.

| States | Mean Cost, NSS | Societal cost using mean IOL cost, Aravind Eye Hospital | Medical (direct) cost using mean IOL cost, Aravind Eye Hospital | Patient (indirect) cost, Aravind Eye Hospital | PHACO with hydrophobic acrylic IOL + Glaucoma, PMJAY | PHACO with hydrophobic acrylic IOL, PMJAY | PHACO with foldable IOL Unilateral, PMJAY | SICS with non-foldable IOL + Glaucoma, PMJAY | SICS with IOL Unilateral, PMJAY | SICS with non-foldable IOL, PMJAY |
|---|---|---|---|---|---|---|---|---|---|---|
| Jammu & Kashmir and Ladakh | 39.00 (33.9,44.8) | 35.50 (30.80, 40.80) | 28.20 (24.50, 32.30) | 7.33 (6.40, 8.42) | 36.20 (31.40, 41.50) | 25.80 (22.40, 29.70) | 24.10 (20.90, 27.70) | 22.40 (19.41, 25.70) | 20.70 (17.90, 23.70) | 17.20 (15.0, 19.80) |
| Himachal Pradesh | 25.20 (21.10, 29.70) | 22.90 (19.20, 27.00) | 18.20 (15.20, 21.40) | 4.72 (4.00, 5.60) | 23.30 (19.60, 27.50) | 16.70 (14.00, 19.60) | 15.60 (13.01, 18.30) | 14.41 (12.10, 17.00) | 13.30 (11.20, 15.70) | 11.10 (9.30, 13.10) |
| Punjab | 73.70 (58.30, 91.30) | 67.00 (53.00, 83.00) | 53.20 (42.10, 65.90) | 13.90 (11.00, 17.20) | 68.20 (54.00, 84.60) | 48.70 (38.60, 60.40) | 45.50 (36.00, 56.40) | 42.20 (33.44, 52.30) | 39.00 (30.80, 48.30) | 32.50 (25.70, 40.30) |
| Uttarakhand | 33.60 (28.50, 39.50) | 30.60 (25.90, 35.90) | 24.20 (20.60, 28.50) | 6.30 (5.40, 7.41) | 31.10 (26.40, 36.60) | 22.20 (18.90, 26.10) | 20.70 (17.60, 24.40) | 19.30 (16.40, 22.70) | 17.80 (15.1, 20.90) | 14.80 (12.60, 17.40) |
| Haryana | 80.50 (68.00, 94.20) | 73.20 (61.90, 85.70) | 58.10 (49.10, 68.00) | 15.20 (12.80, 17.70) | 74.60 (63.00, 87.30) | 53.30 (45.00, 62.40) | 49.70 (42.00, 58.20) | 46.20 (39.00, 54.00) | 42.60 (36.00, 49.90) | 35.50 (30.00, 41.60) |

| | | | | | | | | | | |
|---|---|---|---|---|---|---|---|---|---|---|
| NCT of Delhi | 60.30 (51.30, 70.20) | 54.90 (46.70, 63.90) | 43.50 (37.00, 50.70) | 11.40 (9.70, 13.20) | 55.90 (47.50, 65.10) | 39.90 (33.90, 46.50) | 37.30 (31.70, 43.40) | 34.60 (29.40, 40.30) | 31.90 (27.10, 37.20) | 26.60 (22.60, 31.00) |
| Rajasthan | 190.30 (160.80, 222.70) | 173.10 (146.30, 202.60) | 137.30 (116.10, 160.70) | 35.80 (30.30, 41.90) | 176.30 (149.00, 206.40) | 125.90 (106.40, 147. 40) | 117.60 (99.30, 137.60) | 109.10 (92.20, 127.70) | 100.70 (85.10, 117.90) | 83.90 (70.90, 98.20) |
| Uttar Pradesh | 610.40 (523.70, 715.80) | 555.30 (476.40, 651.20) | 440.50 (377.80, 516.50) | 114.90 (98.50, 134.70) | 565.50 (485.10, 663.20) | 404.00 (346.50, 473.70) | 377.00 (323.40, 442.10) | 350.00 (300.20, 410.40) | 323.10 (277.10, 378.90) | 269.20 (230.90, 315.70) |
| Bihar | 309.20 (263.50, 360.70) | 281.30 (239.70, 328.10) | 223.10 (190.10, 260.30) | 58.20 (49.60, 67.90) | 286.50 (244.10, 334.20) | 204.60 (174.40, 238.70) | 191.00 (162.80, 222.80) | 177.30 (151.10, 206.80) | 163.70 (139.50, 190.90) | 136.40 (116.20, 159.10) |
| Sikkim | 2.00 (1.80, 2.30) | 1.80 (1.60, 2.10) | 1.40 (1.30, 1.70) | 0.40 (0.32, 0.43) | 1.90 (1.62, 2.10) | 1.30 (1.20, 1.52) | 1.20 (1.10, 1.41) | 1.20 (1.05, 1.30) | 1.10 (0.90, 1.20) | 0.90 (0.80, 1.00) |
| Arunachal Pradesh | 3.00 (2.60, 3.50) | 2.80 (2.40, 3.20) | 2.20 (1.90, 2.52) | 0.60 (0.50, 0.70) | 2.80 (2.42, 3.24) | 2.00 (1.73, 2.31) | 1.90 (1.61, 2.20) | 1.70 (1.50, 2.00) | 1.60 (1.41, 1.90) | 1.30 (1.20, 1.50) |
| Nagaland | 4.80 (4.20, 5.40) | 4.32 (3.80, 4.91) | 3.43 (3.08, 3.90) | 0.90 (0.80, 1.01) | 4.40 (3.90, 5.01) | 3.20 (2.80, 3.60) | 2.93 (2.60, 3.33) | 2.72 (2.40, 3.10) | 2.50 (2.20, 2.90) | 2.10 (1.80, 2.40) |
| Manipur | 10.10 (8.80, 11.60) | 9.20 (8.02, 10.60) | 7.30 (6.40,8.40) | 1.90 (1.70, 2.20) | 9.40 (8.20, 10.80) | 6.70 (5.83, 7.70) | 6.24 (5.50, 7.20) | 5.80 (5.10, 6.70) | 5.40 (4.70, 6.20) | 4.50 (3.90, 5.10) |
| Mizoram | 3.10 (2.70, | 2.82 (2.42, | 2.24 (1.92, | 0.60 | 2.90 (2.50, | 2.10 (1.80, | 1.92 (1.64, | 1.80 (1.50, | 1.60 (1.40, | 1.40 (1.20, 1.60) |

| | | | | | | | | | | |
|---|---|---|---|---|---|---|---|---|---|---|
| | 3.60) | 3.30) | 2.61) | (0.50, 0.70) | 3.40) | 2.39) | 2.20) | 2.10) | 1.90) | |
| Tripura | 12.70 (11.00, 14.70) | 11.60 (10.00, 13.30) | 9.20 (7.92, 10.60) | 2.40 (2.10, 2.80) | 11.80 (10.20, 13.60) | 8.40 (7.30, 9.70) | 7.83 (6.80, 9.10) | 7.30 (6.30, 8.40) | 6.70 (5.80, 7.80) | 5.60 (4.90, 6.50) |
| Meghalaya | 7.10 (6.20, 8.10) | 6.43 (5.60, 7.40) | 5.10 (4.44, 5.90) | 1.33 (1.25, 1.52) | 6.60 (5.70, 7.51) | 4.70 (4.10, 5.40) | 4.40 (3.80, 5.01) | 4.10 (3.53, 4.70) | 3.70 (3.30, 4.30) | 3.10 (2.70, 3.60) |
| Assam | 89.20 (77.30, 102.70) | 81.10 (70.30, 93.40) | 64.30 (55.80, 74.10) | 16.80 (14.53, 19.30) | 82.60 (71.60, 95.10) | 59.00 (51.10, 68.00) | 55.10 (47.70, 63.40) | 51.10 (44.30, 58.90) | 47.20 (40.90, 54.40) | 39.30 (34.10, 45.30) |
| West Bengal | 272.60 (231.80, 318.70) | 248.00 (210.90, 289.90) | 196.70 (167.30, 229.90) | 51.30 (43.60, 60.00) | 252.60 (214.80, 295.20) | 180.40 (153.40, 210.90) | 168.40 (143.20, 196.80) | 156.30 (132.90, 182.70) | 144.30 (122.70, 168.70) | 120.20 (102.20, 140.60) |
| Jharkhand | 109.10 (94.10, 126.20) | 99.30 (85.60, 114.80) | 78.70 (67.90, 91.00) | 20.50 (17.70, 23.71) | 101.00 (87.20, 116.90) | 72.20 (62.30, 83.50) | 67.40 (58.10, 77.90) | 62.60 (54.00, 72.40) | 57.80 (49.80, 66.80) | 48.10 (41.50, 55.70) |
| Odisha | 148.90 (126.60, 172.90) | 135.50 (115.20, 157.30) | 107.40 (91.30, 124.80) | 28.00 (23.80, 32.50) | 138.00 (117.30, 160.20) | 98.50 (83.80, 114.40) | 92.00 (78.20, 106.80) | 85.40 (72.60, 99.20) | 78.80 (67.00, 91.50) | 65.70 (55.80, 76.30) |
| Chhattisgarh | 109.10 (94.00, 125.10) | 99.20 (85.50, 113.80) | 78.70 (67.80, 90.30) | 20.50 (17.70, 23.50) | 101.10 (87.10, 115.90) | 72.20 (62.20, 82.80) | 67.40 (58.00, 77.30) | 62.50 (53.90, 71.70) | 57.70 (49.70, 66.20) | 48.10 (41.40, 55.20) |
| Madhya Pradesh | 124.60 (95.60, | 113.30 (86.90, | 89.90 (68.90, | 23.40 (18.00, | 115.40 (88.50, | 82.40 (63.20, | 76.90 (59.00, | 71.40 (54.70, | 65.90 (50.50, | 54.90 (42.10, 68.74) |

| | | | | | | | | | | |
|---|---|---|---|---|---|---|---|---|---|---|
| | 155.90) | 141.80) | 112.50) | 29.30) | 144.40) | 103.10) | 96.30) | 89.30) | 82.40) | |
| Gujarat | 114.80 (85.90, 147.40) | 104.40 (78.20, 134.10) | 82.80 (62.00, 106.30) | 21.60 (16.20, 27.70) | 106.30 (79.60, 136.50) | 75.90 (56.80, 97.50) | 70.90 (53.00, 91.00) | 65.70 (49.20, 84.40) | 60.70 (45.40, 77.90) | 50.60 (37.80, 64.90) |
| Maharashtra | 392.60 (331.00, 462.40) | 357.10 (301.10, 420.60) | 283.30 (238.80, 333.60) | 73.90 (62.30, 87.00) | 363.70 (306.60, 428.40) | 259.80 (219.00, 306.00) | 242.50 (204.40, 285.60) | 225.10 (189.90, 265.10) | 207.80 (175.20, 244.70) | 173.10 (146.00, 203.90) |
| Andhra Pradesh | 227.10 (196.50, 259.10) | 206.60 (178.80, 235.70) | 163.90 (141.80, 187.00) | 42.70 (37.00, 48.80) | 210.40 (182.10, 240.10) | 150.30 (130.00, 171.50) | 140.30 (121.40, 160.00) | 130.20 (112.70, 148.60) | 120.20 (104.00, 137.10) | 100.20 (86.70, 114.30) |
| Karnataka | 184.90 (156.20, 216.20) | 168.20 (142.10, 196.70) | 133.40 (112.70, 156.00) | 34.80 (29.40, 40.70) | 171.30 (144.70, 200.30) | 122.30 (103.30, 143.10) | 114.20 (96.50, 133.60) | 106.00 (89.50, 124.00) | 97.80 (82.60, 114.40) | 81.50 (68.90, 95.40) |
| Goa | 5.90 (5.00, 7.10) | 5.40 (4.50, 6.41) | 4.30 (3.60,5.10) | 1.11 (0.93, 1.32) | 5.50 (4.60, 6.60) | 3.92 (3.30, 4.70) | 3.70 (3.10, 4.40) | 3.40 (2.90, 4.10) | 3.10 (2.60, 3.70) | 2.60 (2.20, 3.10) |
| Kerala | 145.20 (121.30, 172.10) | 132.10 (110.30, 156.60) | 104.80 (87.50, 124.20) | 27.30 (22.80, 32.40) | 134.50 (111.20, 159.40) | 96.00 (80.20, 113.80) | 89.60 (74.80, 106.30) | 83.20 (69.50, 98.60) | 76.80 (64.10, 91.10) | 64.00 (53.40, 75.90) |
| Tamil Nadu | 300.30 (255.00, 351.80) | 273.20 (231.90, 320.00) | 216.70 (184.00, 253.80) | 56.50 (48.00, 66.20) | 278.20 (236.20, 325.90) | 198.70 (168.70, 232.80) | 185.50 (157.50, 217.30) | 172.20 (146.20, 201.70) | 159.00 (134.90, 186.20) | 132.50 (112.50, 155.20) |
| Telangana | 133.90 (113.80, 157.70) | 121.80 (103.50, 143.50) | 96.60 (82.10, 113.80) | 25.20 (21.40, 29.70) | 124.00 (105.40, 146.10) | 88.60 (75.30, 104.4) | 82.70 (70.30, 97.40) | 76.80 (65.20, 90.40) | 70.90 (60.20, 83.50) | 59.10 (50.20, 69.60) |

| | | | | | | | | | | |
|---|---|---|---|---|---|---|---|---|---|---|
| India | 3823.30 (3230.30, 4493.50) | 3478.10 (2938.60, 4087.80) | 2758.60 (2330.70, 3242.20) | 719.56 (607.90, 845.60) | 3541.90 (2992.50, 4162.90) | 2529.90 (2137.50, 2973.50) | 2361.30 (1995.00, 2775.30) | 2192.00 (1851.90, 2576.40) | 2023.40 (1709.40, 2378.20) | 1686.10 (1424.50, 1981.90) |

**Supplementary Table 8: Complication Rate-Adjusted Economic Benefits for Meeting Unmet Needs for Cataract Surgery.** All costs are adjusted to USD for the year 2020. NSS - National Sample Survey, PHACO -Phacoemulsification, SICS -Small Incision Cataract Surgery, DALY -Disability Adjusted Life-Years.

| States | Complication rate-adjusted mean DALY count unmet need total | Complication rate-adjusted PHACO DALY count unmet need | Complication rate-adjusted SICS DALY count unmet need | Complication rate-adjusted value of life-year total | Complication rate-adjusted value of life-year PHACO | Complication rate-adjusted value of life-year SICS |
|---|---|---|---|---|---|---|
| Jammu & Kashmir and Ladakh | 16926.97 | 16926.24 | 16924.43 | 79296412.25 | 79293023.35 | 79284551.11 |
| Himachal Pradesh | 10830.94 | 10830.05 | 10827.82 | 94844365.07 | 94836561.5 | 94817052.59 |
| Punjab | 31664.67 | 31657.95 | 31641.15 | 225939211.2 | 225891246.6 | 225771335.3 |
| Uttarakhand | 14424.82 | 14423.41 | 14419.90 | 130528083 | 130515379.9 | 130483622.1 |
| Haryana | 34850.99 | 34847.04 | 34837.18 | 361838755.8 | 361797789.2 | 361695372.7 |
| NCT of Delhi | 28316.49 | 28313.97 | 28307.66 | 456335736.4 | 456295106.2 | 456193530.5 |
| Rajasthan | 82343.80 | 82336.11 | 82316.89 | 402389475.6 | 402351913.1 | 402258006.9 |
| Uttar Pradesh | 270552.71 | 270524.44 | 270453.76 | 791702337.5 | 791619614.5 | 791412807 |
| Bihar | 132535.14 | 132524.14 | 132496.62 | 241845758.2 | 241825675.2 | 241775467.9 |
| Sikkim | 863.39 | 863.37 | 863.33 | 8953260.018 | 8953086.315 | 8952652.058 |
| Arunachal Pradesh | 1346.17 | 1346.11 | 1345.94 | 13024932.64 | 13024283.65 | 13022661.16 |
| Nagaland | 2090.84 | 2090.80 | 2090.71 | 11240081.82 | 11239881.45 | 11239380.54 |

| | | | | | | |
|---|---|---|---|---|---|---|
| Manipur | 4394.54 | 4394.46 | 4394.27 | 15324798.55 | 15324521.94 | 15323830.41 |
| Mizoram | 1350.61 | 1350.52 | 1350.31 | 10983637.19 | 10982952.14 | 10981239.51 |
| Tripura | 5626.97 | 5626.70 | 5626.02 | 27505625.31 | 27504292.71 | 27500961.2 |
| Meghalaya | 3139.97 | 3139.88 | 3139.66 | 13812605.02 | 13812215.89 | 13811243.09 |
| Assam | 39636.55 | 39634.65 | 39629.88 | 149063112.5 | 149055946.3 | 149038030.7 |
| West Bengal | 126675.82 | 126666.41 | 126642.88 | 587731737.8 | 587688063.1 | 587578876.2 |
| Jharkhand | 49169.84 | 49167.18 | 49160.54 | 163531927.4 | 163523097.2 | 163501021.5 |
| Odisha | 67422.68 | 67418.32 | 67407.43 | 296328241.4 | 296309095.6 | 296261231.1 |
| Chhattisgarh | 49926.52 | 49923.02 | 49914.25 | 224776718.4 | 224760936 | 224721480 |
| Madhya Pradesh | 56410.73 | 56392.65 | 56347.46 | 224785368 | 224713331.9 | 224533241.8 |
| Gujarat | 48483.71 | 48461.81 | 48407.07 | 429473883.1 | 429279903.1 | 428794953 |
| Maharashtra | 165564.97 | 165545.14 | 165495.58 | 1475782250 | 1475605524 | 1475163710 |
| Andhra Pradesh | 98379.90 | 98369.66 | 98344.07 | 669617873.2 | 669548204 | 669374031.2 |
| Karnataka | 88806.26 | 88794.14 | 88763.85 | 829304602.9 | 829191446.2 | 828908554.5 |
| Goa | 2445.50 | 2445.20 | 2444.44 | 38279429.08 | 38274672.82 | 38262782.18 |
| Kerala | 62384.72 | 62379.86 | 62367.73 | 556715986.4 | 556672675.8 | 556564399.4 |
| Tamil Nadu | 123367.01 | 123358.17 | 123336.08 | 1073766297 | 1073689361 | 1073497022 |
| Telangana | 62540.62 | 62533.49 | 62515.67 | 568446186.2 | 568381409.9 | 568219469.3 |
| India | 1687356.68 | 1687167.55 | 1686694.72 | 27252663522 | 27249608866 | 27241972226 |

**Supplementary Table 9: Complication Rate-Adjusted Net Benefits for Meeting Unmet Needs for Cataract Surgery.** All costs are adjusted to millions of USD for the year 2020. NSS - National Sample Survey, PMJAY - Pradhan Mantri Jan Arogya Yojana, PHACO -Phacoemulsification, IOL-Intraocular lens, SICS -Small Incision Cataract Surgery.

| States | Mean Cost, NSS | Societal cost using mean IOL cost, Aravind Eye Hospital | Medical (direct) cost using mean IOL cost, Aravind Eye Hospital | Patient (indirect) cost, Aravind Eye Hospital | PHACO with hydrophobic acrylic IOL + Glaucoma, PMJAY | PHACO with hydrophobic acrylic IOL, PMJAY | PHACO with foldable IOL Unilateral, PMJAY | SICS with non-foldable IOL + Glaucoma, PMJAY | SICS with IOL Unilateral, PMJAY | SICS with non-foldable IOL, PMJAY |
|---|---|---|---|---|---|---|---|---|---|---|
| Jammu & Kashmir and Ladakh | 40.3 | 43.8 | 51.1 | 71.9 | 43.1 | 53.5 | 55.2 | 56.9 | 58.6 | 62.1 |
| Himachal Pradesh | 69.7 | 71.9 | 76.7 | 90.1 | 71.5 | 78.2 | 79.3 | 80.4 | 81.5 | 83.7 |
| Punjab | 152.3 | 158.9 | 172.8 | 212.1 | 157.6 | 177.1 | 180.4 | 183.5 | 186.8 | 193.3 |
| Uttarakhand | 96.9 | 100.0 | 106.3 | 124.2 | 99.4 | 108.3 | 109.8 | 111.2 | 112.7 | 115.7 |
| Haryana | 281.3 | 288.6 | 303.7 | 346.7 | 287.2 | 308.5 | 312.1 | 315.5 | 319.1 | 326.2 |
| NCT of Delhi | 396.0 | 401.5 | 412.8 | 445.0 | 400.4 | 416.4 | 419.0 | 421.6 | 424.3 | 429.6 |
| Rajasthan | 212.1 | 229.3 | 265.1 | 366.6 | 226.0 | 276.4 | 284.8 | 293.1 | 301.5 | 318.3 |
| Uttar Pradesh | 181.3 | 236.4 | 351.3 | 676.8 | 226.1 | 387.7 | 414.6 | 441.4 | 468.3 | 522.2 |
| Bihar | -67.4 | -39.4 | 18.7 | 183.7 | -44.6 | 37.2 | 50.8 | 64.5 | 78.1 | 105.4 |

| | | | | | | | | | | |
|---|---|---|---|---|---|---|---|---|---|---|
| Sikkim | 6.9 | 7.1 | 7.5 | 8.6 | 7.1 | 7.6 | 7.7 | 7.8 | 7.9 | 8.1 |
| Arunachal Pradesh | 10.0 | 10.3 | 10.8 | 12.5 | 10.2 | 11.0 | 11.1 | 11.3 | 11.4 | 11.7 |
| Nagaland | 6.5 | 6.9 | 7.8 | 10.3 | 6.8 | 8.1 | 8.3 | 8.5 | 8.7 | 9.1 |
| Manipur | 5.2 | 6.1 | 8.0 | 13.4 | 6.0 | 8.6 | 9.1 | 9.5 | 10.0 | 10.9 |
| Mizoram | 7.9 | 8.2 | 8.7 | 10.4 | 8.1 | 8.9 | 9.1 | 9.2 | 9.3 | 9.6 |
| Tripura | 14.8 | 15.9 | 18.3 | 25.1 | 15.7 | 19.1 | 19.7 | 20.2 | 20.8 | 21.9 |
| Meghalaya | 6.7 | 7.4 | 8.7 | 12.5 | 7.3 | 9.1 | 9.4 | 9.8 | 10.1 | 10.7 |
| Assam | 59.9 | 67.9 | 84.7 | 132.3 | 66.4 | 90.0 | 94.0 | 97.9 | 101.8 | 109.7 |
| West Bengal | 315.1 | 339.8 | 391.0 | 536.4 | 335.1 | 407.3 | 419.3 | 431.3 | 443.3 | 467.3 |
| Jharkhand | 54.4 | 64.3 | 84.8 | 143.0 | 62.4 | 91.3 | 96.1 | 100.9 | 105.7 | 115.4 |
| Odisha | 147.4 | 160.9 | 188.9 | 268.3 | 158.4 | 197.8 | 204.3 | 210.9 | 217.4 | 230.6 |
| Chhattisgarh | 115.7 | 125.5 | 146.1 | 204.2 | 123.7 | 152.6 | 157.4 | 162.2 | 167.0 | 176.6 |
| Madhya Pradesh | 100.2 | 111.5 | 134.9 | 201.3 | 109.3 | 142.3 | 147.8 | 153.2 | 158.7 | 169.6 |
| Gujarat | 314.7 | 325.1 | 346.7 | 407.9 | 323.0 | 353.4 | 358.4 | 363.1 | 368.1 | 378.2 |
| Maharashtra | 1083.2 | 1118.7 | 1192.5 | 1401.9 | 1111.9 | 1215.8 | 1233.1 | 1250.1 | 1267.4 | 1302.0 |
| Andhra Pradesh | 442.5 | 463.0 | 505.7 | 626.9 | 459.1 | 519.3 | 529.3 | 539.2 | 549.2 | 569.2 |
| Karnataka | 644.4 | 661.1 | 695.9 | 794.5 | 657.9 | 706.9 | 715.0 | 722.9 | 731.1 | 747.4 |
| Goa | 32.3 | 32.9 | 34.0 | 37.2 | 32.8 | 34.3 | 34.6 | 34.9 | 35.1 | 35.6 |
| Kerala | 411.6 | 424.7 | 452.0 | 529.4 | 422.2 | 460.6 | 467.0 | 473.3 | 479.7 | 492.5 |

| | | | | | | | | | | |
|---|---|---|---|---|---|---|---|---|---|---|
| Tamil Nadu | 773.5 | 800.6 | 857.1 | 1017.3 | 795.5 | 875.0 | 888.2 | 901.3 | 914.5 | 941.0 |
| Telangana | 434.5 | 446.6 | 471.8 | 543.2 | 444.3 | 479.8 | 485.7 | 491.4 | 497.4 | 509.2 |
| India | 23429.5 | 23774.7 | 24494.2 | 26533.2 | 23707.7 | 24719.7 | 24888.3 | 25050.0 | 25218.6 | 25555.8 |

**Supplementary Table 10: Percentage Change in Net Benefits for Meeting Unmet Needs for Cataract Surgery After Adjusting for Complication Rate.** All values are represented in percentages. NSS - National Sample Survey, PMJAY - Pradhan Mantri Jan Arogya Yojana, PHACO -Phacoemulsification, IOL-Intraocular lens, SICS -Small Incision Cataract Surgery.

| States | Mean Cost, NSS | Societal cost using mean IOL cost, Aravind Eye Hospital | Medical (direct) cost using mean IOL cost, Aravind Eye Hospital | Patient (indirect) cost, Aravind Eye Hospital | PHACO with hydrophobic acrylic IOL + Glaucoma, PMJAY | PHACO with hydrophobic acrylic IOL, PMJAY | PHACO with foldable IOL Unilateral, PMJAY | SICS with non-foldable IOL + Glaucoma, PMJAY | SICS with IOL Unilateral, PMJAY | SICS with non-foldable IOL, PMJAY |
|---|---|---|---|---|---|---|---|---|---|---|
| Jammu & Kashmir and Ladakh | -12.65 | 0.12 | 0.12 | 0.12 | 0.12 | 0.12 | 0.12 | 0.11 | 0.11 | 0.11 |
| Himachal Pradesh | -11.30 | 0.24 | 0.24 | 0.24 | 0.23 | 0.23 | 0.23 | 0.21 | 0.21 | 0.21 |
| Punjab | 23.89 | 0.61 | 0.61 | 0.61 | 0.59 | 0.59 | 0.59 | 0.54 | 0.54 | 0.54 |
| Uttarakhand | 45.97 | 0.28 | 0.28 | 0.28 | 0.27 | 0.27 | 0.27 | 0.25 | 0.25 | 0.25 |
| Haryana | -3.02 | 0.33 | 0.33 | 0.33 | 0.32 | 0.32 | 0.32 | 0.29 | 0.29 | 0.29 |
| NCT of Delhi | -10.41 | 0.26 | 0.26 | 0.26 | 0.25 | 0.25 | 0.25 | 0.23 | 0.23 | 0.23 |
| Rajasthan | 114.55 | 0.27 | 0.27 | 0.27 | 0.26 | 0.26 | 0.26 | 0.24 | 0.24 | 0.24 |
| Uttar | -36.76 | 0.30 | 0.30 | 0.30 | 0.29 | 0.29 | 0.29 | 0.26 | 0.26 | 0.26 |

| | | | | | | | | | | |
|---|---|---|---|---|---|---|---|---|---|---|
| Pradesh | | | | | | | | | | |
| Bihar | -1,360.87 | 0.24 | 0.24 | 0.24 | 0.23 | 0.23 | 0.23 | 0.21 | 0.21 | 0.21 |
| Sikkim | 2.36 | 0.06 | 0.06 | 0.06 | 0.05 | 0.05 | 0.05 | 0.05 | 0.05 | 0.05 |
| Arunachal Pradesh | 10.39 | 0.14 | 0.14 | 0.14 | 0.14 | 0.14 | 0.14 | 0.13 | 0.13 | 0.13 |
| Nagaland | -16.66 | 0.05 | 0.05 | 0.05 | 0.05 | 0.05 | 0.05 | 0.05 | 0.05 | 0.05 |
| Manipur | -143.42 | 0.05 | 0.05 | 0.05 | 0.05 | 0.05 | 0.05 | 0.05 | 0.05 | 0.05 |
| Mizoram | -28.23 | 0.18 | 0.18 | 0.18 | 0.17 | 0.17 | 0.17 | 0.16 | 0.16 | 0.16 |
| Tripura | -39.27 | 0.14 | 0.14 | 0.14 | 0.13 | 0.13 | 0.13 | 0.12 | 0.12 | 0.12 |
| Meghalaya | -135.55 | 0.08 | 0.08 | 0.08 | 0.08 | 0.08 | 0.08 | 0.07 | 0.07 | 0.07 |
| Assam | -37.19 | 0.14 | 0.14 | 0.14 | 0.13 | 0.13 | 0.13 | 0.12 | 0.12 | 0.12 |
| West Bengal | -10.65 | 0.21 | 0.21 | 0.21 | 0.21 | 0.21 | 0.21 | 0.19 | 0.19 | 0.19 |
| Jharkhand | 113.56 | 0.16 | 0.16 | 0.16 | 0.15 | 0.15 | 0.15 | 0.14 | 0.14 | 0.14 |
| Odisha | -17.26 | 0.19 | 0.19 | 0.19 | 0.18 | 0.18 | 0.18 | 0.16 | 0.16 | 0.16 |
| Chhattisgarh | -16.81 | 0.20 | 0.20 | 0.20 | 0.20 | 0.20 | 0.20 | 0.18 | 0.18 | 0.18 |
| Madhya Pradesh | -32.23 | 0.93 | 0.93 | 0.93 | 0.90 | 0.90 | 0.90 | 0.82 | 0.82 | 0.82 |
| Gujarat | -9.26 | 1.32 | 1.32 | 1.32 | 1.27 | 1.27 | 1.27 | 1.16 | 1.16 | 1.16 |
| Maharashtra | 13.49 | 0.35 | 0.35 | 0.35 | 0.33 | 0.33 | 0.33 | 0.30 | 0.30 | 0.30 |

| | | | | | | | | | | |
|---|---|---|---|---|---|---|---|---|---|---|
| Andhra Pradesh | 16.32 | 0.30 | 0.30 | 0.30 | 0.29 | 0.29 | 0.29 | 0.26 | 0.26 | 0.26 |
| Karnataka | 14.42 | 0.39 | 0.39 | 0.39 | 0.38 | 0.38 | 0.38 | 0.35 | 0.35 | 0.35 |
| Goa | -8.24 | 0.36 | 0.36 | 0.36 | 0.35 | 0.35 | 0.35 | 0.31 | 0.31 | 0.31 |
| Kerala | 11.59 | 0.22 | 0.22 | 0.22 | 0.22 | 0.22 | 0.22 | 0.20 | 0.20 | 0.20 |
| Tamil Nadu | 0.48 | 0.21 | 0.21 | 0.21 | 0.20 | 0.20 | 0.20 | 0.18 | 0.18 | 0.18 |
| Telangana | 13.70 | 0.33 | 0.33 | 0.33 | 0.32 | 0.32 | 0.32 | 0.29 | 0.29 | 0.29 |
| India | 0.08 | 0.08 | 0.07 | 5.24 | 7.57 | 6.51 | 6.37 | 6.20 | 6.06 | 5.81 |

**Supplementary Table 11: Percentage Change in Enhanced Coverage Costs for Meeting Unmet Needs for Cataract Surgery After Adjusting for Complication Rate.** All values are represented in percentages. NSS - National Sample Survey, PMJAY - Pradhan Mantri Jan Arogya Yojana, PHACO -Phacoemulsification, IOL-Intraocular lens, SICS -Small Incision Cataract Surgery.

| States | Mean Cost, NSS | Societal cost using mean IOL cost, Aravind Eye Hospital | Medical (direct) cost using mean IOL cost, Aravind Eye Hospital | Patient (indirect) cost, Aravind Eye Hospital | PHACO with hydrophobic acrylic IOL + Glaucoma, PMJAY | PHACO with hydrophobic acrylic IOL, PMJAY | PHACO with foldable IOL Unilateral, PMJAY | SICS with non-foldable IOL + Glaucoma, PMJAY | SICS with IOL Unilateral, PMJAY | SICS with non-foldable IOL, PMJAY |
|---|---|---|---|---|---|---|---|---|---|---|
| Jammu & Kashmir and Ladakh | 17.90 | 0.12 | 0.12 | 0.12 | 0.12 | 0.12 | 0.12 | 0.11 | 0.11 | 0.11 |
| Himachal Pradesh | 56.53 | 0.24 | 0.24 | 0.24 | 0.23 | 0.23 | 0.23 | 0.21 | 0.21 | 0.21 |
| Punjab | -27.53 | 0.61 | 0.61 | 0.61 | 0.59 | 0.59 | 0.59 | 0.54 | 0.54 | 0.54 |
| Uttarakhand | -47.31 | 0.28 | 0.28 | 0.28 | 0.27 | 0.27 | 0.27 | 0.25 | 0.25 | 0.25 |
| Haryana | 14.08 | 0.33 | 0.33 | 0.33 | 0.32 | 0.32 | 0.32 | 0.29 | 0.29 | 0.29 |
| NCT of Delhi | 359.15 | 0.26 | 0.26 | 0.26 | 0.25 | 0.25 | 0.25 | 0.23 | 0.23 | 0.23 |
| Rajasthan | -37.08 | 0.27 | 0.27 | 0.27 | 0.26 | 0.26 | 0.26 | 0.24 | 0.24 | 0.24 |
| Uttar | 21.43 | 0.30 | 0.30 | 0.30 | 0.29 | 0.29 | 0.29 | 0.26 | 0.26 | 0.26 |

| | | | | | | | | | | |
|---|---|---|---|---|---|---|---|---|---|---|
| Pradesh | | | | | | | | | | |
| Bihar | 31.06 | 0.24 | 0.24 | 0.24 | 0.23 | 0.23 | 0.23 | 0.21 | 0.21 | 0.21 |
| Sikkim | -7.19 | 0.06 | 0.06 | 0.06 | 0.05 | 0.05 | 0.05 | 0.05 | 0.05 | 0.05 |
| Arunachal Pradesh | -23.29 | 0.14 | 0.14 | 0.14 | 0.14 | 0.14 | 0.14 | 0.13 | 0.13 | 0.13 |
| Nagaland | 37.63 | 0.05 | 0.05 | 0.05 | 0.05 | 0.05 | 0.05 | 0.05 | 0.05 | 0.05 |
| Manipur | -63.00 | 0.05 | 0.05 | 0.05 | 0.05 | 0.05 | 0.05 | 0.05 | 0.05 | 0.05 |
| Mizoram | 0 | 0.18 | 0.18 | 0.18 | 0.17 | 0.17 | 0.17 | 0.16 | 0.16 | 0.16 |
| Tripura | 309.90 | 0.14 | 0.14 | 0.14 | 0.13 | 0.13 | 0.13 | 0.12 | 0.12 | 0.12 |
| Meghalaya | -78.39 | 0.08 | 0.08 | 0.08 | 0.08 | 0.08 | 0.08 | 0.07 | 0.07 | 0.07 |
| Assam | 66.65 | 0.14 | 0.14 | 0.14 | 0.13 | 0.13 | 0.13 | 0.12 | 0.12 | 0.12 |
| West Bengal | 16.61 | 0.21 | 0.21 | 0.21 | 0.21 | 0.21 | 0.21 | 0.19 | 0.19 | 0.19 |
| Jharkhand | -20.81 | 0.16 | 0.16 | 0.16 | 0.15 | 0.15 | 0.15 | 0.14 | 0.14 | 0.14 |
| Odisha | 26.61 | 0.19 | 0.19 | 0.19 | 0.18 | 0.18 | 0.18 | 0.16 | 0.16 | 0.16 |
| Chhattisgarh | 27.95 | 0.20 | 0.20 | 0.20 | 0.20 | 0.20 | 0.20 | 0.18 | 0.18 | 0.18 |
| Madhya Pradesh | 66.45 | 0.93 | 0.93 | 0.93 | 0.90 | 0.90 | 0.90 | 0.82 | 0.82 | 0.82 |
| Gujarat | 48.93 | 1.32 | 1.32 | 1.32 | 1.27 | 1.27 | 1.27 | 1.16 | 1.16 | 1.16 |
| Maharashtra | -23.95 | 0.35 | 0.35 | 0.35 | 0.33 | 0.33 | 0.33 | 0.30 | 0.30 | 0.30 |

| | | | | | | | | | | |
|---|---|---|---|---|---|---|---|---|---|---|
| Andhra Pradesh | -20.92 | 0.30 | 0.30 | 0.30 | 0.29 | 0.29 | 0.29 | 0.26 | 0.26 | 0.26 |
| Karnataka | -29.67 | 0.39 | 0.39 | 0.39 | 0.38 | 0.38 | 0.38 | 0.35 | 0.35 | 0.35 |
| Goa | 104.39 | 0.36 | 0.36 | 0.36 | 0.35 | 0.35 | 0.35 | 0.31 | 0.31 | 0.31 |
| Kerala | -22.23 | 0.22 | 0.22 | 0.22 | 0.22 | 0.22 | 0.22 | 0.20 | 0.20 | 0.20 |
| Tamil Nadu | -0.49 | 0.21 | 0.21 | 0.21 | 0.20 | 0.20 | 0.20 | 0.18 | 0.18 | 0.18 |
| Telangana | -27.39 | 0.33 | 0.33 | 0.33 | 0.32 | 0.32 | 0.32 | 0.29 | 0.29 | 0.29 |
| India | 1.00 | -0.01 | -0.01 | -0.01 | -0.03 | -0.03 | -0.03 | -0.05 | -0.05 | -0.05 |

### Supplementary Figures

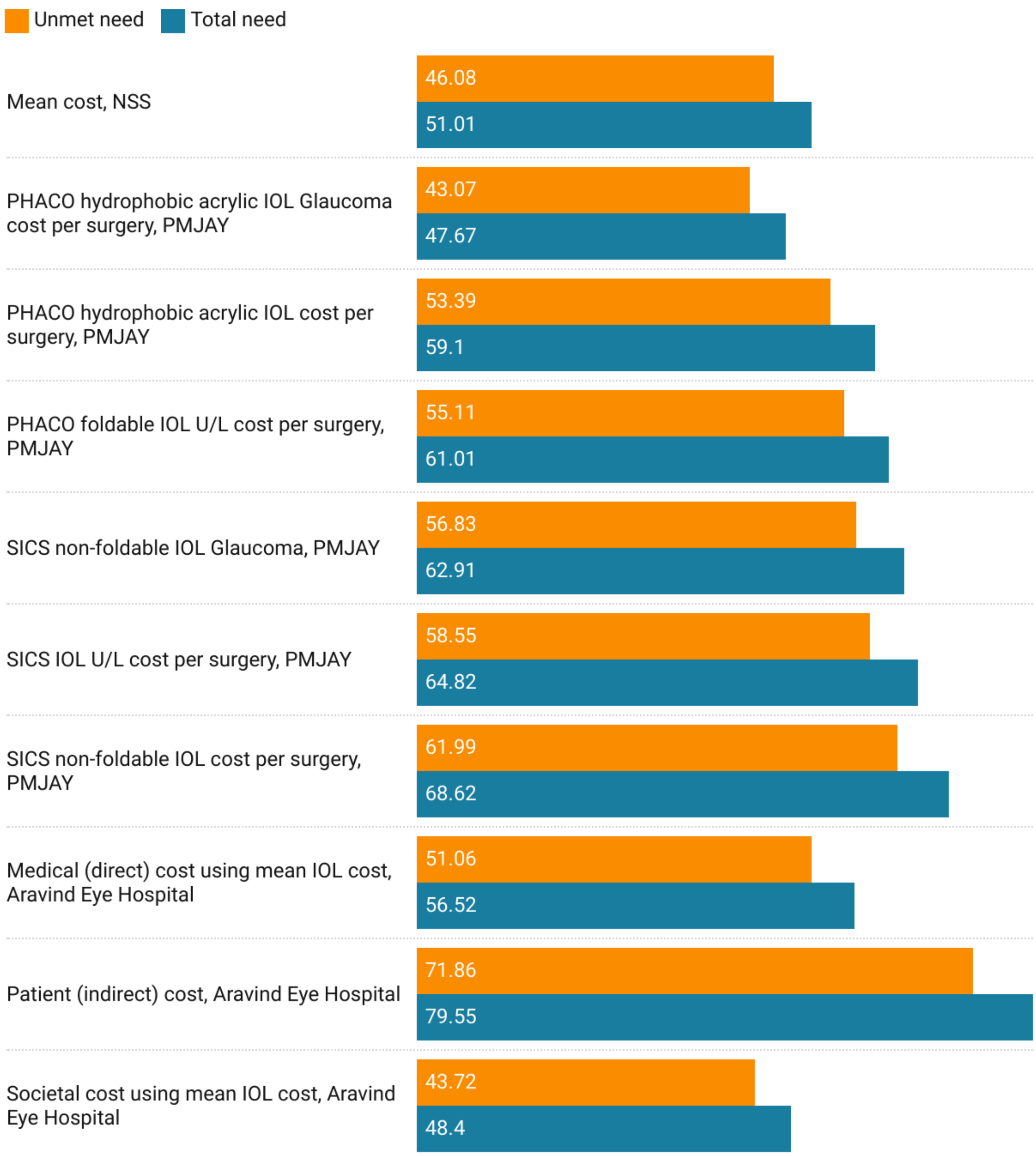


**Supplementary Figure 1: Net Benefits for Meeting Total and Unmet Needs for Universal Cataract Surgical Coverage in Jammu & Kashmir and Ladakh.** All costs are adjusted to millions of USD for the year 2020. Positive values depict net benefit while negative values depict net loss. Abbreviations: NSS - National Sample Survey, PMJAY - Pradhan Mantri Jan Arogya Yojana, PHACO - Phacoemulsification, IOL - Intraocular lens, SICS - Small Incision Cataract Surgery.

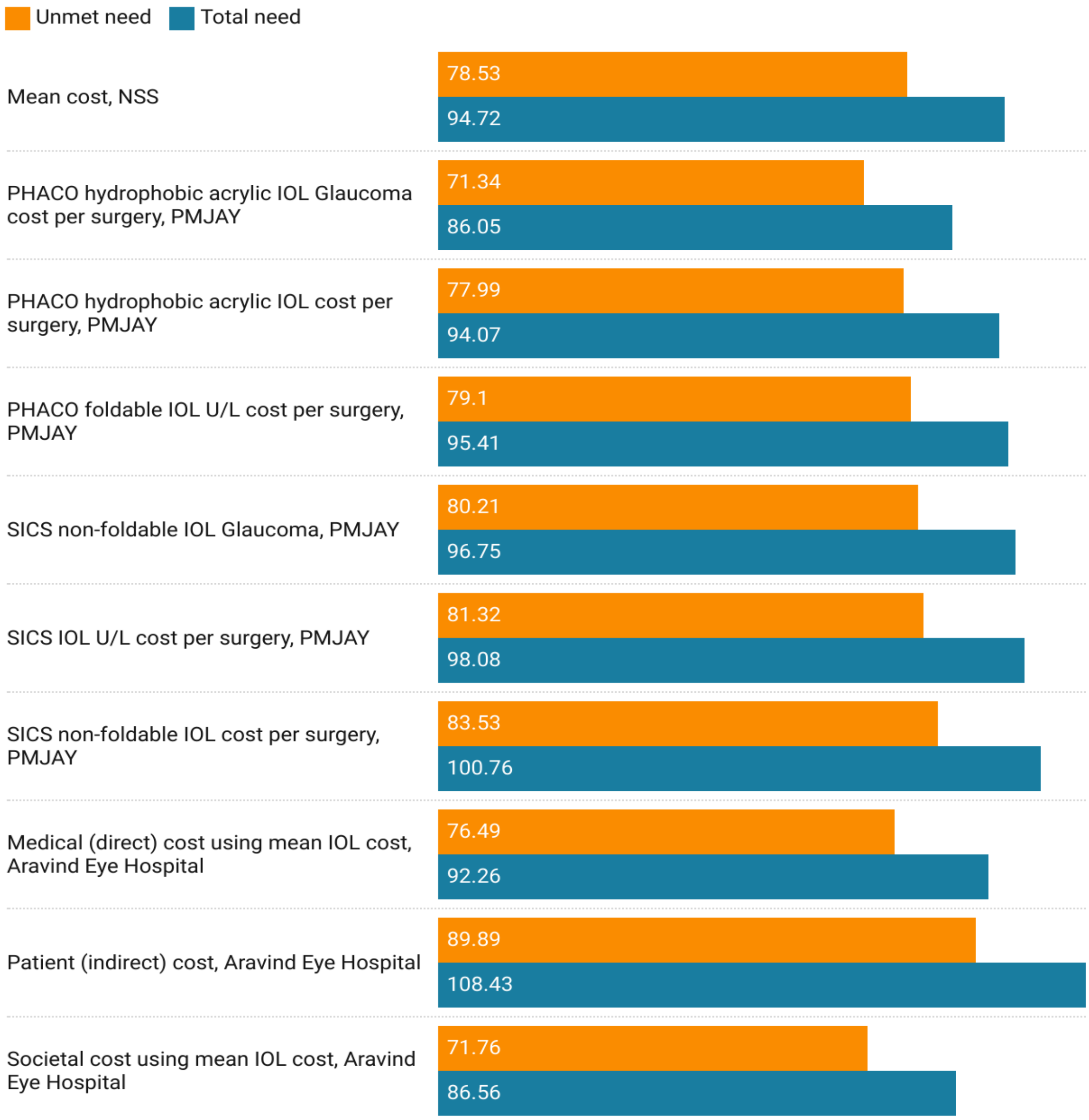


**Supplementary Figure 2: Net Benefits for Meeting Total and Unmet Needs for Universal Cataract Surgical Coverage in Himachal Pradesh.** All costs are adjusted to millions of USD for the year 2020. Positive values depict net benefit while negative values depict net loss. Abbreviations: NSS - National Sample Survey, PMJAY - Pradhan Mantri Jan Arogya Yojana, PHACO - Phacoemulsification, IOL - Intraocular lens, SICS - Small Incision Cataract Surgery.

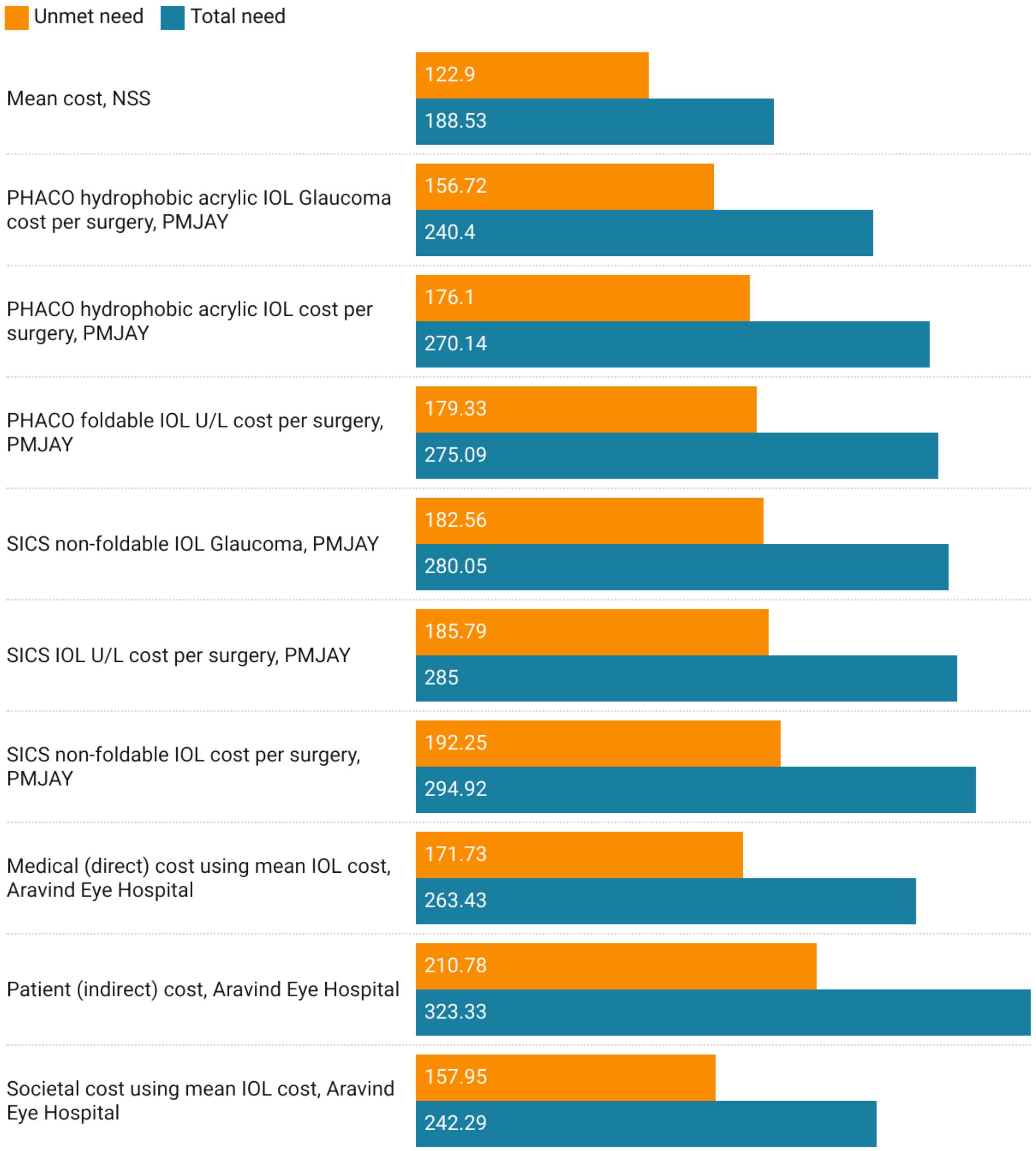


**Supplementary Figure 3: Net Benefits for Meeting Total and Unmet Needs for Universal Cataract Surgical Coverage in Punjab.** All costs are adjusted to millions of USD for the year 2020. Positive values depict net benefit while negative values depict net loss. Abbreviations: NSS - National Sample Survey, PMJAY - Pradhan Mantri Jan Arogya Yojana, PHACO - Phacoemulsification, IOL - Intraocular lens, SICS - Small Incision Cataract Surgery.

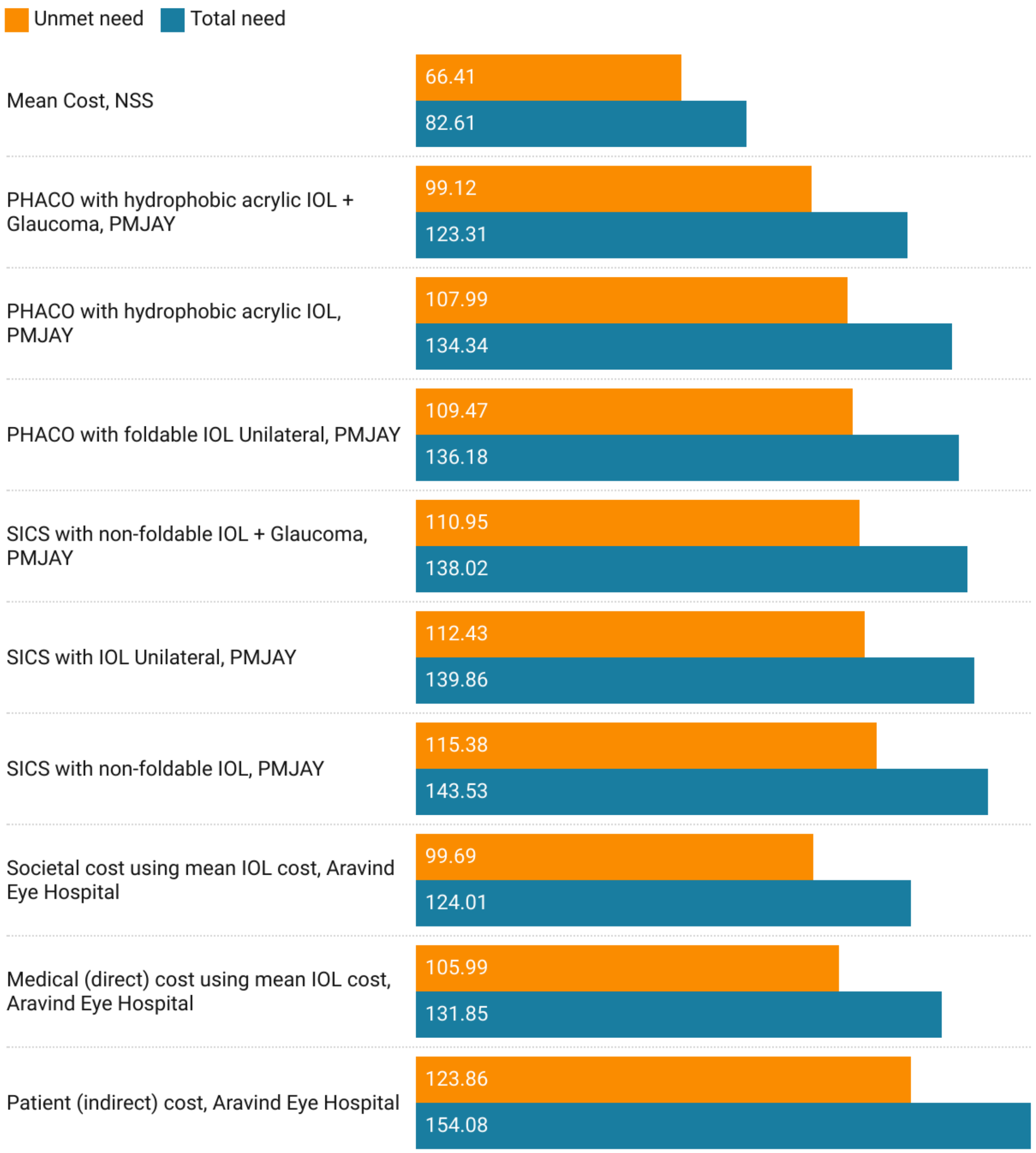


**Supplementary Figure 4: Net Benefits for Meeting Total and Unmet Needs for Universal Cataract Surgical Coverage in Uttarakhand.** All costs are adjusted to millions of USD for the year 2020. Positive values depict net benefit while negative values depict net loss. Abbreviations: NSS - National Sample Survey, PMJAY - Pradhan Mantri Jan Arogya Yojana, PHACO - Phacoemulsification, IOL - Intraocular lens, SICS - Small Incision Cataract Surgery.

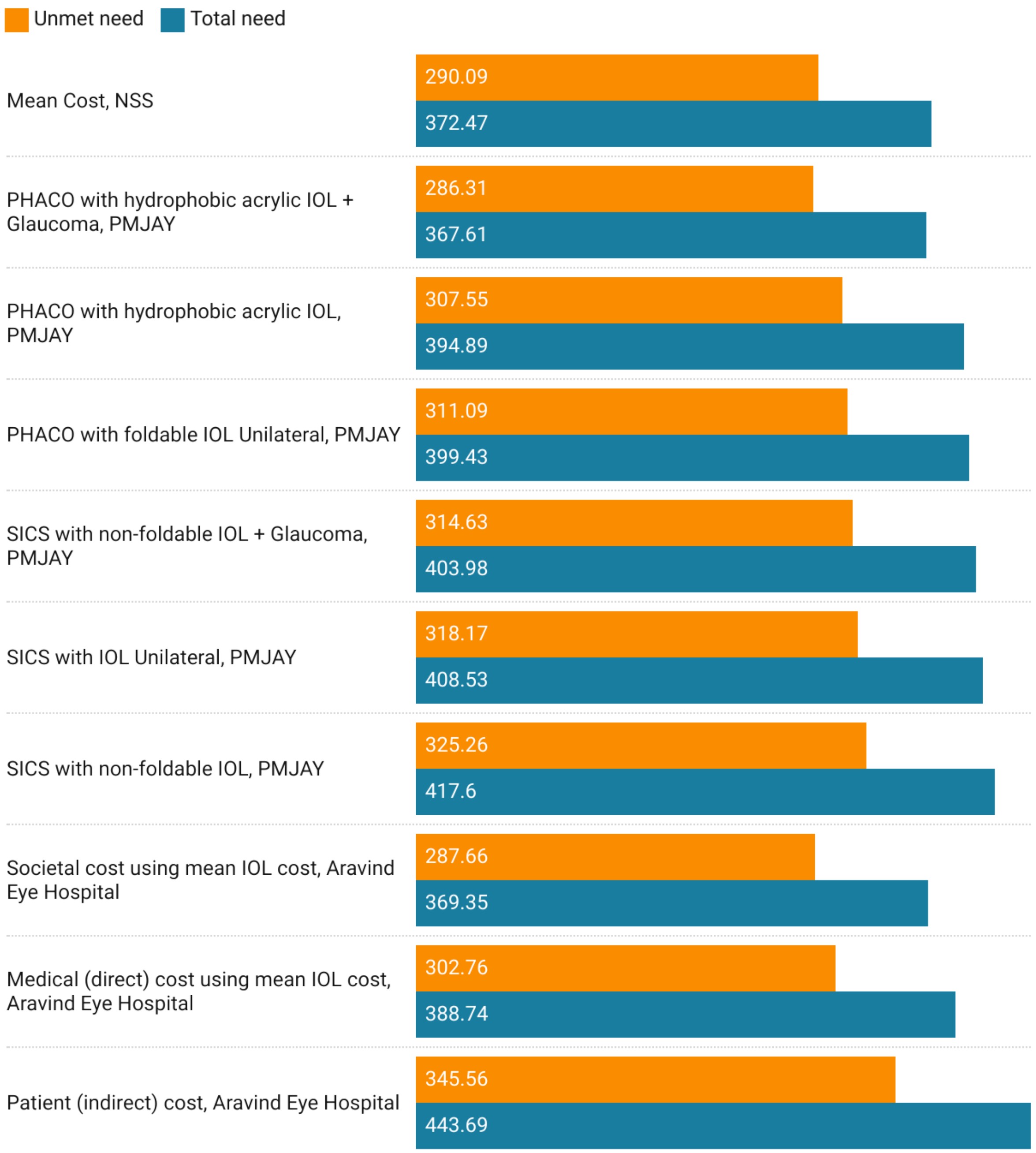


**Supplementary Figure 5: Net Benefits for Meeting Total and Unmet Needs for Universal Cataract Surgical Coverage in Haryana.** All costs are adjusted to millions of USD for the year 2020. Positive values depict net benefit while negative values depict net loss. Abbreviations: NSS - National Sample Survey, PMJAY - Pradhan Mantri Jan Arogya Yojana, PHACO - Phacoemulsification, IOL - Intraocular lens, SICS - Small Incision Cataract Surgery.

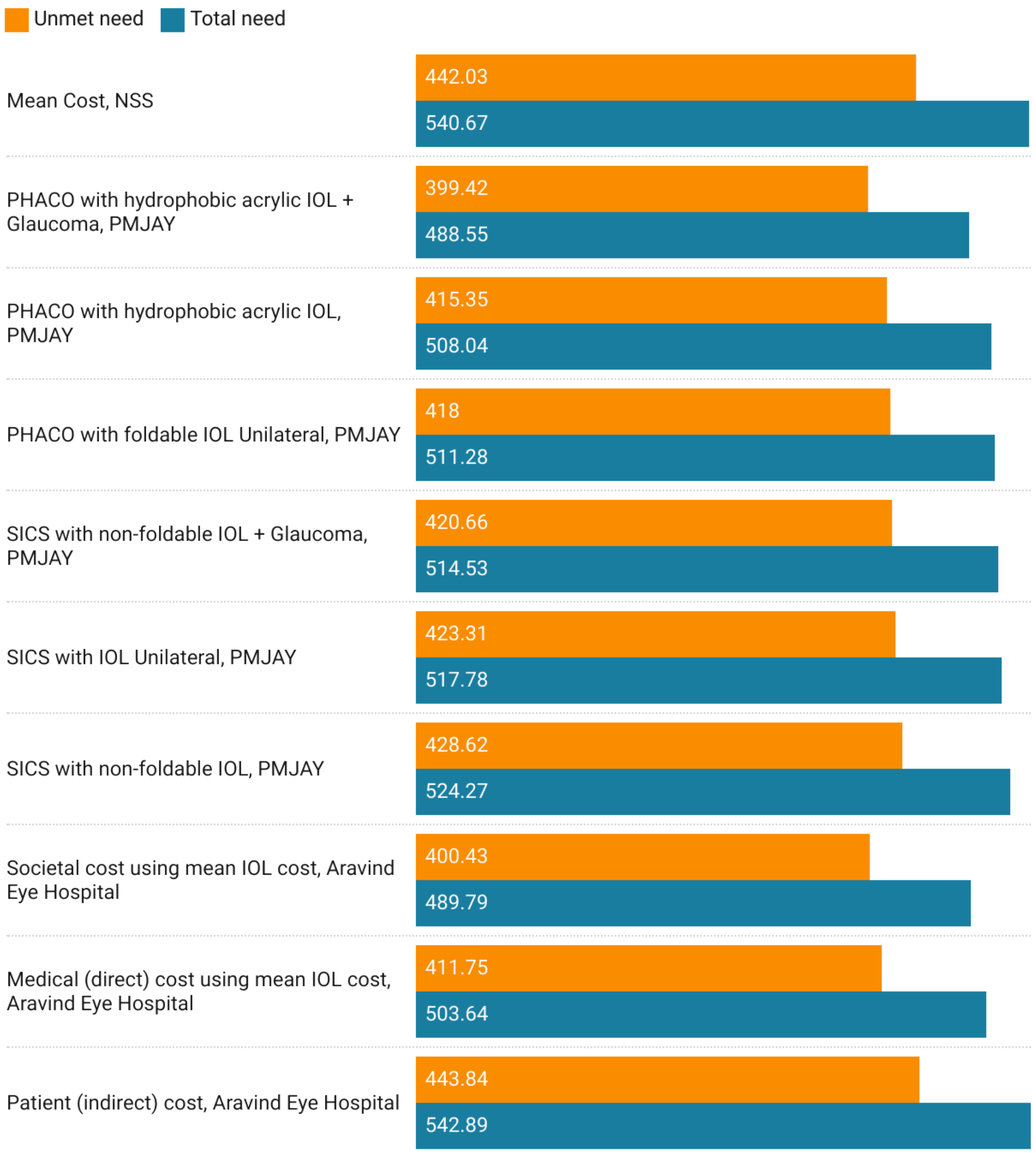


**Supplementary Figure 6: Net Benefits for Meeting Total and Unmet Needs for Universal Cataract Surgical Coverage in Delhi.** All costs are adjusted to millions of USD for the year 2020. Positive values depict net benefit while negative values depict net loss. Abbreviations: NSS - National Sample Survey, PMJAY - Pradhan Mantri Jan Arogya Yojana, PHACO - Phacoemulsification, IOL - Intraocular lens, SICS - Small Incision Cataract Surgery.

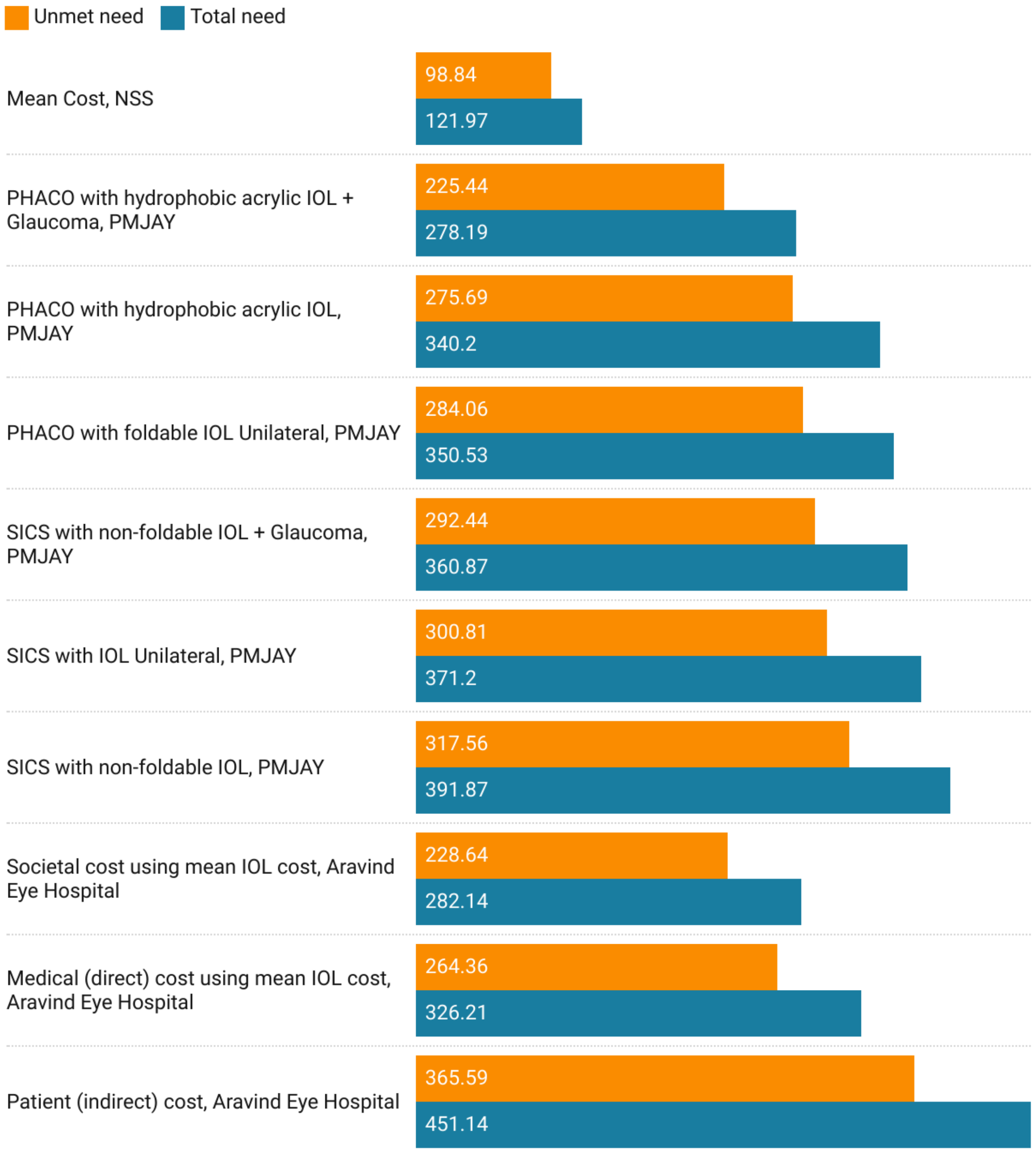


**Supplementary Figure 7: Net Benefits for Meeting Total and Unmet Needs for Universal Cataract Surgical Coverage in Rajasthan.** All costs are adjusted to millions of USD for the year 2020. Positive values depict net benefit while negative values depict net loss. Abbreviations: NSS - National Sample Survey, PMJAY - Pradhan Mantri Jan Arogya Yojana, PHACO - Phacoemulsification, IOL - Intraocular lens, SICS - Small Incision Cataract Surgery.

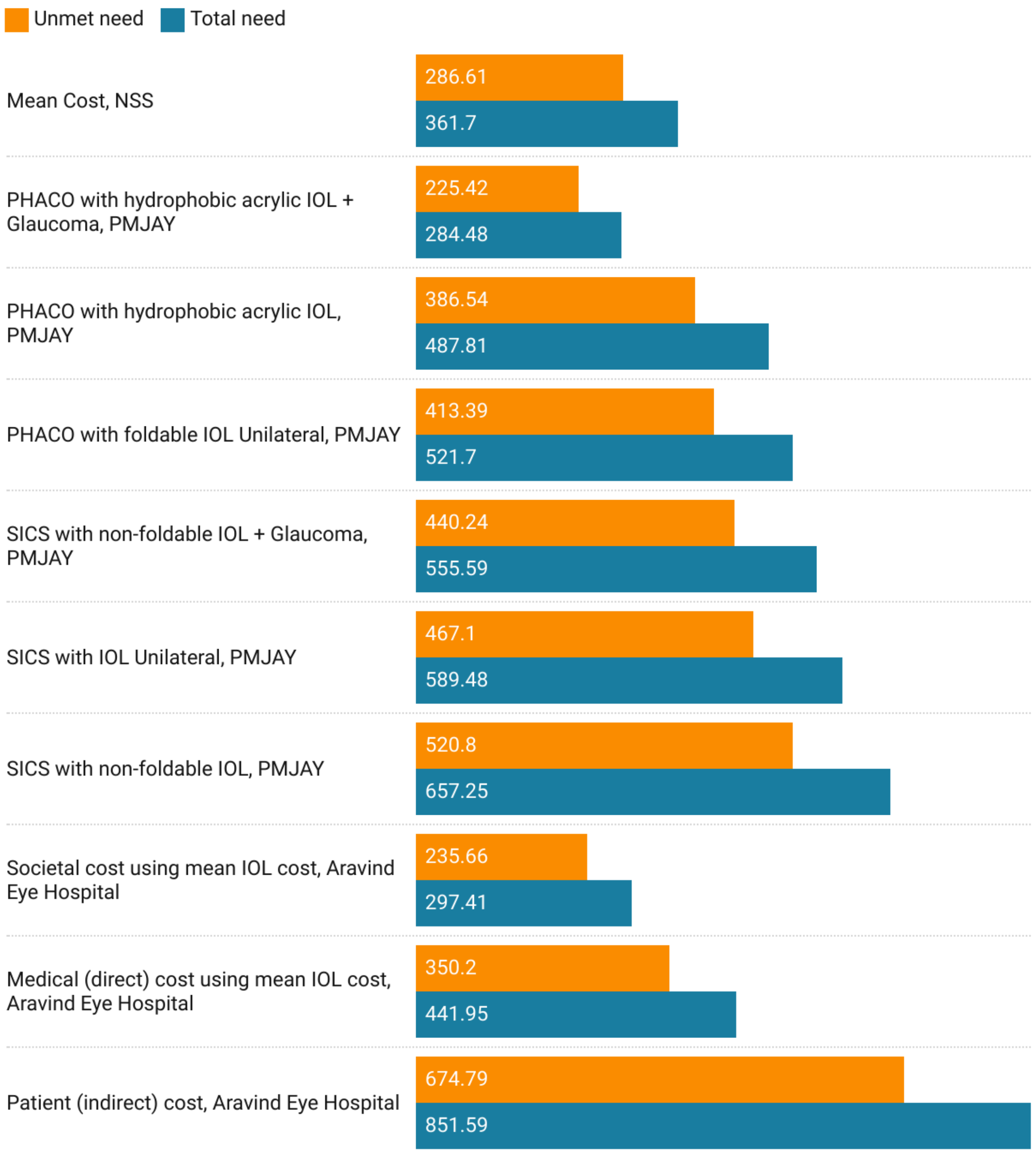


**Supplementary Figure 8: Net Benefits for Meeting Total and Unmet Needs for Universal Cataract Surgical Coverage in Uttar Pradesh.** All costs are adjusted to millions of USD for the year 2020. Positive values depict net benefit while negative values depict net loss. Abbreviations: NSS - National Sample Survey, PMJAY - Pradhan Mantri Jan Arogya Yojana, PHACO - Phacoemulsification, IOL - Intraocular lens, SICS - Small Incision Cataract Surgery.

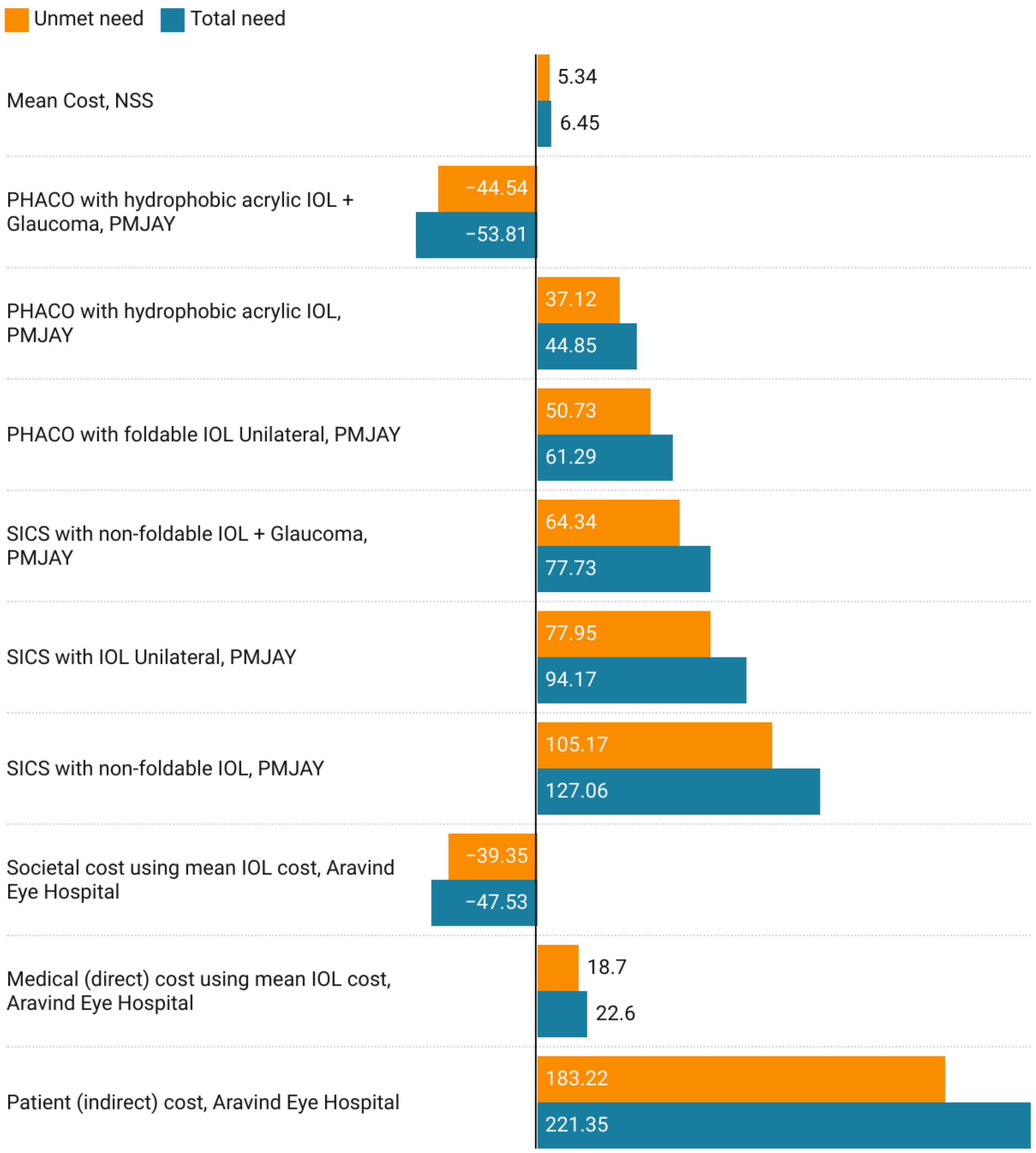


**Supplementary Figure 9: Net Benefits for Meeting Total and Unmet Needs for Universal Cataract Surgical Coverage in Bihar.** All costs are adjusted to millions of USD for the year 2020. Positive values depict net benefit while negative values depict net loss. Abbreviations: NSS - National Sample Survey, PMJAY - Pradhan Mantri Jan Arogya Yojana, PHACO - Phacoemulsification, IOL - Intraocular lens, SICS - Small Incision Cataract Surgery.

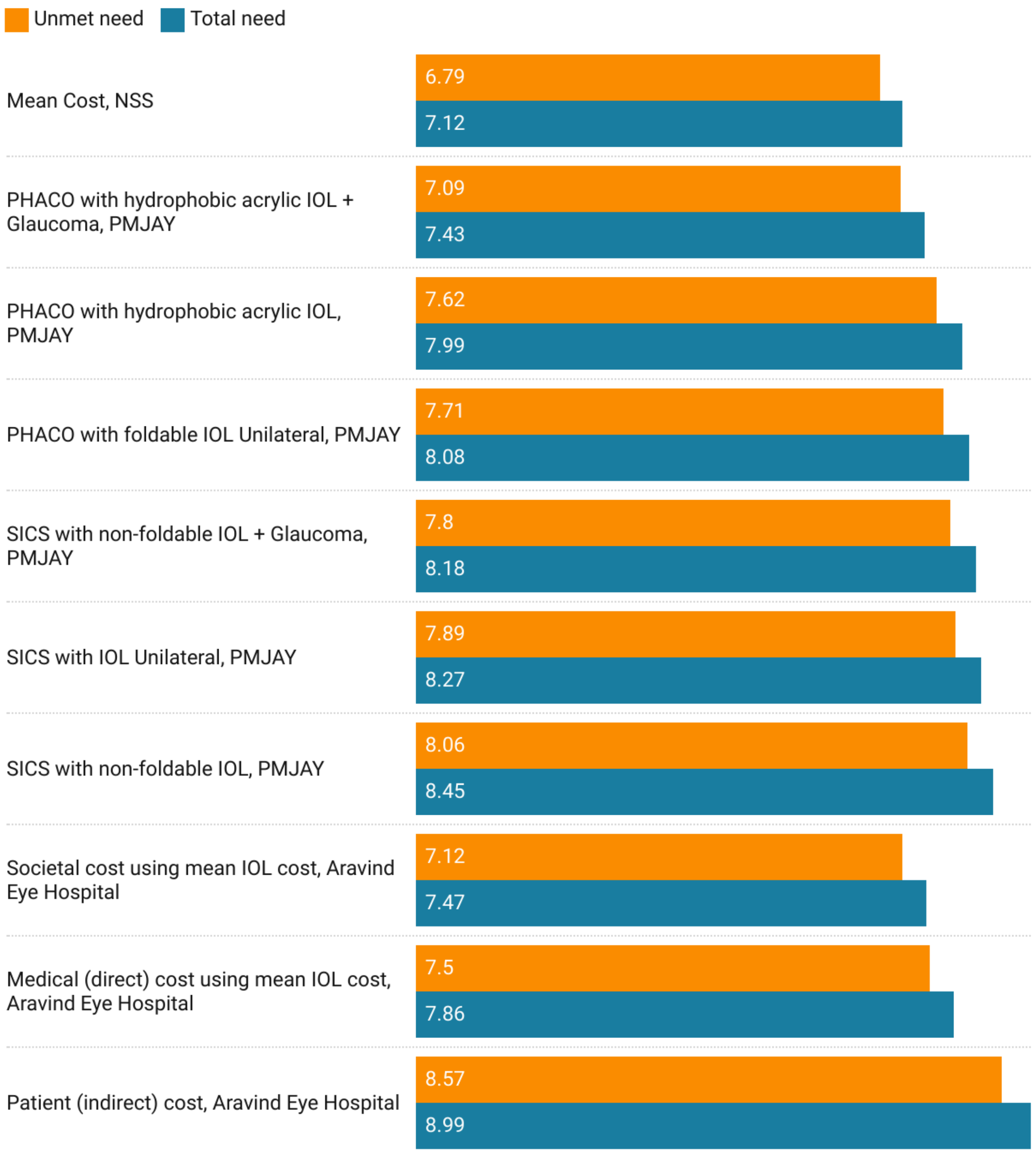


**Supplementary Figure 10: Net Benefits for Meeting Total and Unmet Needs for Universal Cataract Surgical Coverage in Sikkim.** All costs are adjusted to millions of USD for the year 2020. Positive values depict net benefit while negative values depict net loss. Abbreviations: NSS - National Sample Survey, PMJAY - Pradhan Mantri Jan Arogya Yojana, PHACO - Phacoemulsification, IOL - Intraocular lens, SICS - Small Incision Cataract Surgery.

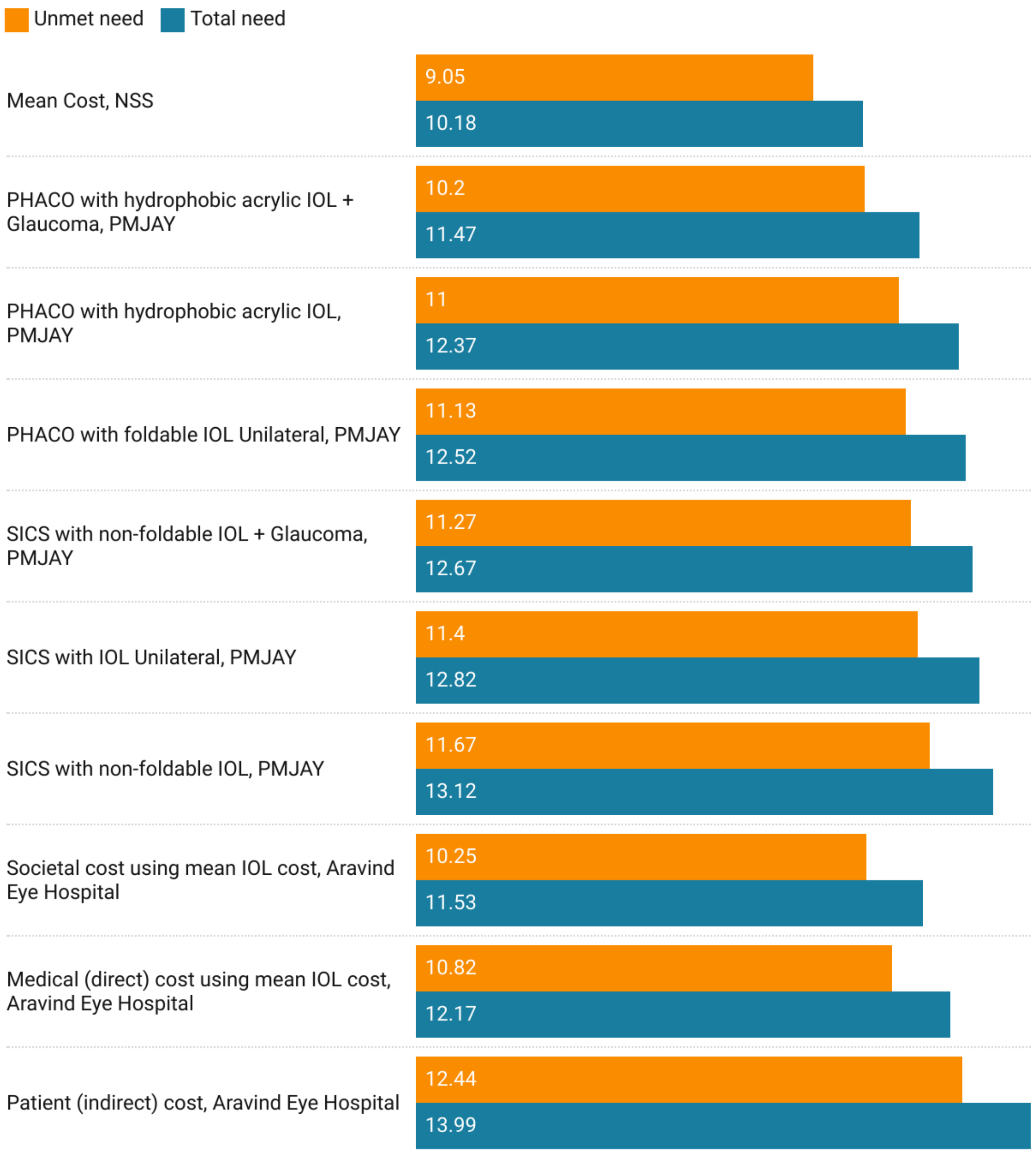


**Supplementary Figure 11: Net Benefits for Meeting Total and Unmet Needs for Universal Cataract Surgical Coverage in Arunachal Pradesh.** All costs are adjusted to millions of USD for the year 2020. Positive values depict net benefit while negative values depict net loss. Abbreviations: NSS - National Sample Survey, PMJAY - Pradhan Mantri Jan Arogya Yojana, PHACO - Phacoemulsification, IOL - Intraocular lens, SICS - Small Incision Cataract Surgery.

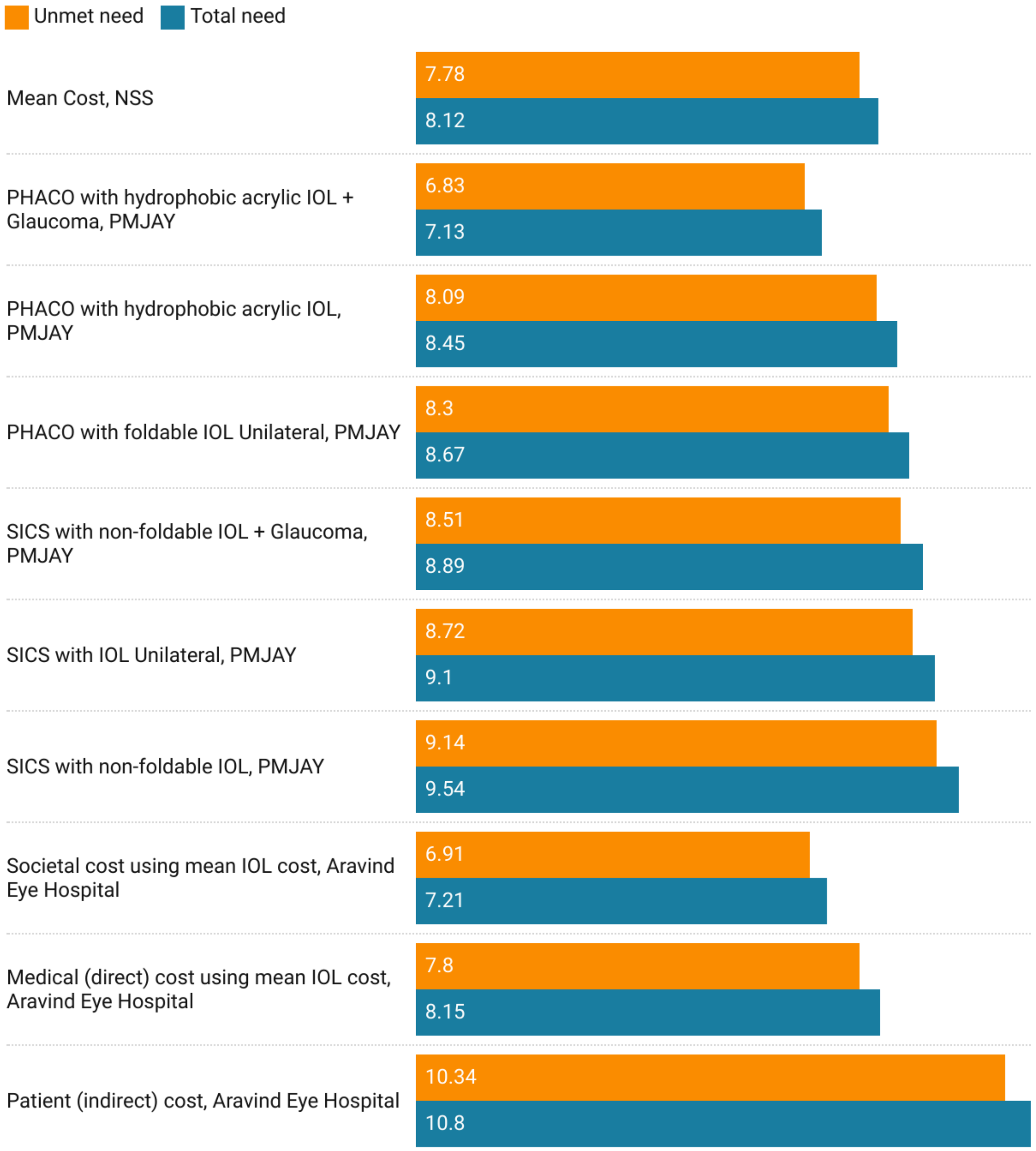


**Supplementary Figure 12: Net Benefits for Meeting Total and Unmet Needs for Universal Cataract Surgical Coverage in Nagaland.** All costs are adjusted to millions of USD for the year 2020. Positive values depict net benefit while negative values depict net loss. Abbreviations: NSS - National Sample Survey, PMJAY - Pradhan Mantri Jan Arogya Yojana, PHACO - Phacoemulsification, IOL - Intraocular lens, SICS - Small Incision Cataract Surgery.

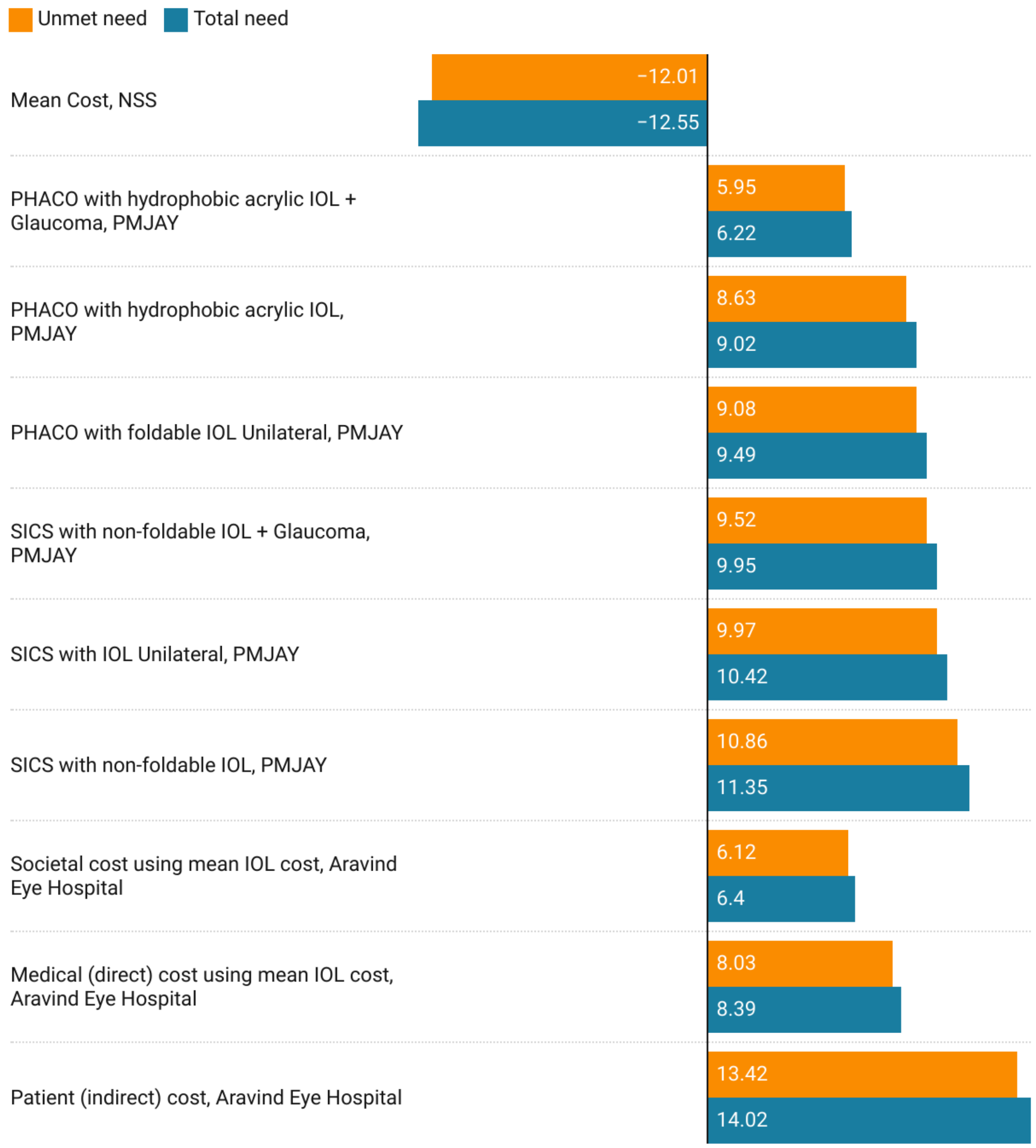


**Supplementary Figure 13: Net Benefits for Meeting Total and Unmet Needs for Universal Cataract Surgical Coverage in Manipur.** All costs are adjusted to millions of USD for the year 2020. Positive values depict net benefit while negative values depict net loss. Abbreviations: NSS - National Sample Survey, PMJAY - Pradhan Mantri Jan Arogya Yojana, PHACO - Phacoemulsification, IOL - Intraocular lens, SICS - Small Incision Cataract Surgery.

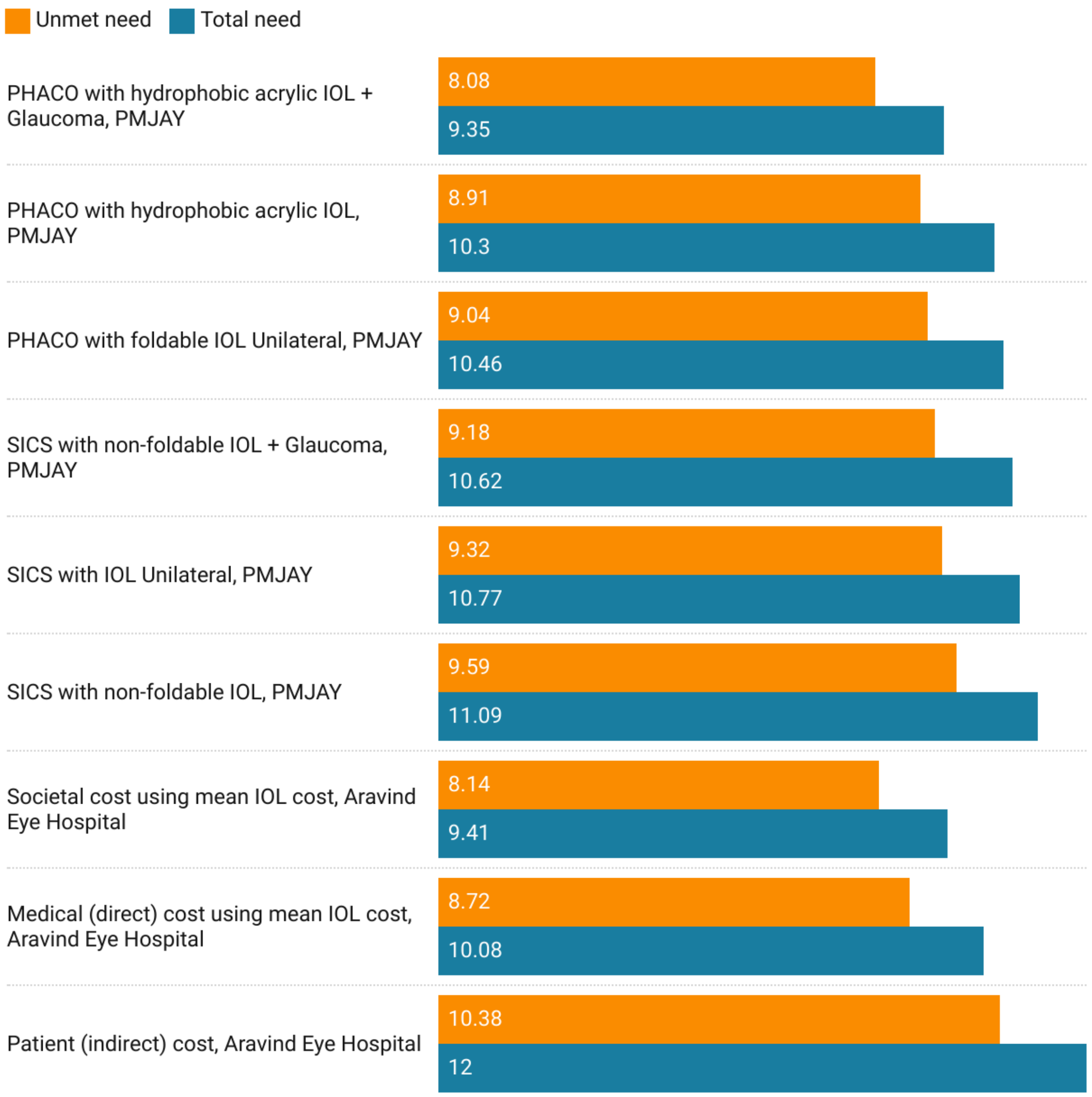


**Supplementary Figure 14: Net Benefits for Meeting Total and Unmet Needs for Universal Cataract Surgical Coverage in Mizoram.** All costs are adjusted to millions of USD for the year 2020. Positive values depict net benefit while negative values depict net loss. Abbreviations: NSS - National Sample Survey, PMJAY - Pradhan Mantri Jan Arogya Yojana, PHACO - Phacoemulsification, IOL - Intraocular lens, SICS - Small Incision Cataract Surgery.

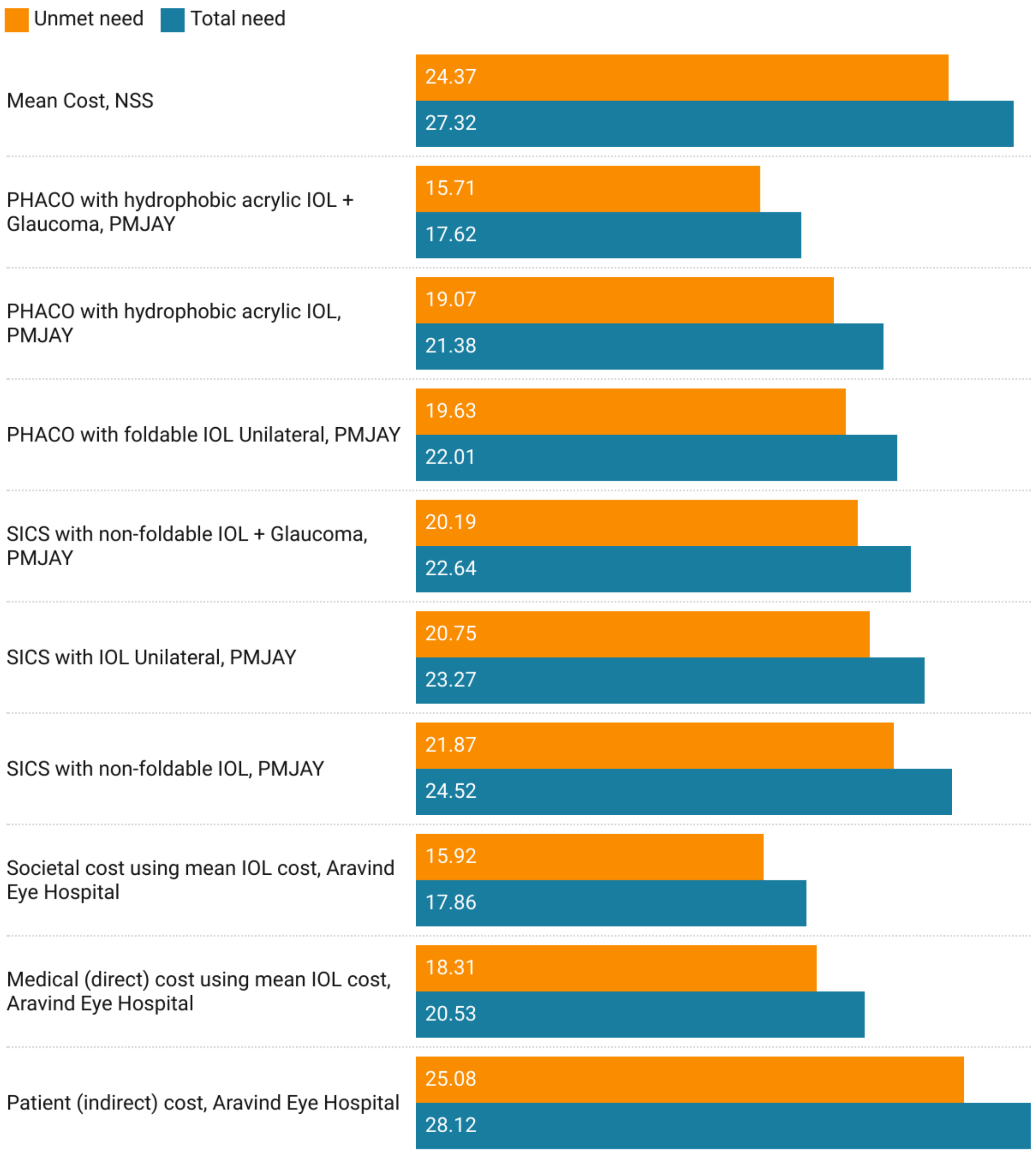


**Supplementary Figure 15: Net Benefits for Meeting Total and Unmet Needs for Universal Cataract Surgical Coverage in Tripura.** All costs are adjusted to millions of USD for the year 2020. Positive values depict net benefit while negative values depict net loss. Abbreviations: NSS - National Sample Survey, PMJAY - Pradhan Mantri Jan Arogya Yojana, PHACO - Phacoemulsification, IOL - Intraocular lens, SICS - Small Incision Cataract Surgery.

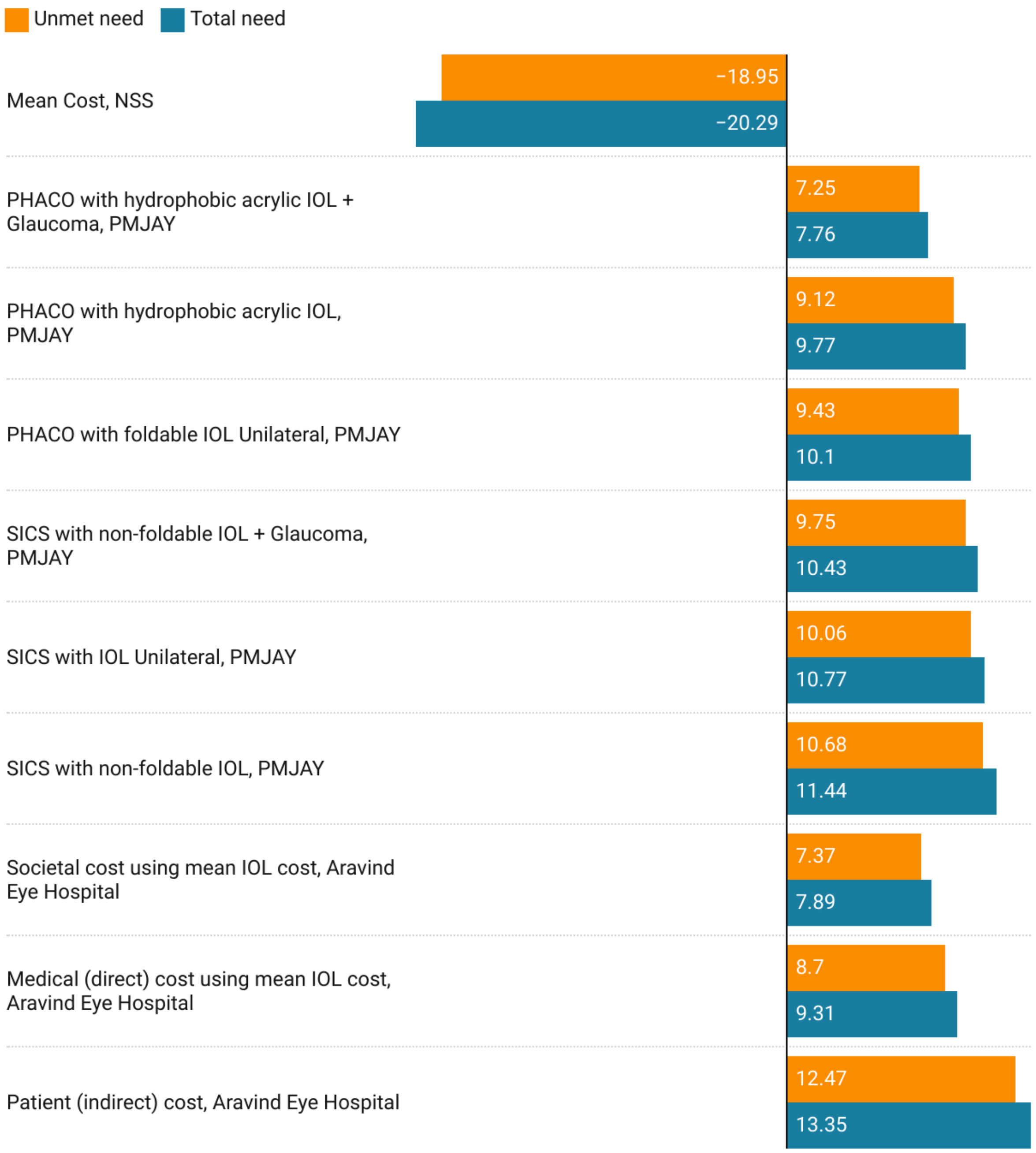


**Supplementary Figure 16: Net Benefits for Meeting Total and Unmet Needs for Universal Cataract Surgical Coverage in Meghalaya.** All costs are adjusted to millions of USD for the year 2020. Positive values depict net benefit while negative values depict net loss. Abbreviations: NSS - National Sample Survey, PMJAY - Pradhan Mantri Jan Arogya Yojana, PHACO - Phacoemulsification, IOL - Intraocular lens, SICS - Small Incision Cataract Surgery.

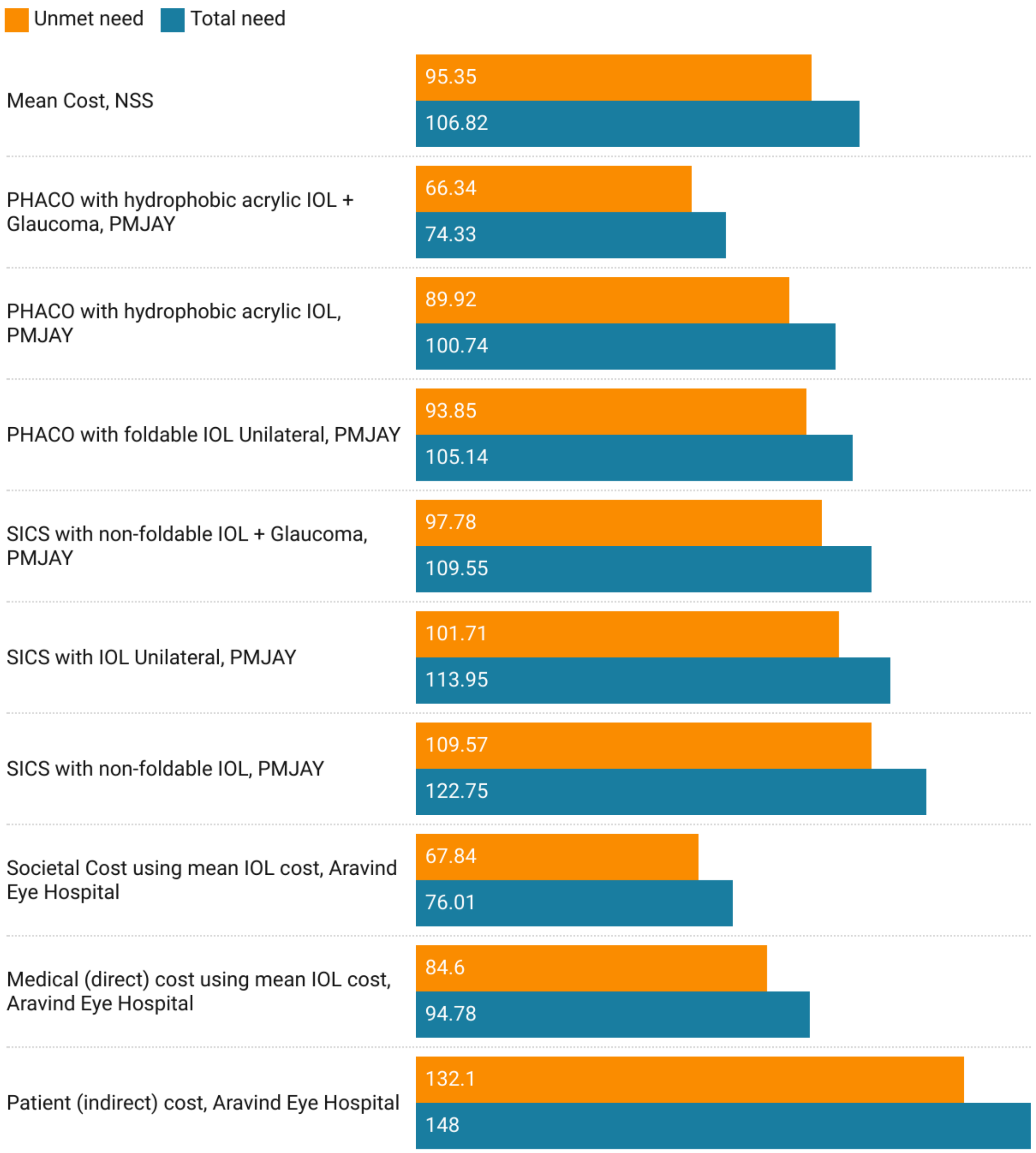


**Supplementary Figure 17: Net Benefits for Meeting Total and Unmet Needs for Universal Cataract Surgical Coverage in Assam.** All costs are adjusted to millions of USD for the year 2020. Positive values depict net benefit while negative values depict net loss. Abbreviations: NSS - National Sample Survey, PMJAY - Pradhan Mantri Jan Arogya Yojana, PHACO - Phacoemulsification, IOL - Intraocular lens, SICS - Small Incision Cataract Surgery.

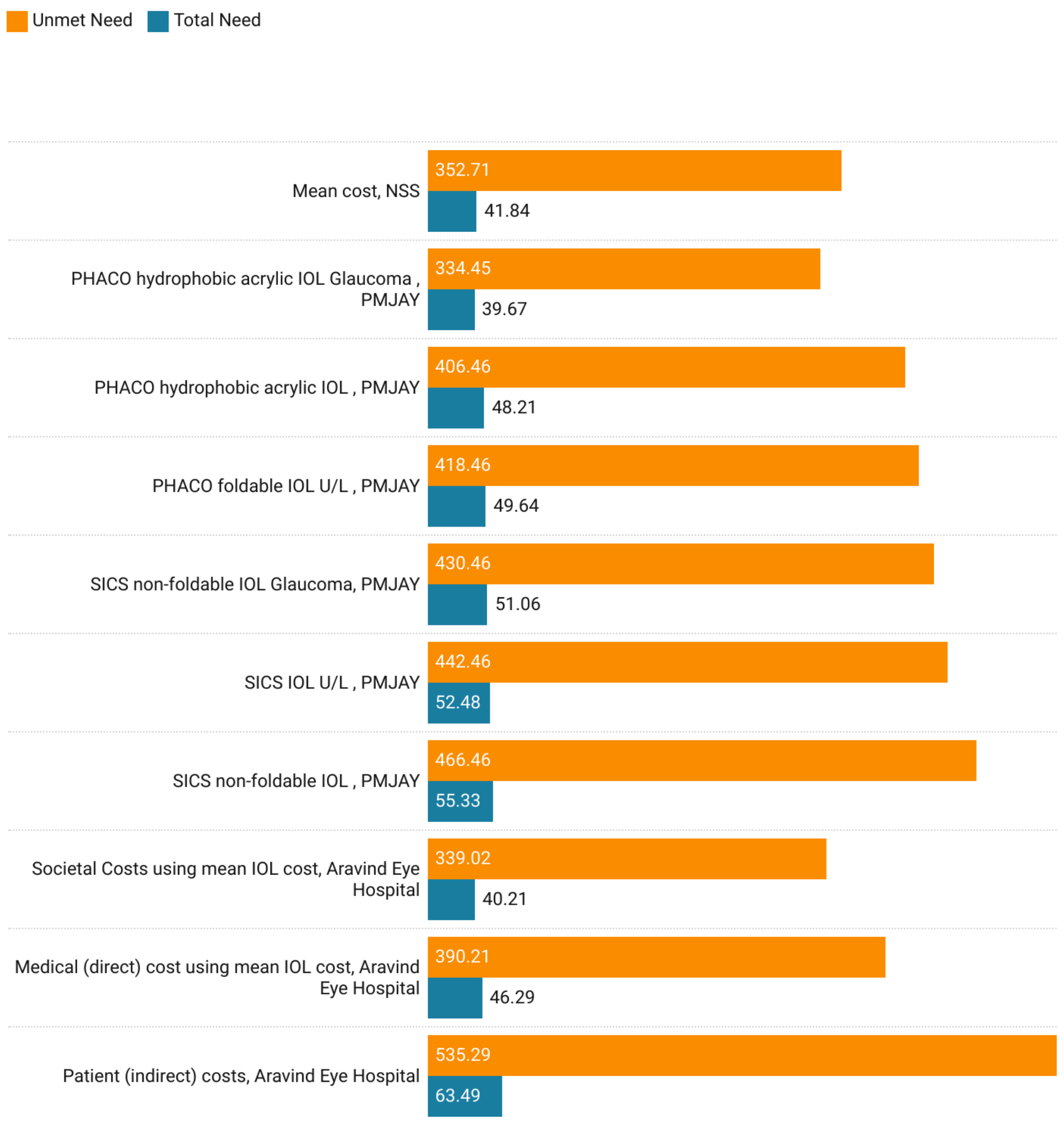


**Supplementary Figure 18: Net Benefits for Meeting Total and Unmet Needs for Universal Cataract Surgical Coverage in West Bengal.** All costs are adjusted to millions of USD for the year 2020. Positive values depict net benefit while negative values depict net loss. Abbreviations: NSS - National Sample Survey, PMJAY - Pradhan Mantri Jan Arogya Yojana, PHACO - Phacoemulsification, IOL - Intraocular lens, SICS - Small Incision Cataract Surgery.

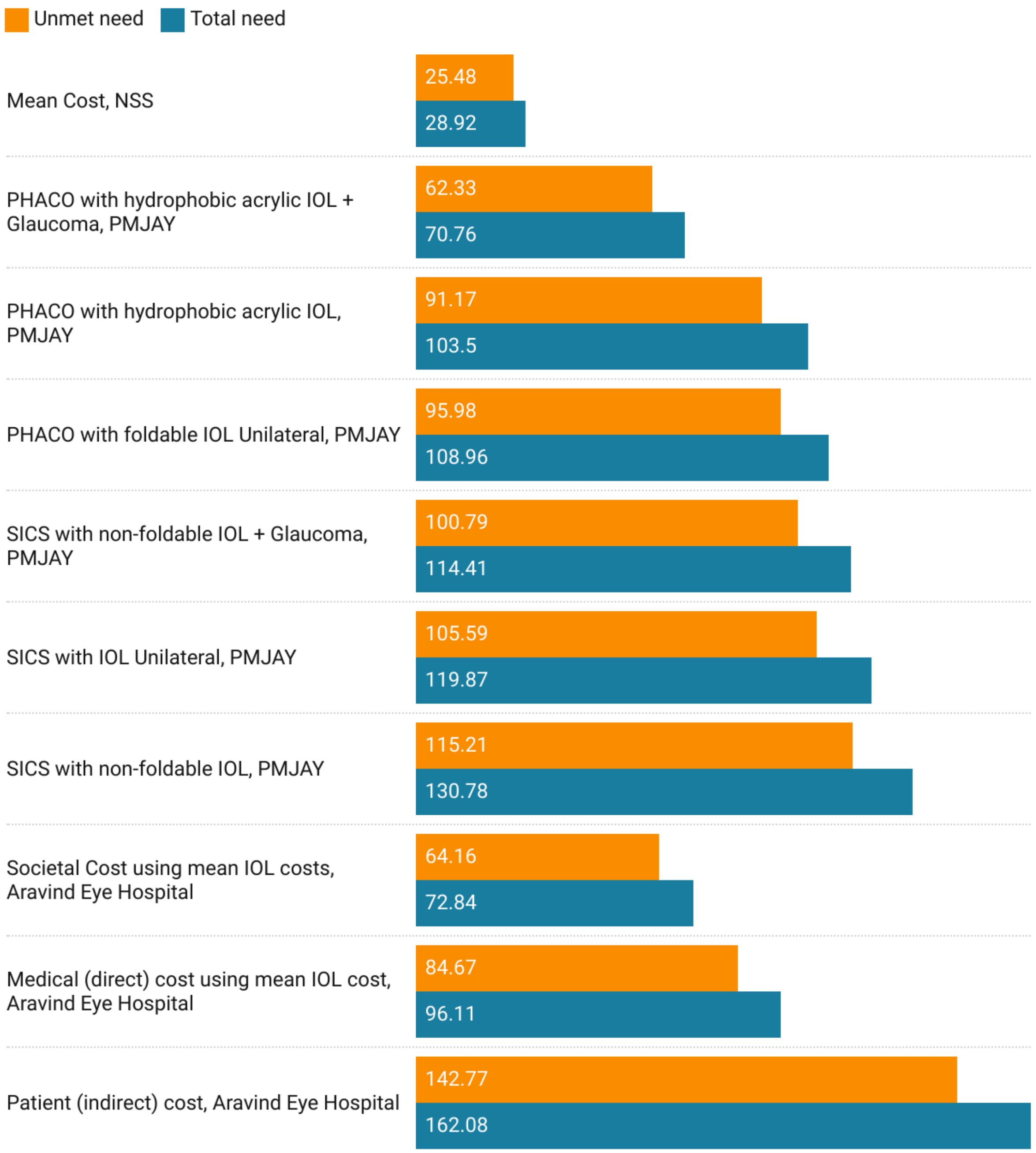


**Supplementary Figure 19: Net Benefits for Meeting Total and Unmet Needs for Universal Cataract Surgical Coverage in Jharkhand.** All costs are adjusted to millions of USD for the year 2020. Positive values depict net benefit while negative values depict net loss. Abbreviations: NSS - National Sample Survey, PMJAY - Pradhan Mantri Jan Arogya Yojana, PHACO - Phacoemulsification, IOL - Intraocular lens, SICS - Small Incision Cataract Surgery.

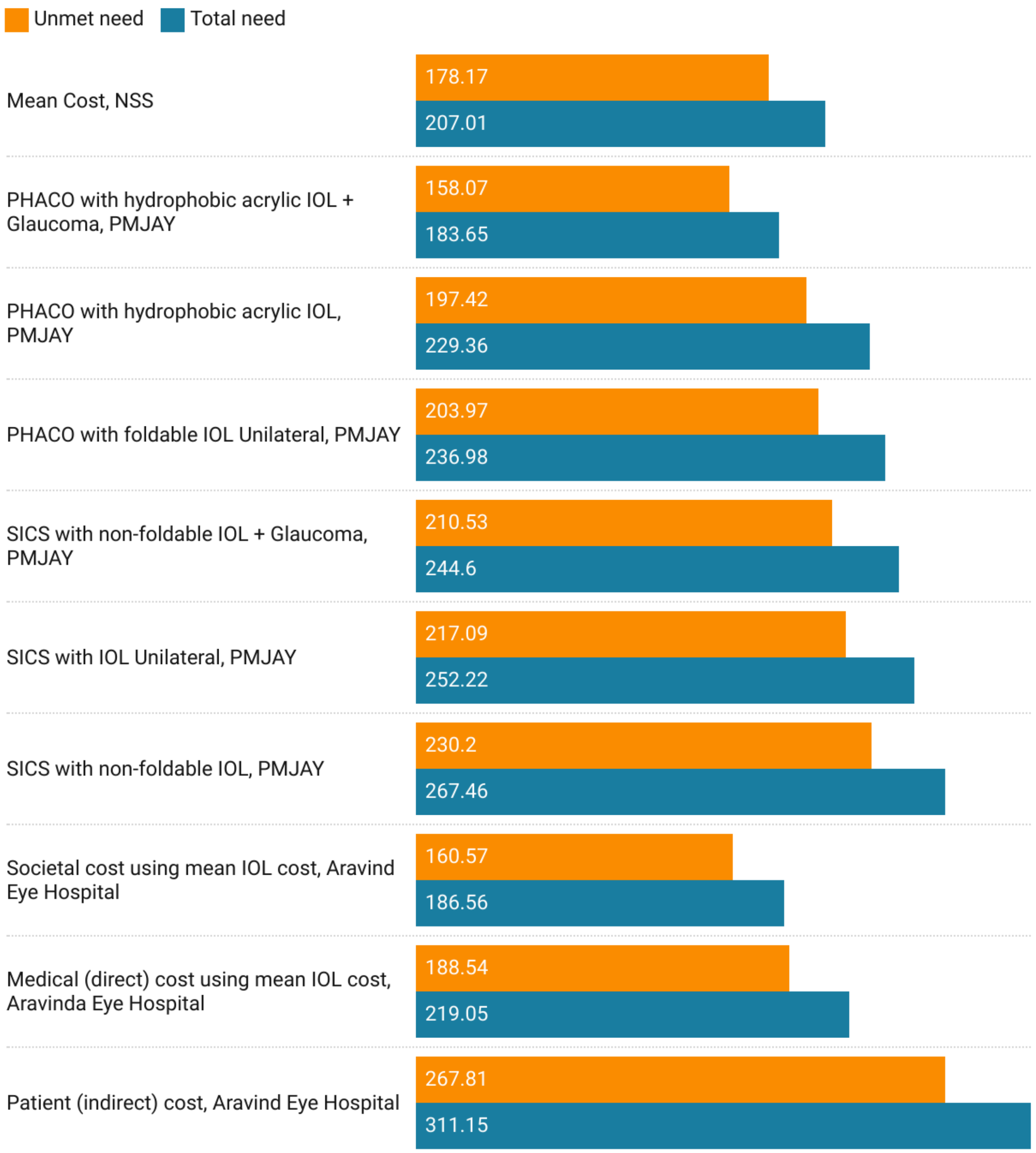


**Supplementary Figure 20: Net Benefits for Meeting Total and Unmet Needs for Universal Cataract Surgical Coverage in Odisha.** All costs are adjusted to millions of USD for the year 2020. Positive values depict net benefit while negative values depict net loss. Abbreviations: NSS - National Sample Survey, PMJAY - Pradhan Mantri Jan Arogya Yojana, PHACO - Phacoemulsification, IOL - Intraocular lens, SICS - Small Incision Cataract Surgery.

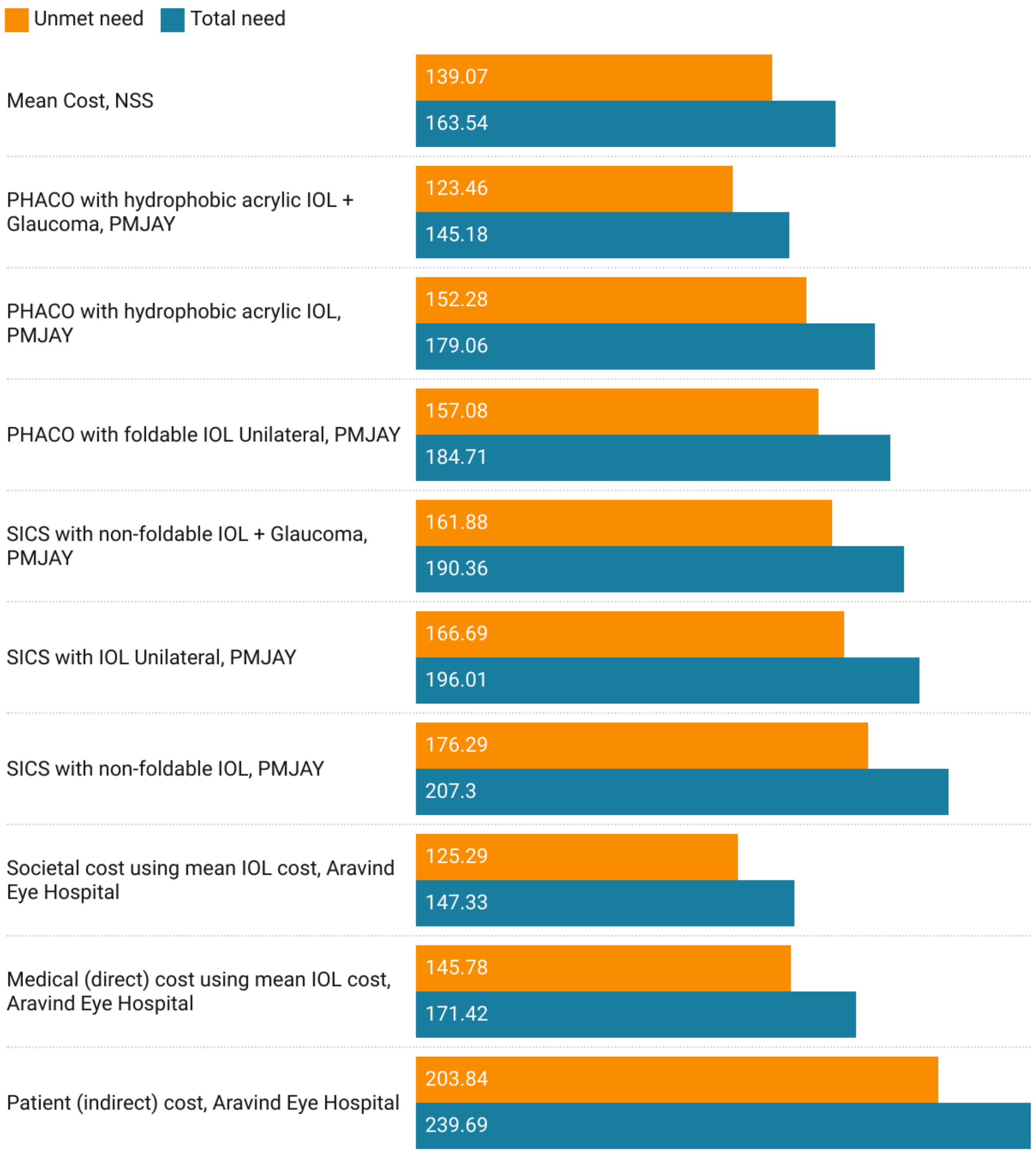


**Supplementary Figure 21: Net Benefits for Meeting Total and Unmet Needs for Universal Cataract Surgical Coverage in Chhattisgarh.** All costs are adjusted to millions of USD for the year 2020. Positive values depict net benefit while negative values depict net loss. Abbreviations: NSS - National Sample Survey, PMJAY - Pradhan Mantri Jan Arogya Yojana, PHACO - Phacoemulsification, IOL - Intraocular lens, SICS - Small Incision Cataract Surgery.

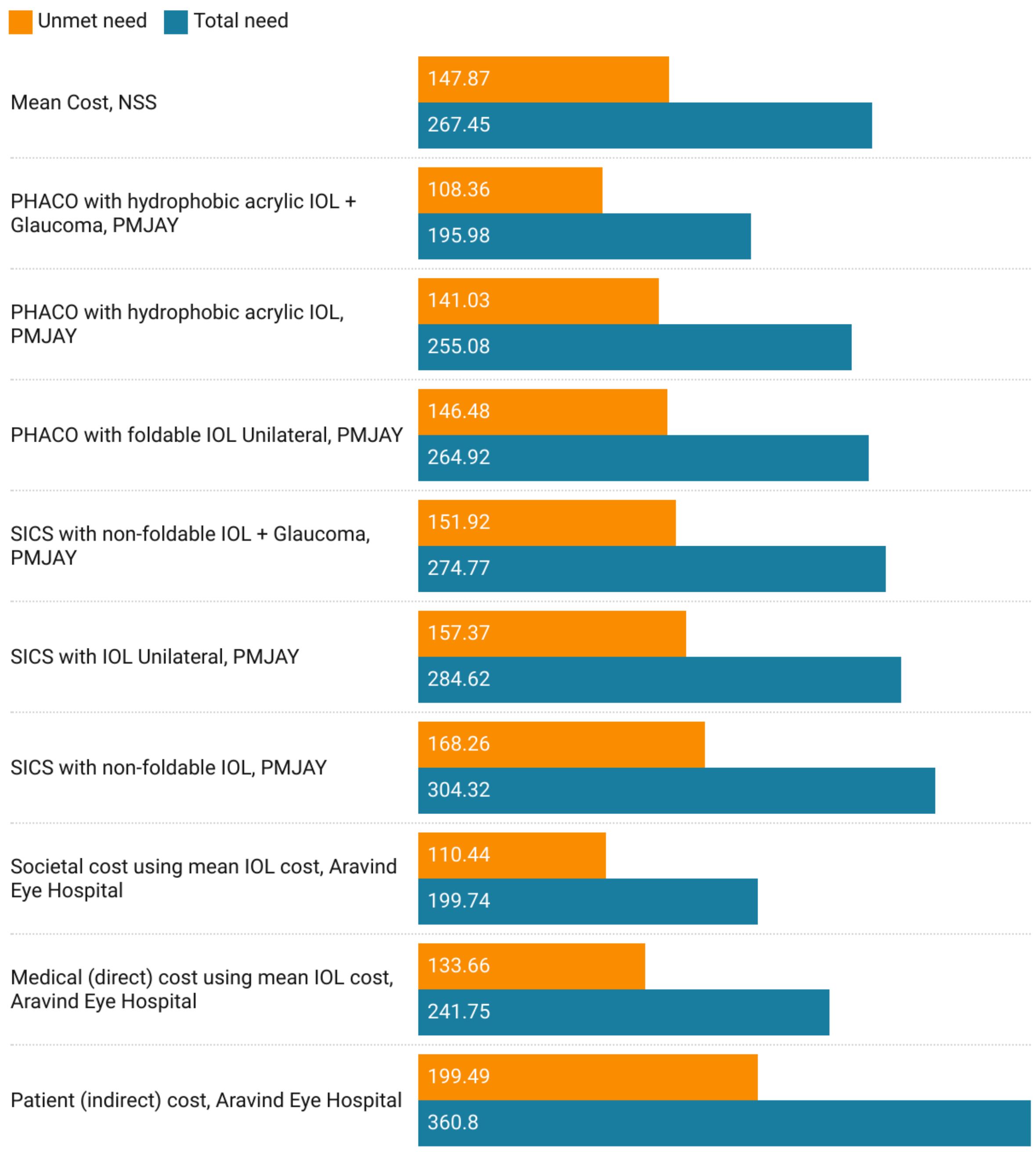


**Supplementary Figure 22: Net Benefits for Meeting Total and Unmet Needs for Universal Cataract Surgical Coverage in Madhya Pradesh.** All costs are adjusted to millions of USD for the year 2020. Positive values depict net benefit while negative values depict net loss. Abbreviations: NSS - National Sample Survey, PMJAY - Pradhan Mantri Jan Arogya Yojana, PHACO - Phacoemulsification, IOL - Intraocular lens, SICS - Small Incision Cataract Surgery.

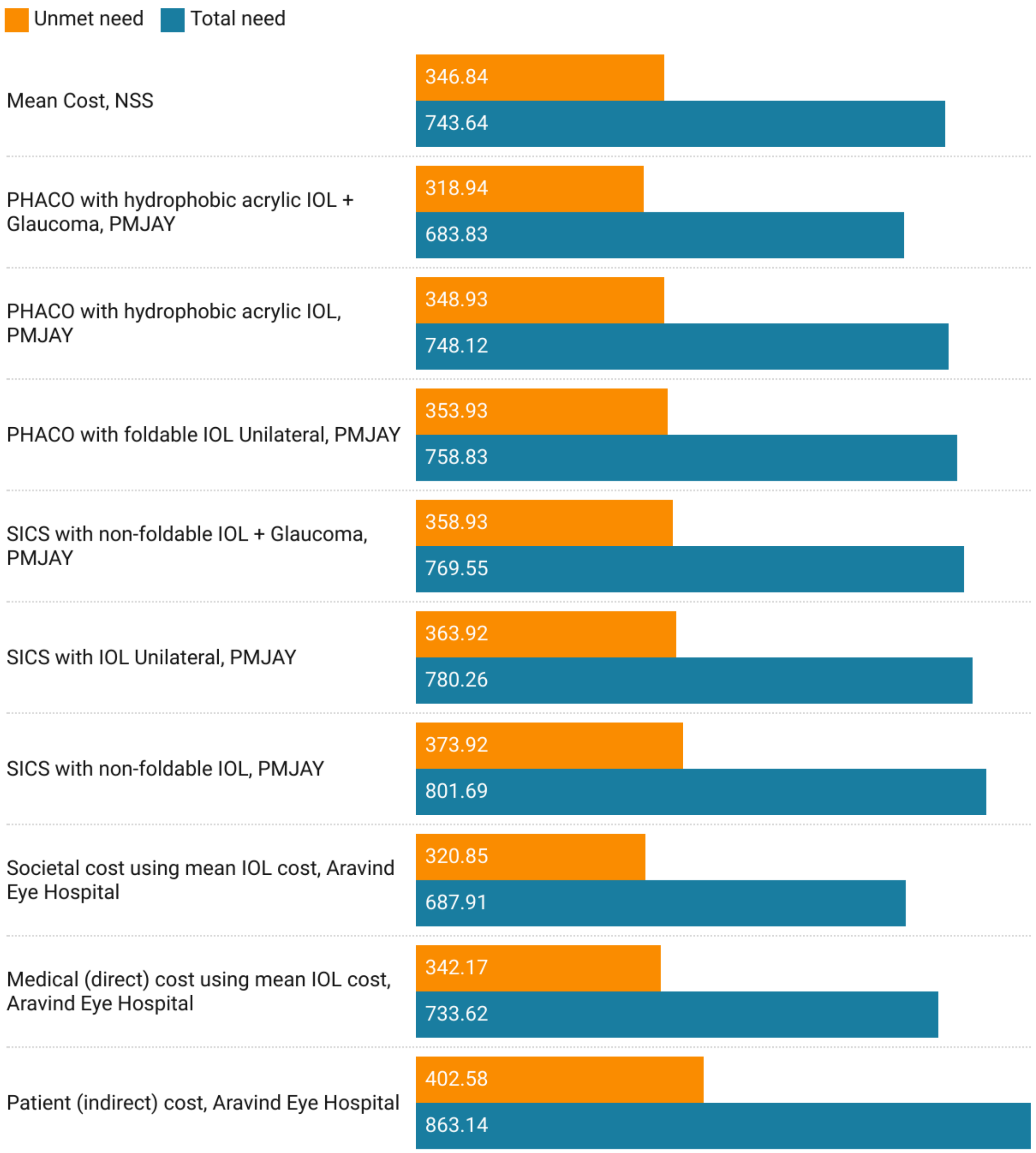


**Supplementary Figure 23: Net Benefits for Meeting Total and Unmet Needs for Universal Cataract Surgical Coverage in Gujarat.** All costs are adjusted to millions of USD for the year 2020. Positive values depict net benefit while negative values depict net loss. Abbreviations: NSS - National Sample Survey, PMJAY - Pradhan Mantri Jan Arogya Yojana, PHACO - Phacoemulsification, IOL - Intraocular lens, SICS - Small Incision Cataract Surgery.

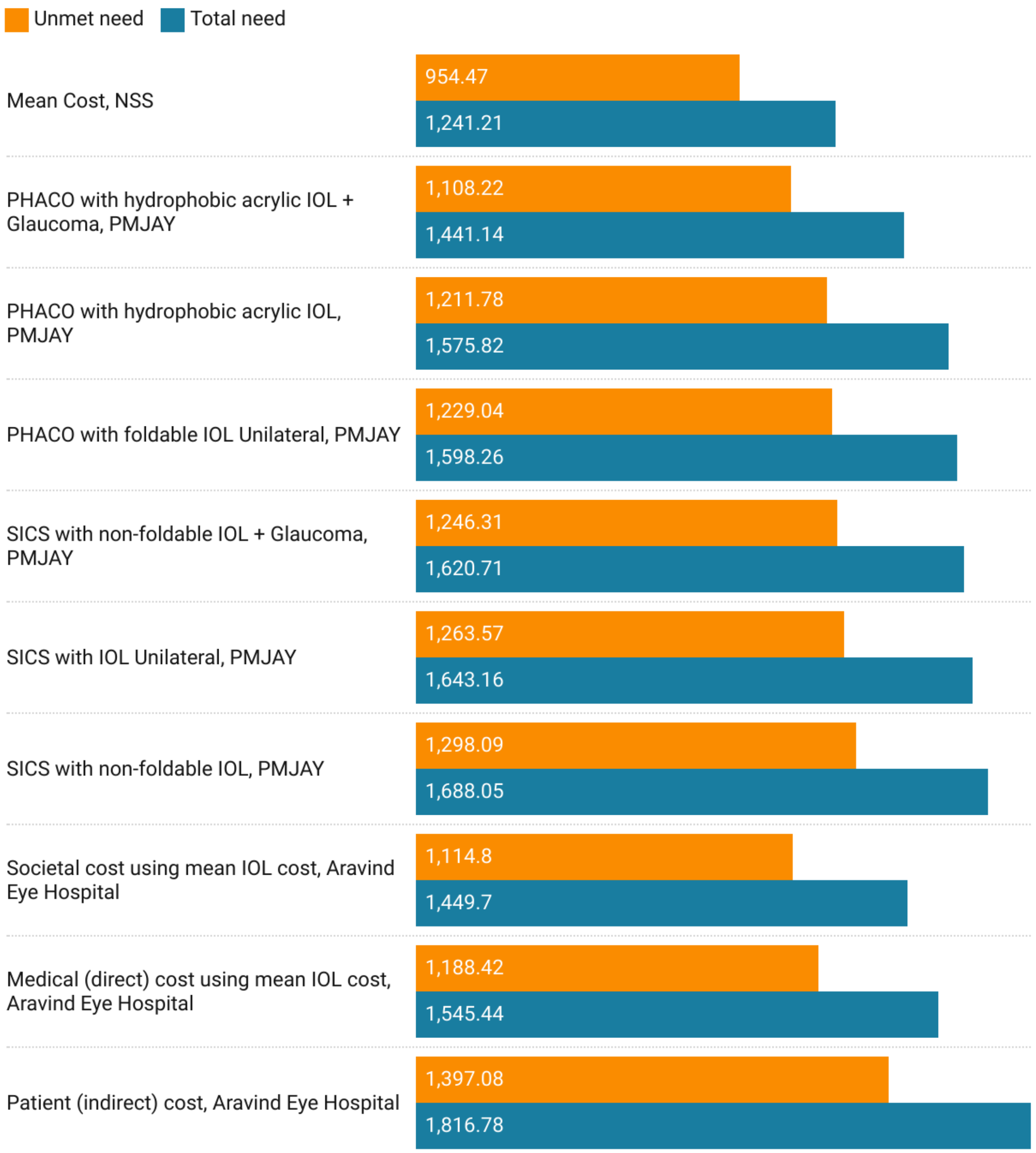


**Supplementary Figure 24: Net Benefits for Meeting Total and Unmet Needs for Universal Cataract Surgical Coverage in Maharashtra.** All costs are adjusted to millions of USD for the year 2020. Positive values depict net benefit while negative values depict net loss. Abbreviations: NSS - National Sample Survey, PMJAY - Pradhan Mantri Jan Arogya Yojana, PHACO - Phacoemulsification, IOL - Intraocular lens, SICS - Small Incision Cataract Surgery.

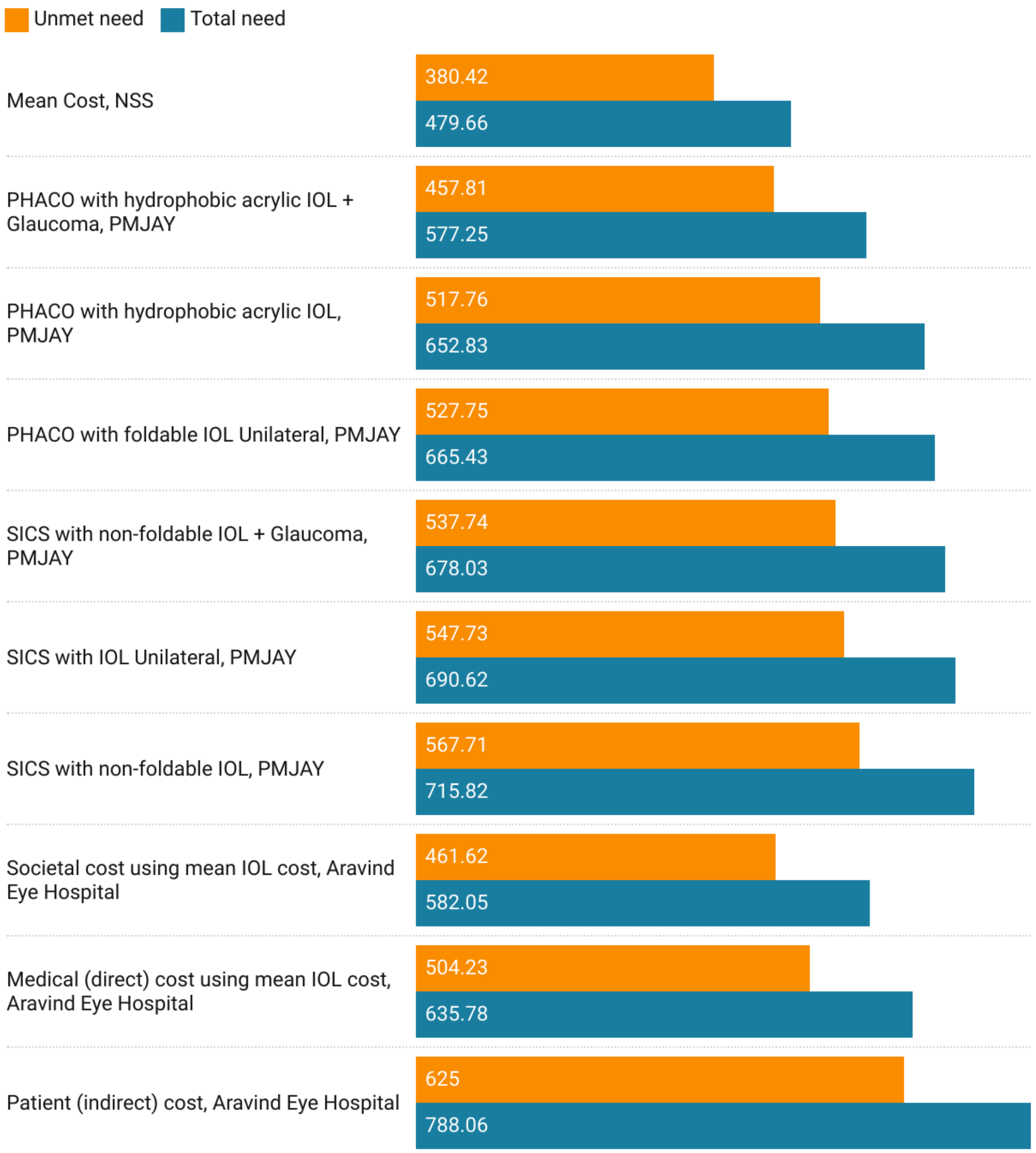


**Supplementary Figure 25: Net Benefits for Meeting Total and Unmet Needs for Universal Cataract Surgical Coverage in Andhra Pradesh.** All costs are adjusted to millions of USD for the year 2020. Positive values depict net benefit while negative values depict net loss. Abbreviations: NSS - National Sample Survey, PMJAY - Pradhan Mantri Jan Arogya Yojana, PHACO - Phacoemulsification, IOL - Intraocular lens, SICS - Small Incision Cataract Surgery.

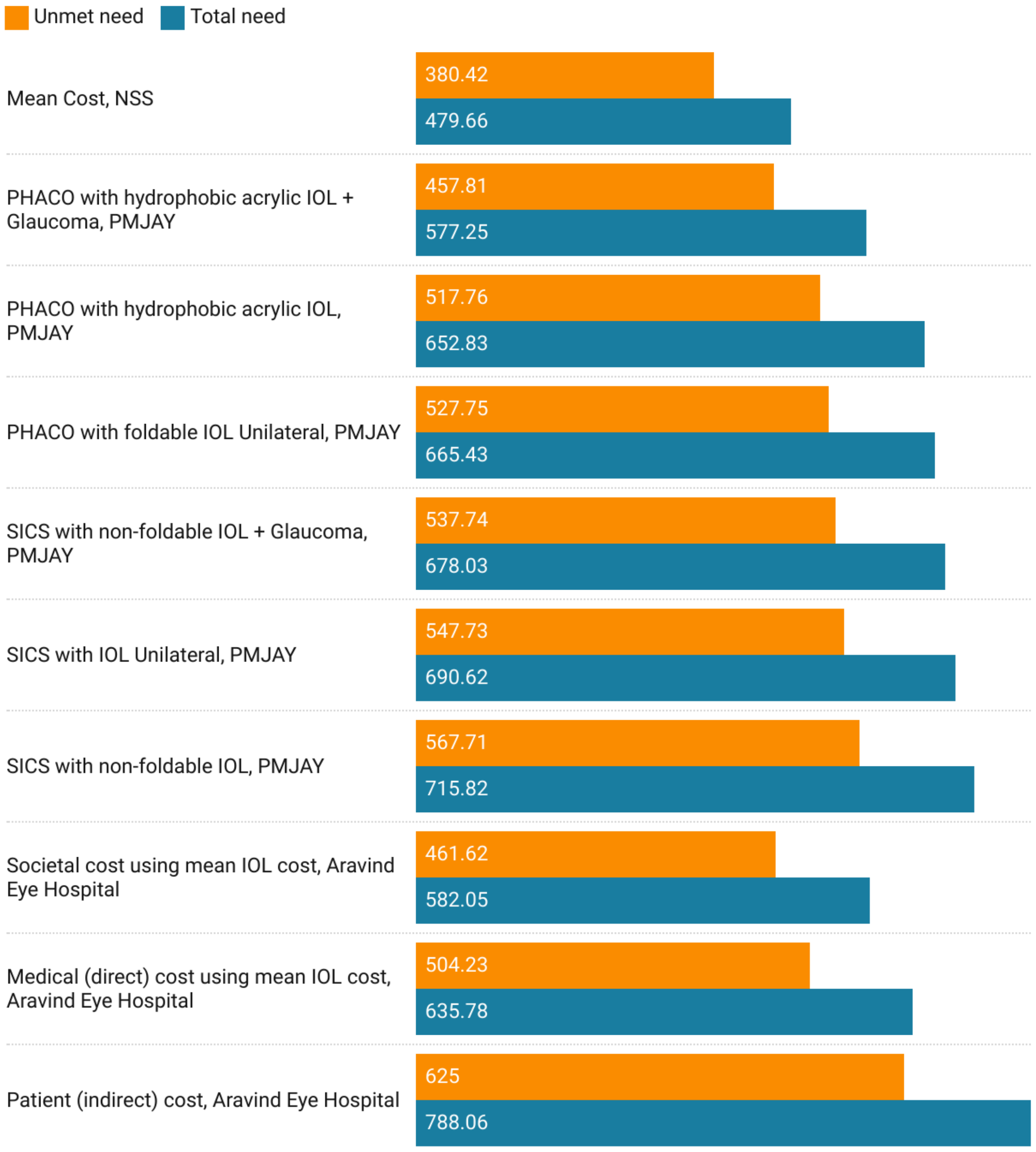


**Supplementary Figure 26: Net Benefits for Meeting Total and Unmet Needs for Universal Cataract Surgical Coverage in Karnataka.** All costs are adjusted to millions of USD for the year 2020. Positive values depict net benefit while negative values depict net loss. Abbreviations: NSS - National Sample Survey, PMJAY - Pradhan Mantri Jan Arogya Yojana, PHACO - Phacoemulsification, IOL - Intraocular lens, SICS - Small Incision Cataract Surgery.

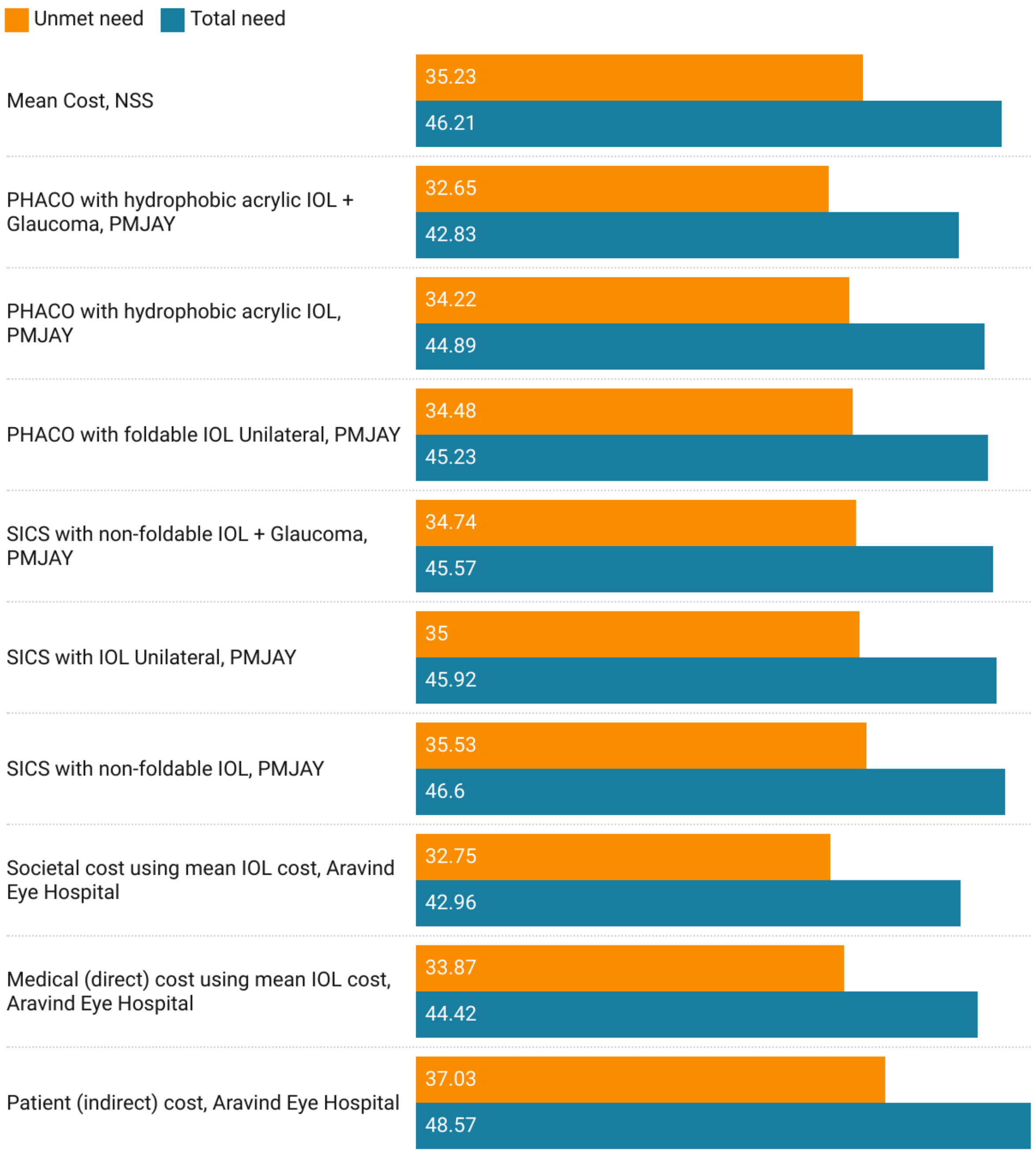


**Supplementary Figure 27: Net Benefits for Meeting Total and Unmet Needs for Universal Cataract Surgical Coverage in Goa.** All costs are adjusted to millions of USD for the year 2020. Positive values depict net benefit while negative values depict net loss. Abbreviations: NSS - National Sample Survey, PMJAY - Pradhan Mantri Jan Arogya Yojana, PHACO - Phacoemulsification, IOL - Intraocular lens, SICS - Small Incision Cataract Surgery.

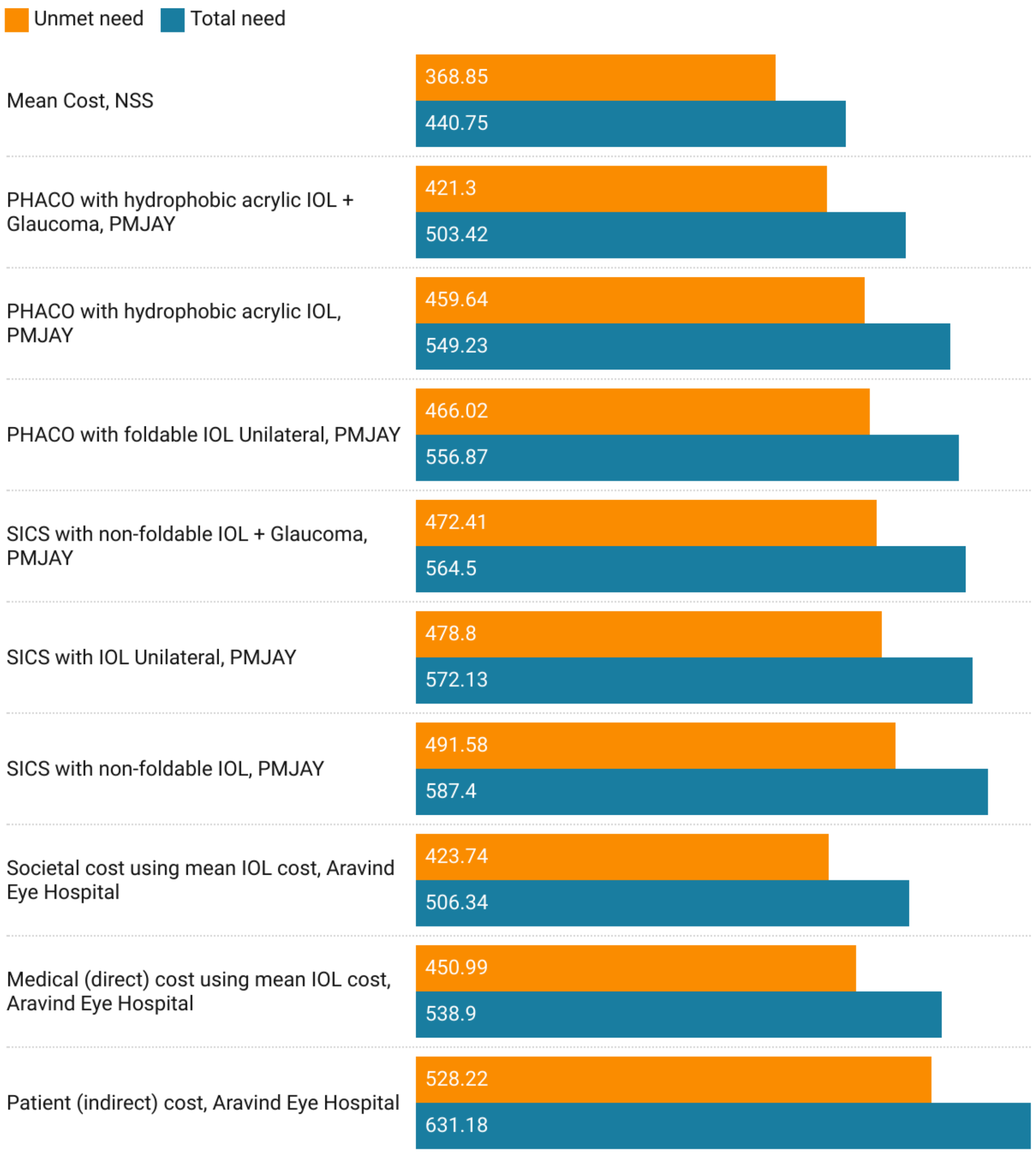


**Supplementary Figure 28: Net Benefits for Meeting Total and Unmet Needs for Universal Cataract Surgical Coverage in Kerala.** All costs are adjusted to millions of USD for the year 2020. Positive values depict net benefit while negative values depict net loss. Abbreviations: NSS - National Sample Survey, PMJAY - Pradhan Mantri Jan Arogya Yojana, PHACO - Phacoemulsification, IOL - Intraocular lens, SICS - Small Incision Cataract Surgery.

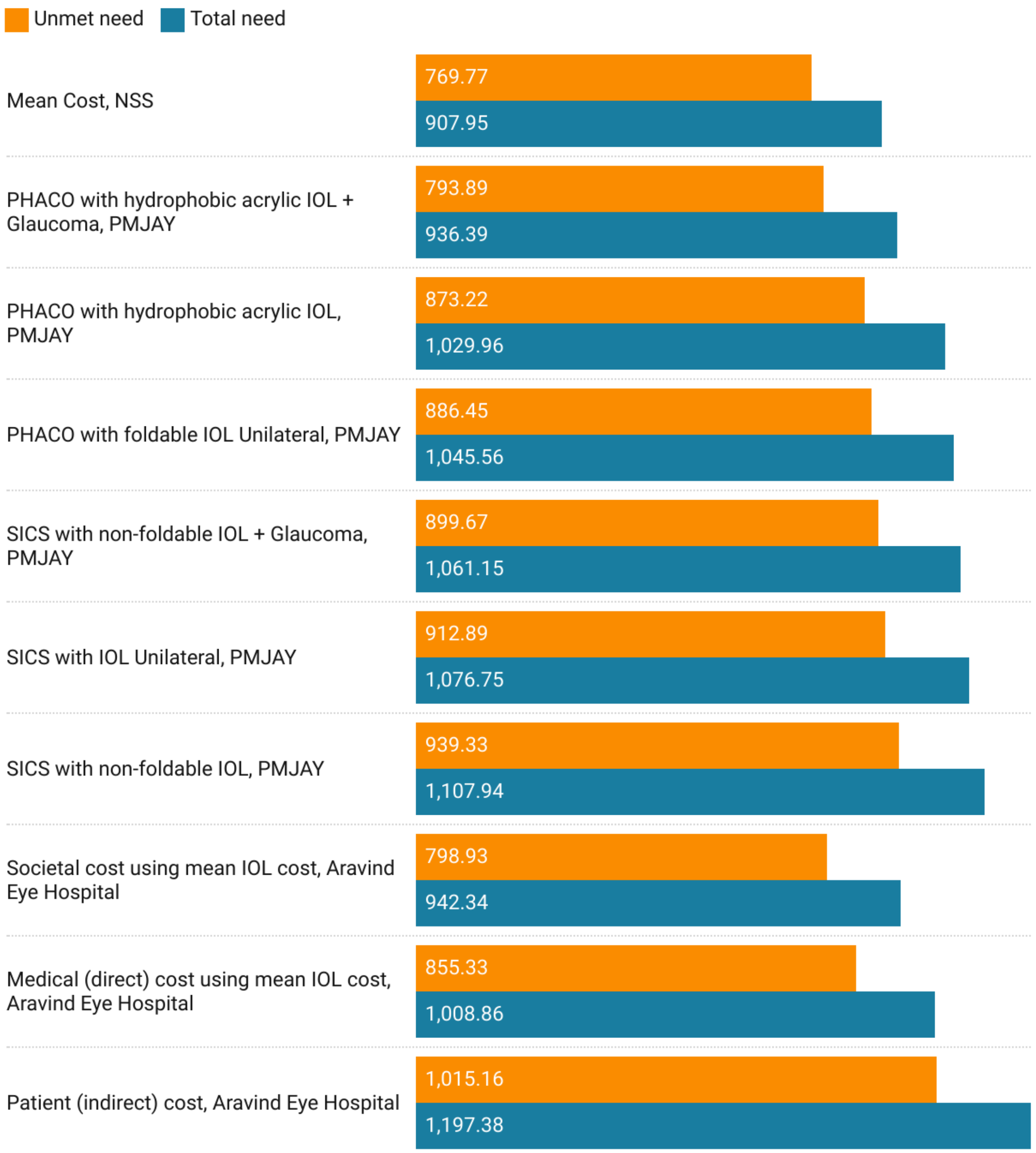


**Supplementary Figure 29: Net Benefits for Meeting Total and Unmet Needs for Universal Cataract Surgical Coverage in Tamil Nadu.** All costs are adjusted to millions of USD for the year 2020. Positive values depict net benefit while negative values depict net loss. Abbreviations: NSS - National Sample Survey, PMJAY - Pradhan Mantri Jan Arogya Yojana, PHACO - Phacoemulsification, IOL - Intraocular lens, SICS - Small Incision Cataract Surgery.

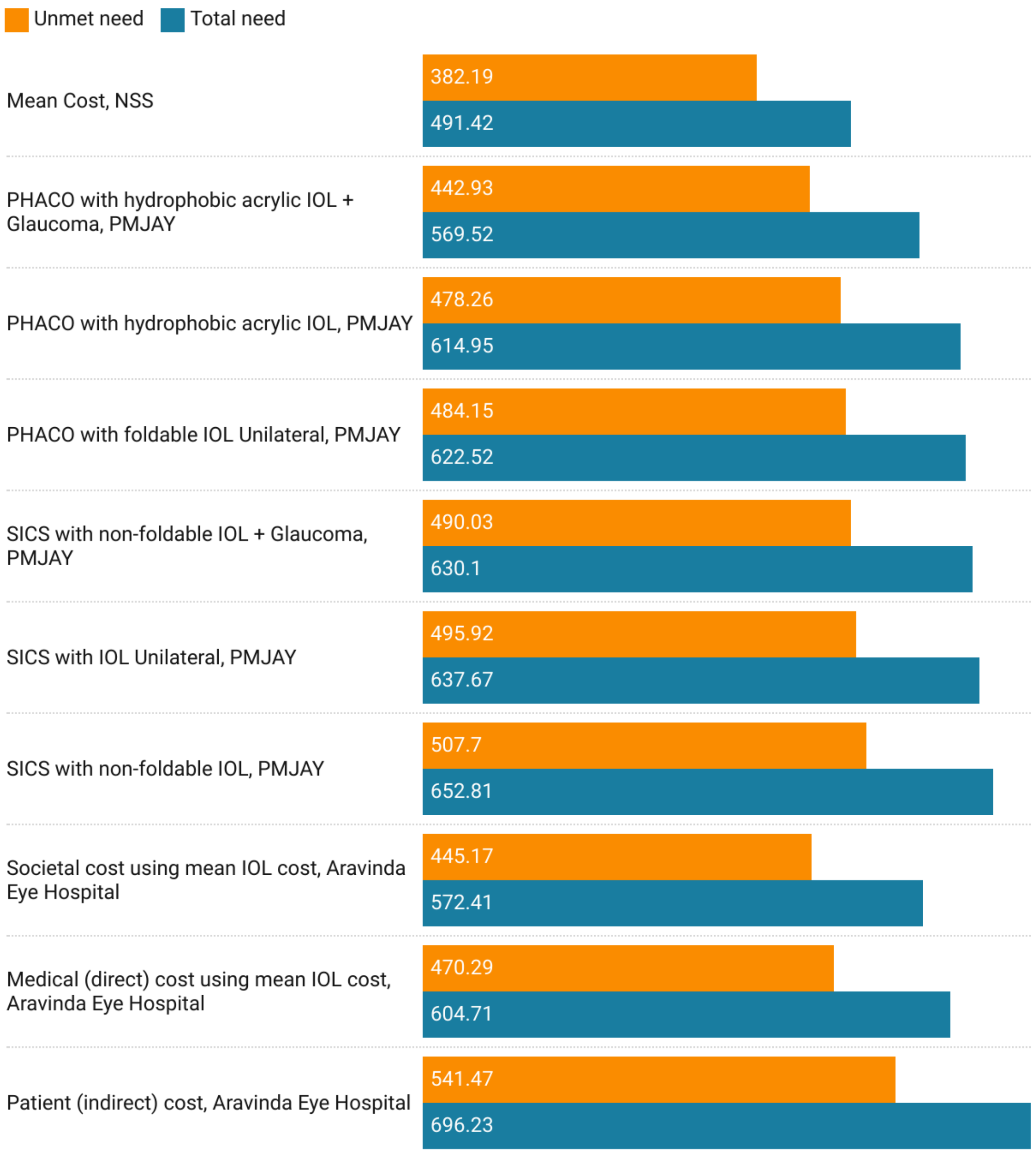


**Supplementary Figure 30: Net Benefits for Meeting Total and Unmet Needs for Universal Cataract Surgical Coverage in Telangana.** All costs are adjusted to millions of USD for the year 2020. Positive values depict net benefit while negative values depict net loss. Abbreviations: NSS - National Sample Survey, PMJAY - Pradhan Mantri Jan Arogya Yojana, PHACO - Phacoemulsification, IOL - Intraocular lens, SICS - Small Incision Cataract Surgery.
